# Impact of socioeconomic status on cardiometabolic multimorbidity progression trajectories: a multi-state model analysis based on three prospective cohort studies

**DOI:** 10.64898/2026.03.09.26347989

**Authors:** Bingchen Zhou, Ping Zeng

## Abstract

**Background:** Cardiometabolic multimorbidity (CMM) has emerged as a persistent global health challenge, yet the role of socioeconomic status (SES) remains insufficiently elucidated. This study aims to investigate the associations between SES and both CMM progression trajectories and mortality risk.

**Methods:** We harmonized data from three UKB, SHARE, and KLoSA. SES was defined based on income, education, and employment via latent class analysis. CMM was defined as the coexistence of two or more cardiometabolic diseases (CMDs), including type 2 diabetes (T2D), coronary artery disease, and stroke. First-onset cardiometabolic disease (FCMD) was defined as the initial manifestation within the spectrum of CMDs. Multi-state models and restricted mean survival time analyses were performed to explore potential underlying relationships.

**Results:** This study included 387,665 UKB participants, 22,505 SHARE participants, and 8,357 KLoSA participants, with median follow-up times of 13.4, 17.3, and 16.0 years, respectively. Individuals with lower SES consistently had higher cumulative risks of disease progression and reached the equivalent 10-year cumulative risk much earlier. Low-SES individuals exhibited a 33.4% higher risk of developing FCMD and a 33.3% increased risk of progressing to CMM compared with the high-SES group. Notably, the strongest socioeconomic disparities in disease progression were observed for the transition from health to T2D (HR=1.45 [1.39-1.52]) and from stroke to CMM (HR=1.51 [1.19-1.93]). Low SES elevated the mortality risk following FCMD (HR=1.41 [1.30-1.54]), which was further exacerbated for transitions from stroke (HR=1.68 [0.95-2.96]) and CMM (HR=1.72 [1.41-2.09]) to death. Across all cohorts, a socioeconomic gradient was evident where disease progression and survival time declined with lower SES, with the onset of FCMD occurring 0.47 (0.16-0.79) years earlier and the progression to CMM occurring 0.76 (0.37-1.14) years earlier in low-SES participants compared to their high-SES counterparts. This trajectory culminated in a significant survival disparity after CMM diagnosis, where medium-SES and low-SES groups lived 1.39 (1.12-1.67) and 1.57 (0.73-2.41) years less, respectively.

**Conclusion:** Significant SES disparities substantially accelerate CMM progression and mortality risk; this association has been consistently observed across populations in various countries. SES intervention strategies should be integrated into CMM assessments to mitigate the adverse health impacts of CMM progression on populations.

**Graphical abstract:** 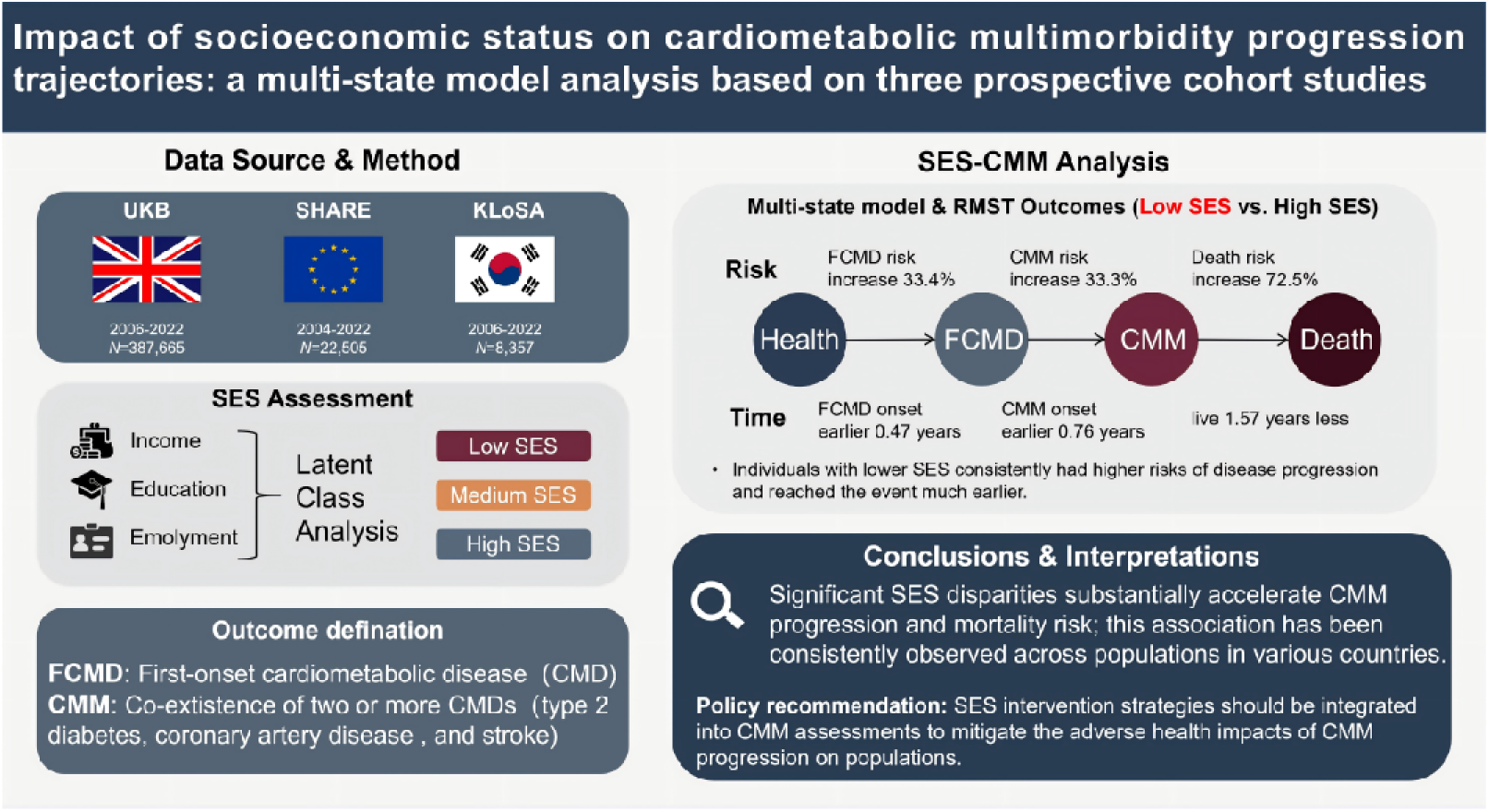

**Research Insights:** *What is currently known about this topic?:* An aging population burden significantly increases CMM prevalence and reduces life expectancy. Lower SES is a known risk factor for cardiovascular disease and general multimorbidity, however its specific impact on CMM risk remains unclear.

*What is the key research question?:* How does lower SES impact the entire developmental trajectory of CMM?

What is new?
Lower SES correlates with increased risks across the entire CMM developmental continuum.
RMST analysis reveals that lower SES significantly accelerates the onset of FCMD and CMM while drastically shortening patient survival time. How might this study influence clinical practice?
Target SES disparities and implement stage specific personalized care for CMM patients.

## Introduction

Cardiometabolic multimorbidity (CMM) is defined as the coexistence of two or three cardiometabolic diseases (CMDs), including type 2 diabetes (T2D), coronary artery disease (CAD), and stroke (Glynn, 2009). With the aging of the global population, CMM has grown increasingly prevalent worldwide, presenting a major challenge to public health (Fan, et al., 2022; Reynolds, et al., 2024). Compared with a single CMD alone, CMM is associated with a multiplicative increase in mortality risk, a significant reduction in life expectancy (Di Angelantonio, et al., 2015), and an elevated healthcare resource utilization (Otieno, et al., 2023). Therefore, identifying potential risk factors for CMM is essential to mitigating its health burden and promoting healthy aging (Jiang, et al., 2023; Jiang, et al., 2023; Yang, et al., 2026; Zhang, et al., 2024).

Despite widespread socioeconomic progress in recent decades, disparities in individual economic circumstances persist, and inequalities in health outcomes across socioeconomic status (SES) remain pronounced. SES reflects an individual’s or group’s relative standing and access to resources within a socioeconomic hierarchy, traditionally differentiated through dimensions such as income, education, or occupation (Antonoplis, 2023). Substantial evidence has identified income (Li, et al., 2025), educational attainment (Magnani, et al., 2024), and employment status (Li, et al., 2022) as significant determinants of CMM, with its incidence and mortality following a distinct socioeconomic gradient whereby individuals with greater resources and higher SES generally experience lower disease risk. Compared to any single indicator, a composite SES measurement better captures the multifaceted nature of socioeconomic disparity and demonstrates superior predictive validity for health outcomes (Darin-Mattsson, et al., 2017; Gong, et al., 2025).

Although extensive research has been conducted, existing studies still leave room for further elucidating the relationship between SES and CMM. First, a predominant focus was placed on examining the association between SES and individual CMDs (Hahad, et al., 2024; Jung, et al., 2025; Pantoja-Ruiz, et al., 2025; Zhang, et al., 2021; Zhu, et al., 2023), or broadly defined multimorbidity (e.g., hypertension, diabetes, coronary heart disease, atherosclerosis, chronic obstructive pulmonary disease, chronic nephritis, stroke, osteoporosis) (Gong, et al., 2025; Mair and Jani, 2020; Zhou, et al., 2024; Zhou, et al., 2024), resulting in a lack of specific investigation dedicated to CMM itself. Second, prior studies largely depended on a single metric, such as occupational status (Pan, et al., 2026; Singh-Manoux, et al., 2018) or area-level Townsend deprivation index (Jiang, et al., 2023), to represent SES (Geyer, et al., 2006), which cannot systematically integrate SES and investigate the synergistic effects of multiple interrelated socioeconomic factors. Thus, constructing a composite index that encompasses the key dimensions of SES is of critical importance. Third, some studies employed simplified measurement scales to assess SES (Li, et al., 2022), which may fail to identify previously unrecognized, prototypical subgroups within the population that share distinct constellations of risk characteristics. Fourth, while the progression of CMM is a dynamic process, previous evidence regarding the association between SES and CMM among hypertensive patients has been limited by a cross-sectional approach (Wang, et al., 2022). Such an investigation focused on a single stage of disease development and lacked large-scale and long-term cohorts capable of capturing the actual dynamic trajectory of disease progression. Consequently, the understanding of the association between SES and CMM remains largely static. To our knowledge, currently, only one study conducted within a

Chinese cohort explicitly investigated the association between SES and CMM (Zhang, et al., 2022). However, its findings were constrained by a relatively small sample size and indicated a positive association between higher SES and an increased risk of first-onset cardiometabolic disease (FCMD), a trend that contrasts with the broader literature (Hahad, et al., 2024; Kivimäki, et al., 2025; Pantoja-Ruiz, et al., 2025; Zhou, et al., 2025; Zhu, et al., 2023). Finally, previous evidence originated primarily from a single population cohort (Jiang, et al., 2023; Zhang, et al., 2022), it lacks sufficient cross-population validation and comparison; further multi-cohort studies are therefore required to examine the generalizability of these associations in populations with varying SES and genetic backgrounds.

Utilizing data from three regionally representative prospective cohorts, including the UK Biobank (UKB) (Sudlow, et al., 2015), the Survey of Health, Ageing and Retirement in Europe (SHARE) (Börsch-Supan, et al., 2013), and the Korean Longitudinal Study of Aging (KLoSA) (Lee, 2020), this study systematically assessed the longitudinal associations between SES and distinct incident stages of CMM. Each of these cohorts has its own unique characteristics; their integration not only increases sample sizes but also provides an opportunity for cross-population verification of the SES-CMM relationship. Latent class analysis (LCA) was first employed to define SES classifications (Bandeen-Roche, et al., 1997; Linzer and Lewis, 2011). Then, by estimating cumulative risk and comparing state transition probabilities between different SES levels, we were able to quantitatively identify critical transition points at which health inequalities emerged. Next, multi-state models (Bühler, et al., 2023; Hougaard, 1999; Le-Rademacher, et al., 2022) were utilized to evaluate the association between SES and the progression of CMM across the three cohorts, which offers the potential to identify critical intervention points for preventing or delaying both the onset and mortality of CMM. Finally, differences in event occurrence time across distinct disease states stratified by SES were quantified using restricted mean survival time (RMST) (Royston and Parmar, 2011). Our findings aim to reveal potential variations in the relationship between SES and CMM across diverse cohorts, and provide a new analytical perspective for formulating tailored interventions and policies targeting specific socioeconomic groups.

## Methods and materials

### Study populations

We utilized data from three prospective cohorts (Figure 1): UKB from 2006 to 2022, SHARE from 2004 to 2022 (waves 1 to 9), and KLoSA from 2006 to 2022 (waves 1 to 9). Each cohort obtained ethical approval, and written informed consent was provided by all participants. Exclusion criteria included: (i) loss to follow-up or withdrawal from the cohort during the study period; (ii) missing data on key socioeconomic factors; or (iii) a pre-existing diagnosis of T2D, CAD, or stroke at baseline, or participants with undocumented event times. The final sample comprised 418,527 individuals, consisting of 387,665 participants from UKB, 22,505 from SHARE, and 8,357 from KLoSA.

**Figure 1.**
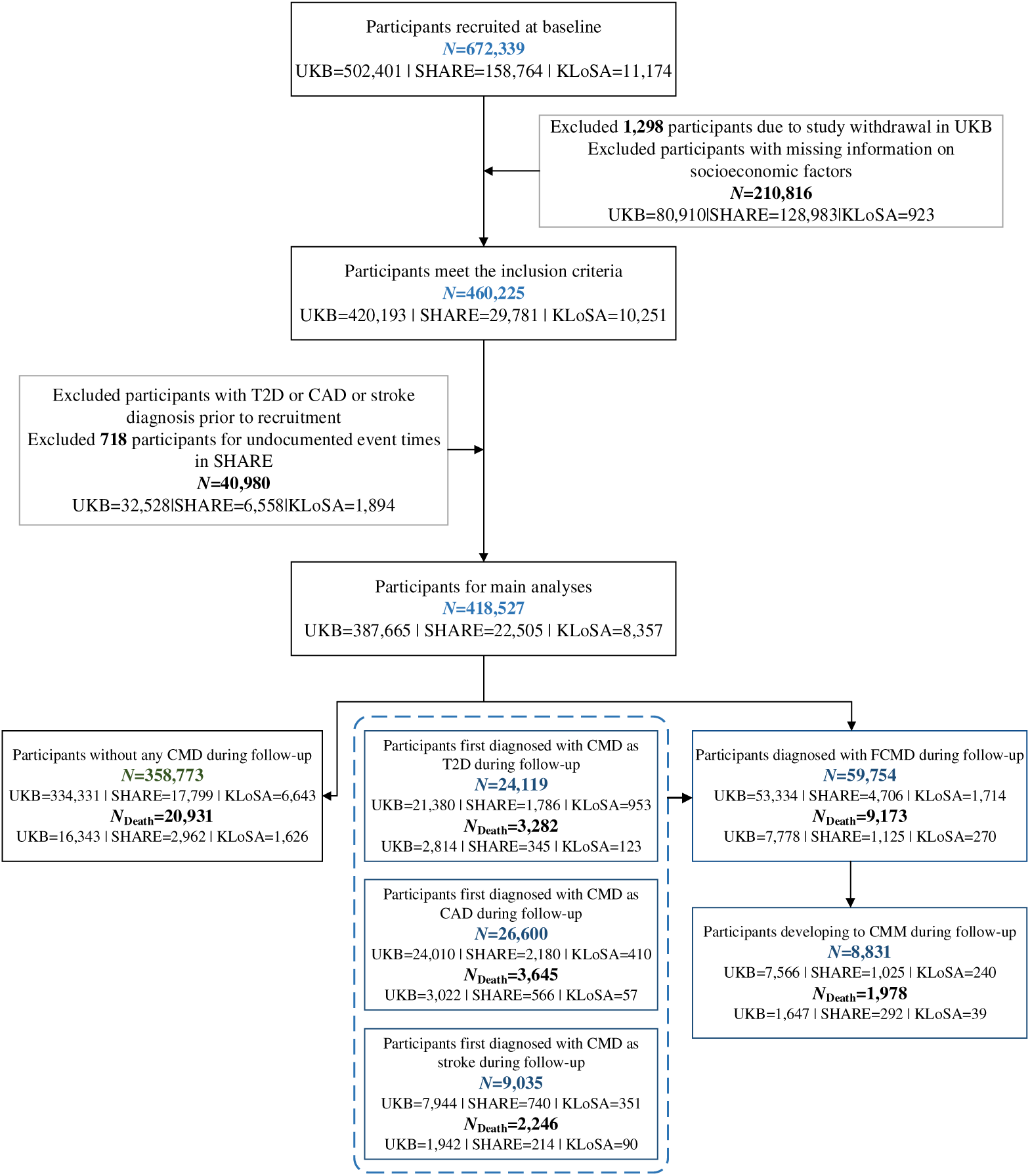
Flow chart of the population selection process. UKB, the UK Biobank; SHARE, the Survey of Health, Ageing and Retirement in Europe; KLoSA, Korean Longitudinal Study of Ageing; T2D, type 2 diabetes; CAD, coronary artery disease; CMD, cardiometabolic disease; FCMD, first-onset cardiometabolic disease; CMM, cardiometabolic multimorbidity.

### Assessment of socioeconomic status

SES was determined based on three baseline variables (Hahad, et al., 2024; Li, et al., 2023; Wang, et al., 2024; Zhang, et al., 2021; Zhou, et al., 2025): income, education, and employment. In UKB, these variables included average total household income before tax, educational qualification, and current employment status. In SHARE, household income was calculated as gross total household income divided by the number of household members and subsequently categorized into quartiles (lowest, lower-middle, upper-middle, and highest); and educational attainment was classified into lower secondary or below, upper secondary, and tertiary education. In KLoSA, we utilized total household income and categorized it into four quartiles (lowest, lower-middle, upper-middle, and highest); educational level was categorized as primary school or below, secondary school, and college or above. In all cohorts, employment status was categorized as employed or unemployed (Supplementary File).

A composite measure of SES was subsequently developed using LCA (Ye, et al., 2023). This method utilizes multiple observed categorical variables to infer an unmeasured latent variable, thereby classifying individuals into a set of mutually exclusive latent classes. We identified three distinct latent classes corresponding to high, medium, and low SES, based on their respective item-response probabilities, with more details provided in the Supplementary File.

### Outcome ascertainment

FCMD was defined as the first diagnosis of any one of T2D, CAD, and stroke (Table S4). CMM onset date was determined by the diagnosis date of the second CMD. In UKB, diagnostic information was sourced from linked hospital records and death registrations across England, Scotland, and Wales. Identification of CMDs was based on ICD-10 codes, supplemented by self-reported illness. In both SHARE and KLoSA, CMDs were ascertained through self-reported information obtained during follow-up survey waves. In each interview, participants were asked whether a doctor had ever informed them that they had, or currently have, diabetes or high blood sugar, heart disease, or stroke. An affirmative response led to the classification of the individual as having the corresponding CMD. The endpoint was defined as the earliest occurring date among the following: confirmed cardiovascular disease event diagnosis, date of death, date of loss to follow-up, or the end of the visit period.

### Available covariates

We included several risk covariates (Table S5), such as age, sex, ethnicity (only UKB), smoking status (Zhang, et al., 2024), alcohol consumption (Pampel, et al., 2010), physical activity (Chudasama, et al., 2019), body mass index (BMI) (Kivimäki, et al., 2017), blood pressure/hypertension (Qin, et al., 2023), and sleep patterns (He, et al., 2023). In SHARE and KLoSA, due to the absence of blood pressure measurements, a prior diagnosis of hypertension at baseline was included as a covariate. To handle missing values in covariates, we employed the imputation by chained equations method (van Buuren and Groothuis-Oudshoorn, 2011).

### Statistical analysis

#### Statistical description

Baseline characteristics were stratified by FCMD and CMM status. Continuous variables were reported as mean ± standard deviation (SD), and categorical variables as frequency and percentages.

#### Cumulative transition probability under various SES levels

To reflect the long-term transition probability from a healthy state to either a disease state or death within the CMM framework, we first estimated the crude cumulative incidence of CMM for distinct SES groups within each state transition of Transition Model A (Figure 2) using the Kaplan-Meier method (Kaplan and Meier, 1958).

**Figure 2.**
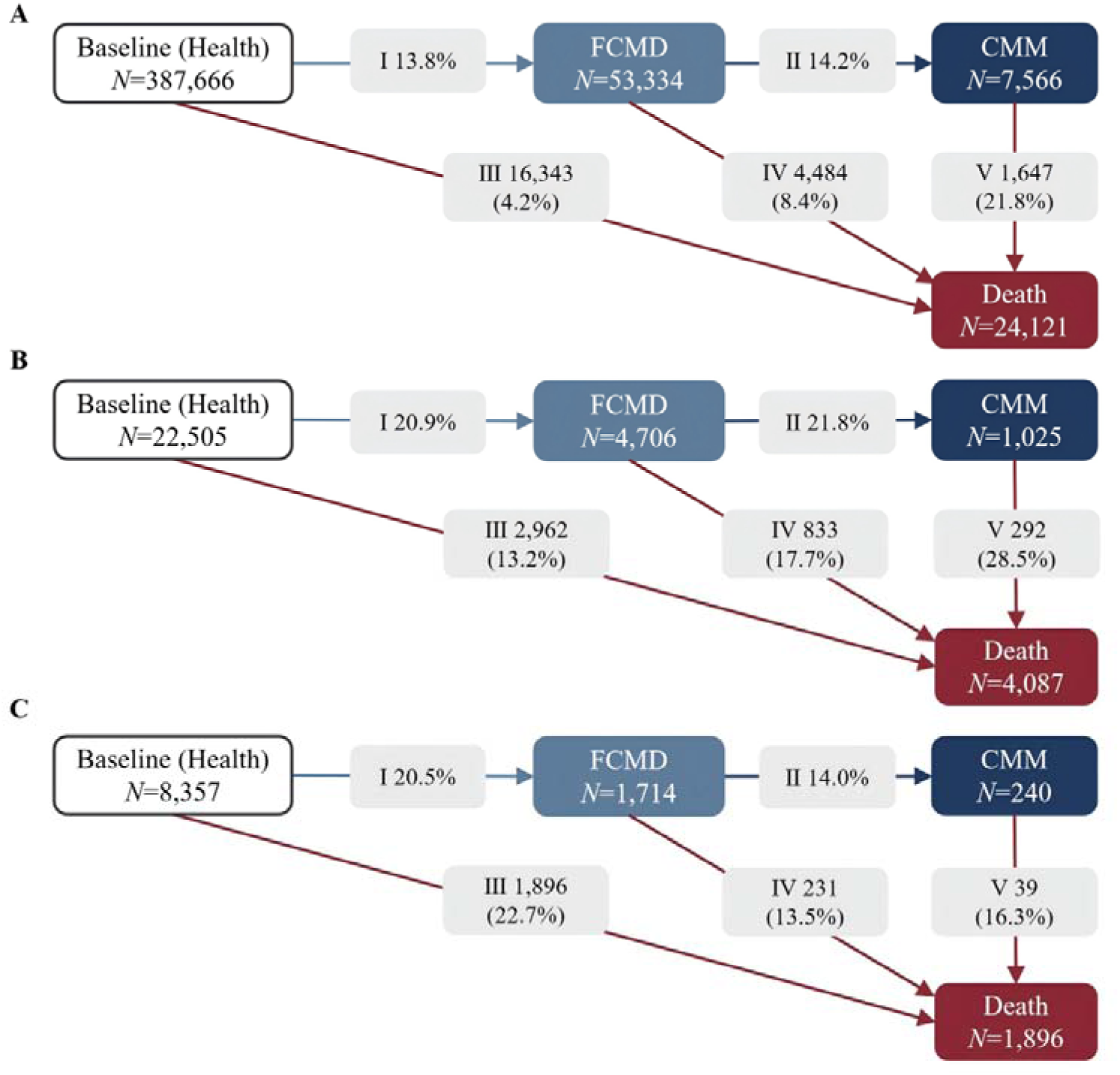
Disease progression pattern for multi-state model A in (A) UKB, (B) SHARE, and (C) KLoSA. FCMD, first-onset cardiometabolic disease; CMM, cardiometabolic multimorbidity.

#### Multi-state model for assessing the effect of SES

We employed Transition Model A (Figure 2) to evaluate the impact of SES on CMM progression through five state transitions: (I) baseline → FCMD; (II) FCMD → CMM; (III) baseline → death; (IV) FCMD → death; (V) CMM → death. Death was designated as an absorbing state. To refine disease-specific associations, we subdivided the baseline → FCMD transition into three sub-pathways (T2D, CAD, stroke), resulting in Transition Model B (Figures 3) with a total of 11 transitions.

**Figure 3.**
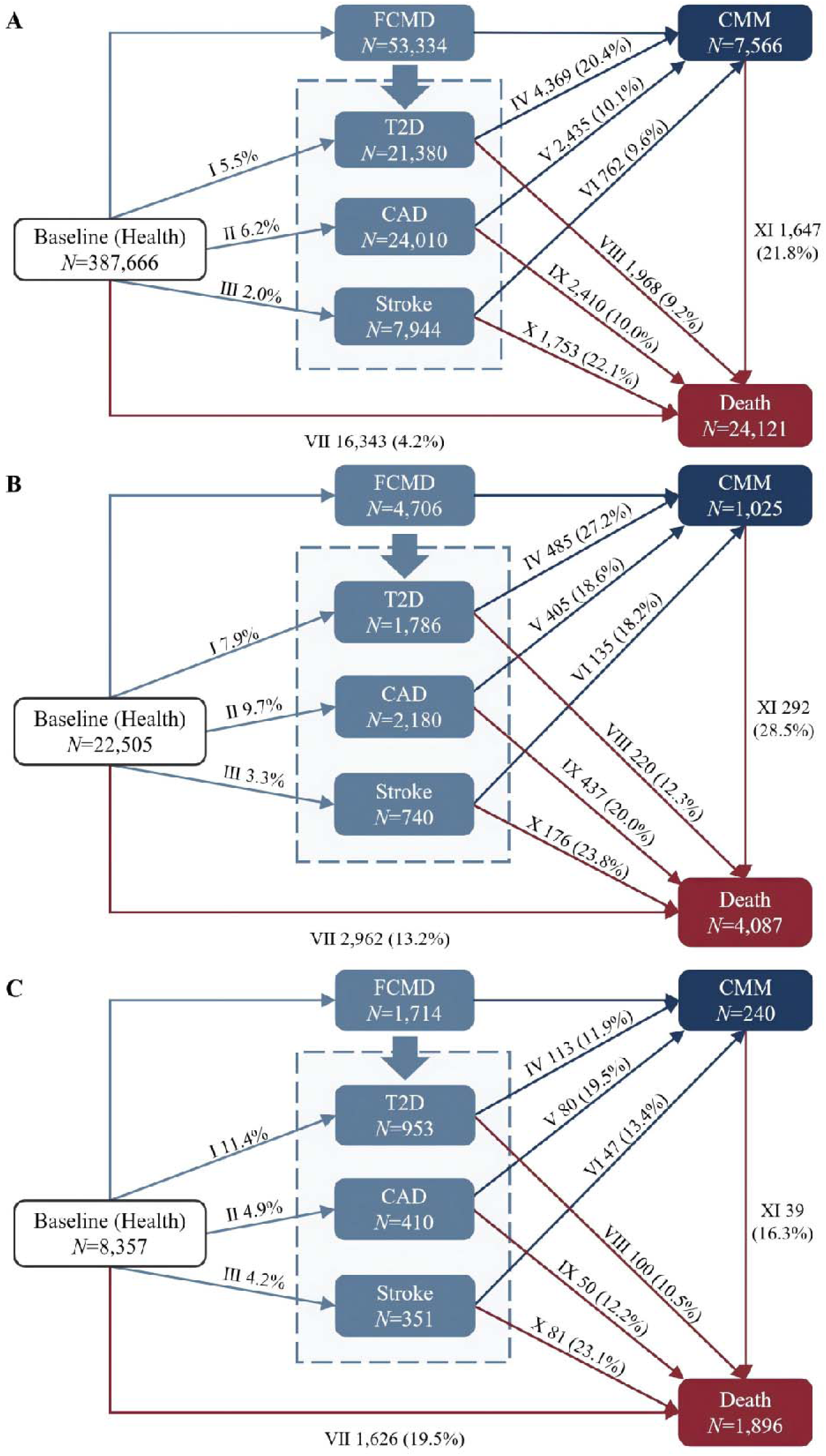
Disease progression pattern for multi-state model B for (A) UKB, (B) SHARE, and (C) KLoSA. T2D, type 2 diabetes; CAD, coronary artery disease; FCMD, first-onset cardiometabolic disease; CMM, cardiometabolic multimorbidity.

According to previous studies (Han, et al., 2021; Pan, et al., 2025), for individuals who were diagnosed with two diseases on the same day, we operationally defined the theoretical entry date of the preceding state as 0.5 days before the date of the subsequent state, to determine the sequence of disease occurrence in the order of T2D, CAD, and stroke (Jiang, et al., 2024).

Covariate-adjusted multi-state models were used to estimate the effect of SES, using the high SES group as the reference. Hazard ratio (HR) and corresponding 95% confidence interval (CI) were reported in each cohort; then, an inverse-variance weighted meta-analysis approach was applied to yield an overall effect estimate across different cohorts(Maitra, 2025). Heterogeneity in the cohort-specific estimates was assessed via Cochran’s *Q* test and the I² statistic.

To evaluate the adequacy of the sample sizes and the observed number of transition events across the three cohorts, we conducted a power analysis (Hsieh and Lavori, 2000). With the actual observed number of transition events, the total sample size of the risk set, and the allocation ratio between the comparison groups, the statistical power was independently estimated for detecting pre-specified HRs (1.25, 1.50, and 2.00) when comparing lower SES with the high SES reference group for each of the five predefined multi-state transition paths. A statistical power of 80% was considered adequate to reliably detect the corresponding effect sizes.

#### Disease progression time and survival time stratified by SES

We calculated RMST for the five pathways in Model A as the average time spent in a specific state before transitioning to a subsequent state (Andersen, et al., 2004; Han and Jung, 2022; Tian, et al., 2014); that is, RMST = f S(t) dt, where *S*(*t*) is the covariate-adjusted survival function. To ensure the stability of the RMST estimates, we selected τ=13 years, a time that was approximately equal to the median follow-up time in UKB and minimal among the three cohorts to guarantee that the size of the risk set was large enough in each cohort. Specifically, the disease progression time was defined as the RMST from a specific state to disease progression, whereas the survival time was defined as the RMST from a given state until death. We quantified between-SES disparities in RMST separately in every cohort and employed an inverse-variance weighted meta-analysis to derive overall estimates.

#### Subgroup and sensitivity analyses

To validate robustness and examine heterogeneity among the three cohorts (Head, et al., 2020), we conducted stratified analyses by sex (male and female), smoking status (never vs. former or current smoker), alcohol consumption (more than once per week vs. less than once per week), physical activity (low-to-moderate vs. high), sleep pattern (healthy vs. unhealthy), and age (<60 years and ≥60 years, WHO-defined elderly).

Sensitivity analyses conducted in three cohorts included: (1) decomposing SES into mutually adjusted income, occupation and education components to account for potential graded associations between distinct socioeconomic factors and the progression of CMM; (2) excluding early events (<3-year follow-up) to address reverse influence.

We conducted inverse-variance weighted meta-analyses on subgroup and sensitivity analyses across the three cohorts. Due to the limited sample size in KLoSA, meta-analyses for alcohol consumption stratification and for the exclusion of cases with disease onset within the initial three years of follow-up were not performed.

#### Software and R packages

Statistical analyses were performed using R software (version 4.3.2). All statistical tests were two-sided, with *P*<0.05 considered statistically significant. Multiple imputation of missing data was conducted using the mice package (van Buuren and Groothuis-Oudshoorn, 2011). SES indicators were derived using the poLCA package (Linzer and Lewis, 2011). Power analyses were performed with the powerSurvEpi package. RMST analyses were performed with the survRM2 package (Uno, et al., 2014), while multi-state modeling was carried out with the mstate package (Putter, et al., 2007). Meta-analyses were executed using the meta package (Balduzzi, et al., 2019).

## Results

### Participant characteristics

The final sample comprised 418,527 individuals, including 387,665 participants from UKB (43.5% males, mean age 55.8 years), 22,505 from SHARE (41.9% males, mean age 62.5 years), and 8,357 from KLoSA (42.5% males, mean age 60.7 years). The median follow-up time was 13.4 years (IQR: 12.7∼14.1 years) for UKB, 17.3 years (IQR: 16.5∼17.6 years) for SHARE, and 16.0 years (IQR: 15.8∼16.1 years) for KLOSA. The UKB cohort documented 53,334 FCMD, 7,566 CMM and 24,121 death cases; the corresponding numbers were 4,706, 1,025, and 4,087 in SHARE, and 1,714, 240, and 1,896 in KLoSA.

The baseline characteristics of these cohorts indicated that individuals with FCMD and CMM were more likely to be older, male, have hypertension or higher blood pressure, report poor sleep quality, engage in less physical activity, have a higher BMI, be current or former smokers, and have a lower SES compared to those without these conditions (Tables S6-S8).

### Estimated cumulative risk curve

A progressive increase in cumulative incidence over time was accompanied by a widening gap in risk across various SES levels (Figure 4). The transition probabilities at 5-, 10- and 15- years are detailed in Table S9, we focused here on the 10-year comparison between high and low SES groups because this time point captured significant disparities while maintaining a stable sample size representative of long-term disease progression.

**Figure 4.**
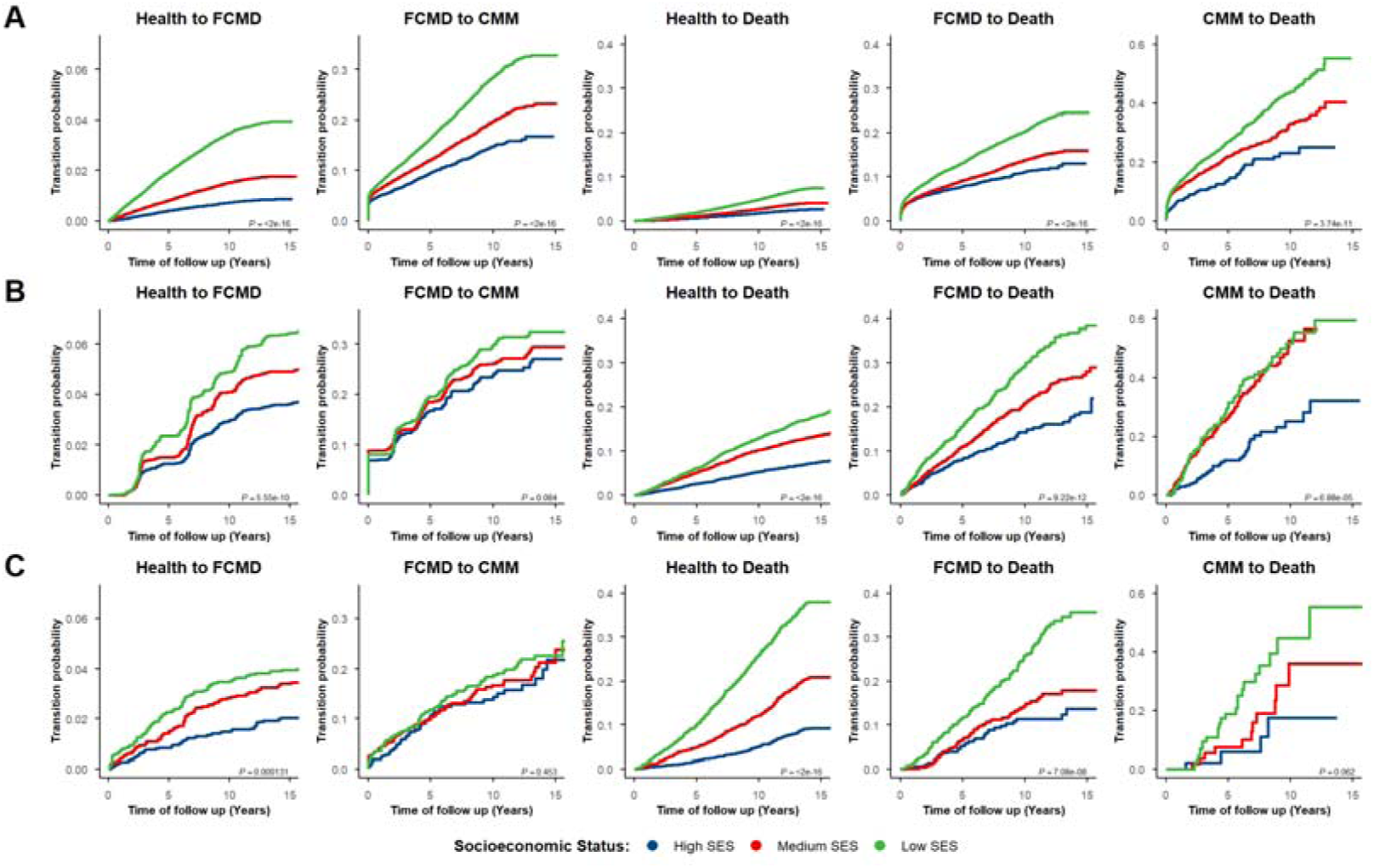
Cumulative incidence of CMM transitions by SES for (A) UKB, (B) SHARE, and (C) KLoSA. FCMD, first-onset cardiometabolic disease; CMM, cardiometabolic multimorbidity.

First, in the transition from health to FCMD, the differences in cumulative risk between high- and low-SES individuals were minimal across all transitions, ranging from 1.9% (SHARE) to 2.8% (UKB). Here, the high-SES group consistently showed the lowest cumulative risk. Second, patients with FCMD from low-SES backgrounds exhibited a higher risk of progressing to CMM, the difference in cumulative risk of progression to CMM was 13.6% in UKB, in contrast to 6.0% in SHARE and 4.3% in KLoSA.

Third, the risk of direct progression from health to death was elevated in low-SES groups, with a difference as high as 20.8% in KLoSA, compared to 3.1% in UKB and 7.8% in SHARE. Among patients with FCMD, the differences in cumulative mortality were 9.2% in UKB, 15.1% in SHARE, and 14.5% in KLoSA. Fourth, for patients with CMM, the mortality in the low-SES group was 43.9% in UKB, 51.5% in SHARE, and 44.1% in KLoSA. The differences in cumulative mortality between high- and low-SES CMM patients were 21.0% in UKB, 26.6% in SHARE, and 27.2% in KLoSA.

Importantly, the existence of one CMD or CMM significantly amplified the effect of SES on disease burden. For instance, across the three cohorts, low-SES patients with FCMD had a 6.5-fold higher risk of developing another CMD compared to healthy individuals. In terms of mortality, low-SES patients with FCMD faced a 2.5-fold greater risk, while those with CMM had a 5.0-fold higher risk relative to healthy individuals. In addition, low-SES patients with CMM exhibited a 2.0-fold higher mortality risk than those with FCMD.

On average, during the transition from health to FCMD across these cohorts (Table S10), low-SE individuals reached the equivalent 10-year cumulative risk of their high-SES counterparts 6.2 years earlier (an example is shown in the top left of Figure 4). This accelerated disease progression persisted into the subsequent stage, with the low-SES group developing a 10-year risk of CMM 4.3 years sooner than the high-SES group. The impact of socioeconomic disparities was equally evident across all mortality pathways. Among individuals transitioning directly from health to death, the low-SES group reached the 10-year mortality risk of the high-SES group 6.0 years earlier. Among those already diagnosed with FCMD, the low-SES group attained the equivalent 10-year mortality risk 5.7 years ahead of the high-SES group. Finally, for those progressing from CMM to death, the time to reach the corresponding mortality risk was advanced by 5.6 years in the low-SES group compared to the high-SES group.

### Results of multi-state regression

As shown in Figure 5, we found that a lower SES level typically demonstrated a significant dose-response relationship with higher risks of FCMD, CMM and death in most transition phases (*P*_trend_<0.05); and these associations were largely homogenous across the three cohorts (*P*_heterogeneity_>0.05).

**Figure 5.**
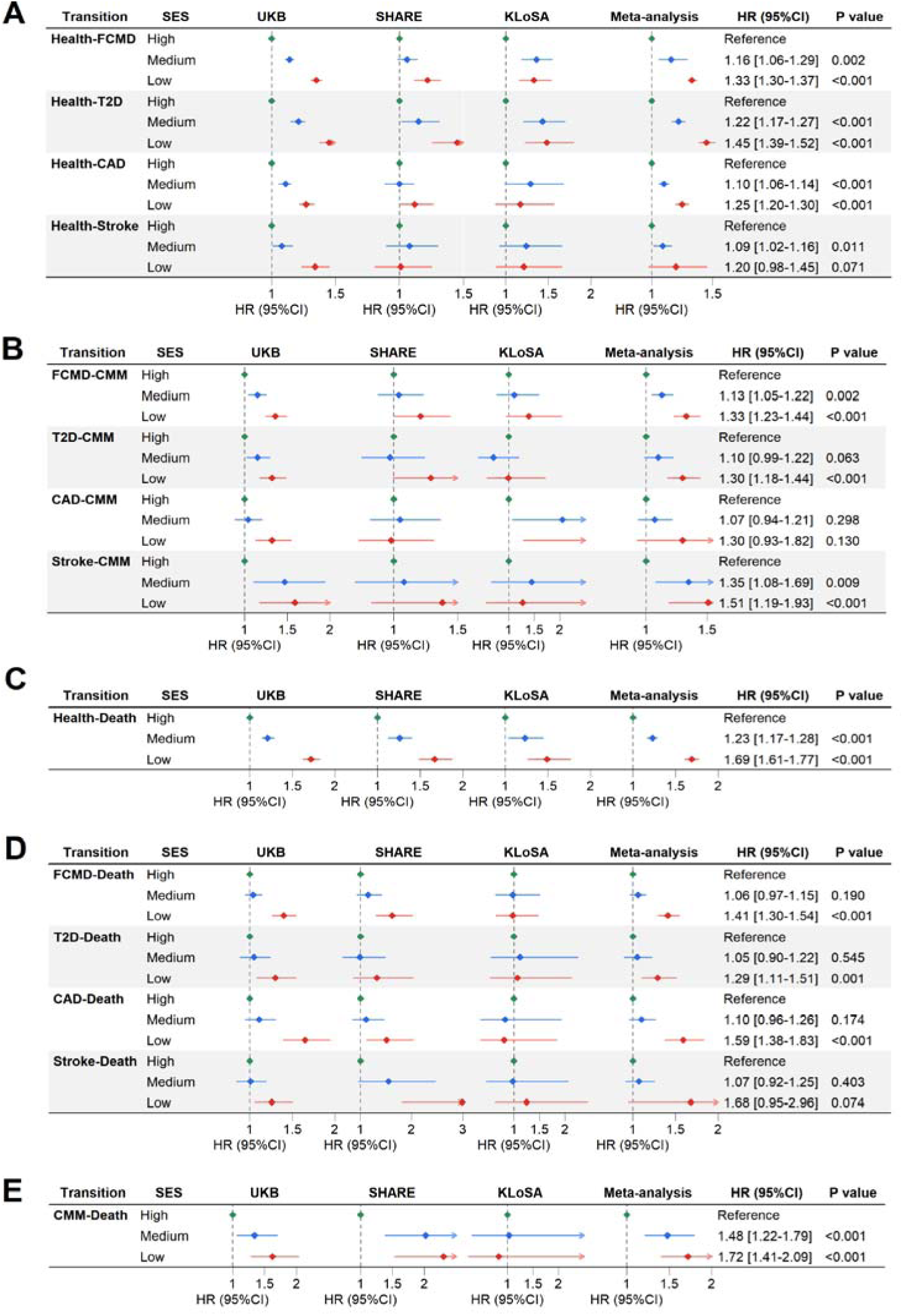
Forest plots of association of SES levels with multiple-state transitions. The reported HR (95%CI) and *P*-values are based on the meta-analysis results. SES, socioeconomic status; T2D, type 2 diabetes; CAD, coronary artery disease; FCMD, first-onset cardiometabolic disease; CMM, cardiometabolic multimorbidity.

In the transition from health to FCMD (Figure 5A and Table S11), compared to high SES, medium SES was related to an elevated FCMD risk in UKB (HR=1.14 [1.11-1.17]) and KLoSA (HR=1.36 [1.19-1.54]), but not in SHARE (*P*=0.110), indicating notable heterogeneity regarding the impact of medium SES across cohorts (*P*_heterogeneity_=0.005). Especially, low SES was associated with a 33.4% greater risk in this stage (HR_meta_=1.33 [1.30-1.37]), a finding that was robustly significant (*P*<0.05) across all three cohorts. Among distinct FCMD subtypes, T2D was more affected by lower SES, showing a 23.1% higher risk for low SES and an approximately 12.6% greater risk for medium SES compared to the risk of CAD or stroke. Moreover, a stable and statistically significant increase in progression risk was observed throughout the transition from health to T2D across all the three cohorts (*P*<0.05); whereas the relatively higher risk of CAD and stroke due to lower SES was only identified in UKB.

The risks of transitioning from FCMD to CMM also increased as the SES level decreased (HR_meta_=1.13 [1.05-1.22] for medium SES; HR_meta_=1.33 [1.23-1.44] for low SES) (Figure 5B and Table S12); however, these significant relationships were only present in UKB or SHARE. Particularly, lower-SES participants who were first diagnosed with stroke were more likely to develop CMM (low SES: HR_meta_=1.51 [1.19-1.93]; medium SES: HR_meta_=1.35 [1.08-1.69]) compared to T2D and stroke patients.

The risk of death from a healthy state (Figure 5C and Table S13) was 69.1% higher for low-SES participants (HR_meta_=1.69 [1.61-1.77]) and 22.1% greater for those with medium SES (HR_meta_=1.23 [1.17-1.28]), both of which were highly consistent across three cohorts.

In the FCMD-death stage (Figure 5D and Table S14), low SES was associated with a 41.2% higher risk (HR_meta_=1.41 [1.30-1.54]), whereas this significance was only detected in UKB and SHARE. In contrast, medium SES did not show a substantial influence on death risk (*P*=0.190). Particularly, low SES was related to a higher mortality risk among CAD (HR_meta_=1.59 [1.38-1.83]) and stroke (HR_meta_=1.68 [0.95-2.96]) patients, although the latter was only marginally significant (*P*=0.074). Additionally, the substantial impacts of low SES were observed in UKB (HR=1.26 [1.06-1.50]) as well as in SHARE (HR=2.99 [1.82-4.89]). Very interesting, all of the SES on death impacts in this transition were not significant in KLoSA.

For transition from CMM to death (Figure 5E and Table S15), participants with medium and low SES levels were associated with 47.7% (HR_meta_=1.48 [1.22-1.79]) and 71.7% (HR_meta_=1.72 [1.41-2.09]) increased risks, respectively; but such associations were nonsignificant in KLoSA (*P*_medium-SES_=0.926 and *P*_low-SES_=0.771).

The power analysis for Transition Model A is shown in Figure S4. For the given SES effects, we achieved a moderate-to-high power for all of the state transitions in UKB and SHARE; whereas we could not exclude the possibility of low power in some transitions of KLoSA (e.g., FCMD → CMM) due to limited events. However, the insignificant and weak effects of SES observed in the CMM-to-death and FCMD-to-death transitions of KLoSA seemed not to be explained by low power alone.

### Disease progression time and survival time

Figure 6 and Table S17 demonstrate a gradient where lower SES levels correlated with accelerated progression toward FCMD, CMM, and death in all three cohorts, with RMST differences expanding significantly alongside disease progression. However, there are significant heterogeneities in the effects of SES on multiple paths across these cohort (*P*_heterogeneity_<0.05).

**Figure 6.**
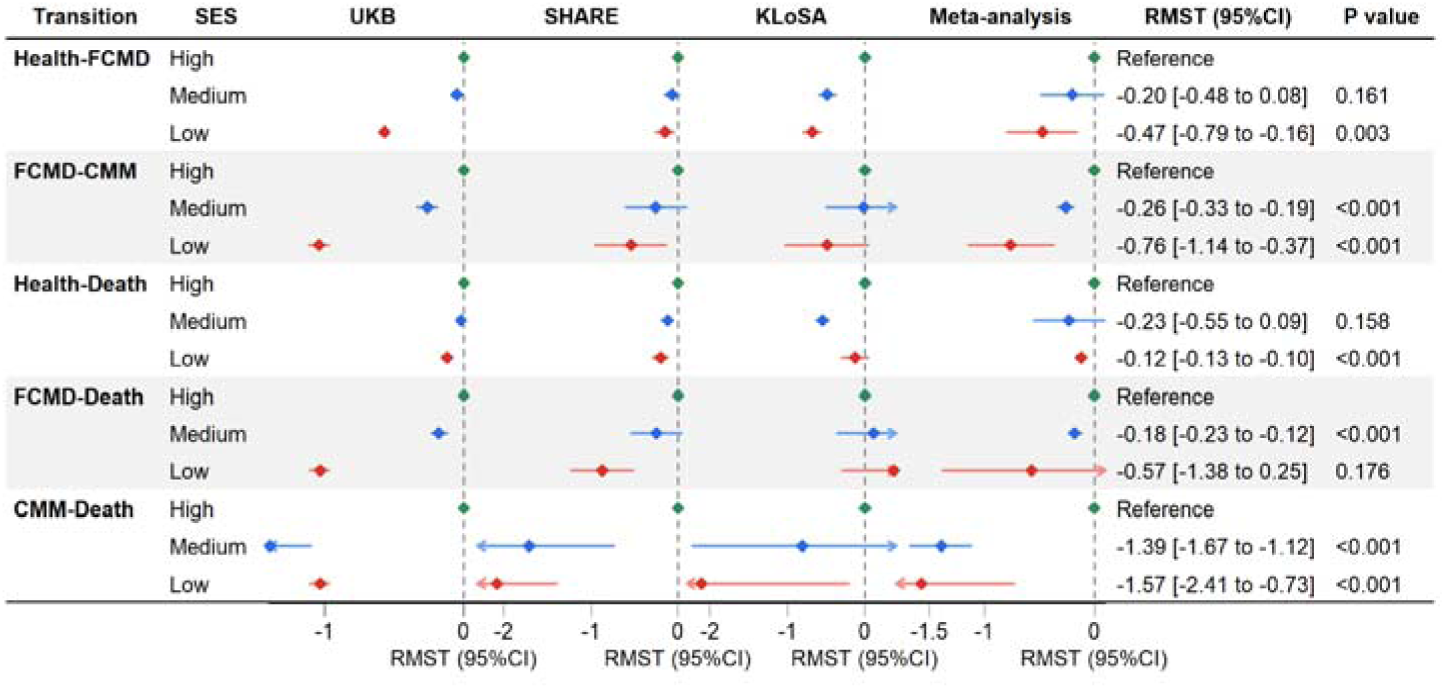
Forest plots of RMST difference (in years) stratified by transitions and SES. The reported RMST (95%CI) and *P*-values are based on the meta-analysis results. SES, socioeconomic status; FCMD, first-onset cardiometabolic disease; CMM, cardiometabolic multimorbidity.

Compared to high-SES participants, the medium- and low-SES groups showed 0.20 (0.08-0.48) years and 0.47 (0.16-0.79) years earlier onset of FCMD, respectively.

These observations were consistently significant in all three cohorts (*P*<0.05); for instance, on average, the early onset of FCMD for low-SES participants was 0.57 (0.55-0.60) years in UKB, 0.15 (0.05-0.26) years in SHARE, and 0.68 (0.57-0.80) years in KLoSA.

The socioeconomic disparity was also shown during the transition from FCMD to CMM (*P*_heterogeneity_=0.129), where the progression to CMM was accelerated by 0.76 (0.37-1.14) years for low-SES participants relative to high-SES counterparts. Notably, while significant reductions in disease progression time of this transition were observed among low-SES participants in both UKB (1.04 [0.97-1.11] years) and SHARE (0.54 [0.14-0.95] years), the decrease was non-significant in KLoSA (*P*=0.072).

The impact of SES was most pronounced in mortality transitions. In the health-death stage, the survival time of low-SES participants reduced about 0.12 (0.10-0.13) years compared to high-SES ones. In the transition from FCMD to death, medium-SES participants lived 0.18 (0.12-0.23) years shorter than the high-SES group. Low-SES participants experienced a substantially larger reduction that was significant in UKB and SHARE (*P*<0.05), with the notable exception of KLoSA which demonstrated a non-significant change in survival time (*P*=0.273). The disparity of survival time due to SES peaked in the CMM to death trajectory, where medium- and low-SES participants exhibited substantial survival deficits of 1.39 (1.12-1.67) years and 1.57 (0.73-2.41) years, respectively. Notably, in this transition low-SES individuals exhibited a significant survival time loss of 2.08 (1.40-2.75) years in SHARE and 2.11 (0.22-4.01) years in KLoSA, approximately doubling the loss of 1.03 (0.97-1.10) years observed in UKB.

### Subgroup and sensitivity analyses

Stratified analyses confirmed significant effect modification of SES disparities on CMD progression. The association between low SES and mortality among patients with CMM was particularly stronger in individuals under 60 years of age (HR=2.53 [1.69-3.79]) than in those aged 60 or above (HR=1.48 [1.19-1.86]) (Tables S17-S20). Among patients with FCMD, the risk of progression to CMM associated with low SES was relatively smaller in females (HR=1.26 [1.08-1.46]) than in males (HR=1.37 [1.25-1.51]) (Tables S21-S24). Concurrently, some modifiable behavioral factors exacerbated SES gradients. For instance, smoking (Tables S25-S28) was associated with a significantly higher risk of progression from CMM to death (HR=1.90 [1.42-2.56]). Similarly, high alcohol consumption amplified this risk in UKB (HR=1.71 [1.32-2.23]) and SHARE (HR=2.78 [0.82-9.43]) cohorts (Tables S29-S30).

Moreover, unhealthy sleep patterns (HR=1.90 [1.40-2.58]) and physical inactivity (HR=1.79 [1.39-2.31]) also contributed to elevated risks for this transition (Tables S31-S38).

In the analysis examining individual SES components (Tables S39-42), income was significantly associated with nearly all pathways of CMM progression. Specifically, low income was associated with an increased risk in the transition from CAD to death (HR=1.25 [1.09-1.44]). Education showed significant associations across all stages of CMM transition, with the strongest effect observed for the transition from health to death (HR=1.16 [1.01-1.32]). Occupation was significantly associated with the transitions from T2D to death and from CMM to death (HR=1.38 [1.17-1.63]). After excluding cases with the disease in the first three years, robust associations between the SES level and CMM progression were still observed (Tables S43-S44).

## Discussion

### Summary of our study

Across three prospective cohorts including UKB, SHARE and KLoSA, we demonstrated that the increased risk linked to lower SES spanned the full spectrum of CMM development, from onset and progression to prognosis and mortality, peaking in the critical stage from CMM to death. However, in disease-specific transitions, the impact of SES varied depending on disease types. Especially, the strongest effect of SES was observed on T2D for the transition from baseline to FCMD, on stroke for the transition from FCMD to CMM and death. Lower SES was also linked to early onset, reduced survival time and marked disparities in cumulative incidence throughout the disease cascade, a pattern consistently observed across cohorts and most pronounced in the progression from CMM to death.

### Comparison with previous studies

#### Findings in the dynamic modeling framework of CMM

The adverse effects of SES on some transitions of CMM observed here were similar to prior studies, which, however, only reported the association between SES and CMD. For instance, epidemiological studies have established the link between SES and an increased risk of incident CMDs (Schultz, et al., 2018; Zhang, et al., 2021), which aligns with our observation of elevated risk for the transition from baseline health to FCMD.

We found that the association between SES and both the incidence of FCMD and subsequent progression to CMM varied according to specific disease types. For example, the influence of SES on the transition from health to FCMD was most obvious for T2D, which is consistent with a prior study reporting a stronger association between SES and the incidence of T2D compared to major cardiovascular diseases (Wang, et al., 2024).

Regarding the transition from specific FCMD to CMM, previous studies have reported increased cardiovascular risk associated with lower SES among T2D patients (Enguita-Germán, et al., 2024; Safieddine, et al., 2023), suggesting an important role of socioeconomic factors in the progression from FCMD condition to CMM. In the present work, the impact of SES was most pronounced among stroke patients, exceeding the effects observed in individuals with T2D or CAD. Similarly, within the FCMD-to-death transition, the influence of SES remained most evident for those in the stroke subgroup. This phenomenon may be attributable to common post-stroke sequelae (McHutchison, et al., 2019; Pantoja-Ruiz, et al., 2025), such as mobility impairment, cognitive dysfunction, and depression, which necessitate assistance with activities of daily living. Low SES could contribute to a higher likelihood of developing CMM and mortality in these vulnerable patients (Lindmark, et al., 2024). Consequently, ensuring timely and adequate treatment especially for stroke patients is essential to mitigate the burden of CMM in low-SES populations. Furthermore, our results highlighted that the association between low-SES and progression from CMM to death is particularly strong, suggesting that low SES exacerbates the mortality risk associated with CMM (Di Angelantonio, et al., 2015).

While our analysis demonstrated consistent socioeconomic inequalities across all three cohorts, the temporal dynamics of these disparities diverged significantly throughout the disease trajectories. In UKB and SHARE, we observed a pattern of cumulative disadvantage, where the impact of low SES intensified with disease progression, culminating in substantial survival gaps during the transition from FCMD to death and CMM to death. In stark contrast, this SES gradient seemed to be null in KLoSA during the same critical phase. We hypothesize that this phenomenon may be partly attributable to the expanded health insurance coverage policy in Korea, which drastically reduces co-payments for major diseases (Ji, et al., 2024). This policy intervention appears to largely decouple economic deprivation from mortality risk at the terminal stage, even though low SES continues to drive the initial onset of disease.

Notably, a study conducted within CLHLS reported that high SES was associated with a greater risk in the transition from health to FCMD, as well as in the transitions from FCMD to death and from CMM to death (Zhang, et al., 2022), representing a discrepancy with our conclusions. The precise reasons for this inconsistency remain unclear but may be attributable to methodological and population differences. Specifically, this prior study defined SES using a principal component of six economy-related variables (education years, sufficient financial source, housing type, own bedroom, pension, and self-reported economic condition), whereas we adopted a framework integrating income, education, and employment. Moreover, their focus on the unique oldest-old cohort from the CLHLS may restrict the broader generalizability of their results.

#### Findings of cumulative risk and RMST

We observed that the cumulative risk among individuals with lower SES increased at a faster rate over time throughout the CMM progression cascade compared to their high-SES counterparts, demonstrating a clear socioeconomic divergence. This accelerating disparity was particularly pronounced in both the progression from FCMD to CMM and in the associated mortality. Notably, the cumulative risk of death following CMM exceeded 50% at 15 years for the low-SES group across all three cohorts, underscoring the urgent need for timely prognostic interventions. We found that individuals with lower SES experienced both an earlier onset and a higher incidence of progression within the CMM cascade, implying that efforts to mitigate socioeconomic disparities may not only reduce the incidence of CMM outcomes among patients with FCMD but also prolong life free of such outcomes.

Our study further demonstrated lower SES contributed to a reduction in RMST estimates throughout the CMM progression cascade, a pattern consistent with prior evidence (Davies, et al., 2022; Lyons, et al., 2023). Notably, individuals with low-SES exhibited the most pronounced reduction in survival time during the transition from CMM to death, indicating a more deteriorated situation after illness for these socioeconomically disadvantaged populations.

### Potential reasons of the association between SES and CMM

Our findings indicate that low SES significantly exacerbates the mortality risk associated with CMM, an association likely driven by multidimensional mechanisms (Lu, et al., 2024). First, behavioral factors may play a pivotal mediating role (Zhang, et al., 2021). Low SES often constrains social support, lifestyle choices and health literacy, leading to a clustering of adverse health behaviors (Pathirana and Jackson, 2018; Wang, et al., 2022; Wang, et al., 2025; Wang, et al., 2025; Zhang, et al., 2020). Furthermore, chronic stress resulting from prolonged exposure to socioeconomic adversity can precipitate neuroendocrine dysregulation, financial strain, heightened loneliness (Jespersen, et al., 2025), and a depletion of personal resources and social competence, all of which may detrimentally impact emotional well-being (Pavlova, 2021; Qualter, et al., 2021). Additionally, during disease progression, disparities in healthcare accessibility are critical; individuals with low SES often encounter diagnostic delays and treatment discontinuities (Lindmark, et al., 2024), thereby accelerating the transition toward CMM and mortality. Collectively, these mechanisms constitute a cumulative disadvantage that ultimately contributes to the elevated CMM burden and mortality risk observed among socioeconomically disadvantaged populations.

### Public health implications

From a public health perspective, this study significantly expands the existing evidence base by establishing a robust link between SES and the entire trajectory of CMM progression across diverse cohorts. These findings underscore the necessity of explicitly integrating SES into CMM burden assessments and preventive strategies (Hahad, et al., 2024; Wang, et al., 2022; Wang, et al., 2025). Practically, the accessibility of individual-level SES renders it an ideal metric for risk stratification in primary care. The elevated risk observed among baseline healthy individuals specifically supports incorporating SES data into community health records and routine check-ups (Platzer, et al., 2023), thereby facilitating proactive management and the efficient allocation of public health resources. Against the backdrop of rapidly aging global populations, implementing targeted policies to narrow income disparities, enhance educational attainment, optimize employment conditions, and strengthen medical security is pivotal for mitigating disease risks and extending healthy life expectancy. Furthermore, recognizing the transitions from FCMD and CMM to death as critical windows of vulnerability underscores the imperative to optimize resource allocation and intensify tertiary prevention efforts during these phases (Zhu, et al., 2023), and targeted upstream structural interventions are essential to maximize survival outcomes and extend life expectancy (Wang, et al., 2025). Routine inclusion of SES in prevention frameworks would thus enhance risk stratification and potentially mitigate the societal burden of CMM at its source.

### Strengths and limitations

Our study demonstrates evident advantages over previous research. First, to our knowledge, this study represents the first multi-state analysis utilizing multiple cohorts to differentiate the influence of SES on distinct progression stages of CMM. This approach effectively addresses limitations inherent in traditional Cox models concerning competing risks and intermediate health states (Putter, et al., 2007), enabling an assessment of the role of SES throughout CMM progression. Second, the analysis incorporated three mature prospective cohorts. Long-term follow-up ensured the availability of complete outcome data, and the concordance of results across these independent cohorts strengthened the validity of the findings. Third, we constructed a composite SES indicator, which facilitated adjustment for confounding factors and allowed a comprehensive evaluation of the complex relationship between SES and the stages of CMM development (Darin-Mattsson, et al., 2017). Fourth, the extended follow-up duration enabled the acquisition of complete health outcome data (Börsch-Supan, et al., 2013; Lee, 2020; Sudlow, et al., 2015). Finally, we conducted a series of sensitivity analyses to demonstrate the robustness of the findings.

Several important limitations warrant consideration. First, the application of multi-state models relies heavily on precise definitions of disease progression states and prespecified clinical transition sequences. Data on CMDs in SHARE were calculated based on participant waves, potentially introducing information bias. Inaccuracies in determining the temporal order of disease stages may lead to misclassification bias, compromising the validity of transition probability estimates. Second, heterogeneity in the definition of CMDs across cohorts may affect the comparability of the results. Specifically, CMDs was defined using ICD codes in UKB, whereas in the other two cohorts it was based on self-reported diagnoses of diabetes, stroke, and heart problems, potentially introducing heterogeneity in endpoint ascertainment. Third, the potential healthy volunteer effect in these cohorts may reduce the general population representativeness of the results (Fry, et al., 2017). Fourth, our reliance on baseline SES measurements restricted our ability to capture the effects of time-varying socioeconomic mobility on disease trajectories. Fifth, the complex sociological and biological pathways underlying the association between SES and CMM transitions are not fully elucidated. Finally, as an observational study, it cannot establish solid causal relationships (Hess, 2023). Additionally, although we adjusted for a comprehensive set of covariates, potential residual confounding from unmeasured factors, such as genetic susceptibility and early-life environmental exposures, remains a concern.

## Conclusions

The findings from three large cohorts, including UKB, SHARE and KLoSA, indicate that lower SES is significantly associated with the occurrence of CMM, accelerates the progression of multimorbidity and increases mortality. These findings help screen individuals at high risk for CMM and related mortality, underscoring the necessity of implementing targeted interventions tailored to different socioeconomic groups to slow disease progression and alleviate the population burden of CMM.

## Additional File

Supplementary File

## Abbreviations

SES: socioeconomic status
CMD: cardiometabolic disease
CMM: cardiometabolic multimorbidity
FCMD: first-onset cardiometabolic disease
T2D: type 2 diabetes
CAD: coronary artery disease
HR: hazard ratio
CI: confidence interval
RMST: restricted mean survival time
UKB: UK Biobank
SHARE: Survey of Health, Ageing and Retirement in Europe
KLOSA: Korean Longitudinal Study of Ageing
LCA: latent class analysis BMI body mass index
SD: standard deviation

## Declarations

### Acknowledgments

This study was partly based on the UK Biobank resource under application number 88159. The UK Biobank was established by the Wellcome Trust medical charity, Medical Research Council, Department of Health, Scottish Government, and the Northwest Regional Development Agency. It has also had funding from the Welsh Assembly Government, British Heart Foundation and Diabetes UK. This study also relied on SHARE and KLOSA, the authors express their great gratitude to these research teams and all individuals who participated in the studies. The data analyses in the present study were carried out with the high-performance computing cluster that was supported by the special central finance project of local universities for Xuzhou Medical University.

## Authors’ contributions

PZ conceived the idea for the study. PZ obtained the data. BCZ cleared up the datasets; BCZ performed the data analyses. PZ and BCZ interpreted the results of the data analyses. PZ and BCZ wrote the manuscript with the help from other authors. All authors read and approved the final manuscript.

## Availability of data and materials

All data generated or analyzed during this study are included in this published article and its supplementary information files. This study used the UK Biobank resource with the application ID 88159. Researchers can access to the UK Biobank by applying to the UK Biobank official website (https://www.ukbiobank.ac.uk/). The SHARE and KLOSA data were publicly available from https://releases.sharedataportal.eu/, and https://survey.keis.or.kr/eng/index.jsp, respectively.

## Ethics approval and consent to participate

The original surveys granted ethical approval and were conducted in accordance with the principles of the Declaration of Helsinki. Informed consent was obtained from all participants using the original surveys.

## Consent for publication

All authors have approved the manuscript and given their consent for submission and publication.

## Data Availability

https://survey.keis.or.kr/eng/index.jsp

https://releases.sharedataportal.eu/

