## Supplementary File for "Impact of socioeconomic status on cardiometabolic multimorbidity progression trajectories: a multi-state model analysis based on three prospective cohort studies"

### Assessment of socioeconomic factors

In the UK Biobank (UKB) cohort, all information on socioeconomic was obtained through questionnaires. All questionnaires could be obtained through the website (https://biobank.ndph.ox.ac.uk/showcase/index.cgi).

- Family income level: Total household income before tax was obtained through questionnaires, and participants could choose an option from “less than ₤18,000”, “₤18,000 to 30,999”, “₤31,000 to 51,999”, “₤52,000 to 100,000”, “greater than ₤100,000”, “do not know”, or “prefer not to answer”.
- Education attainment: Education qualification was obtained through questionnaires, and participants reported their education qualifications as “College or university degree”, “A levels/AS levels or equivalent”, “O levels/GCSEs or equivalent”, “CSEs or equivalent”, “NVQ or HND or HNC or equivalent”, “Other professional qualifications”, “None of the above” (equivalent to less than high school diploma), or “Prefer not to answer”.
- Employment status: The UKB only acquired employment status instead of specific occupation information at baseline through questionnaires, and participants could report their employment status as in paid employment or self-employed, retired, doing unpaid or voluntary work, being full or part-time students, looking after home/family, unable to work because of sickness or disability, unemployed, none of the above, or prefer not to answer. Those who choose the first four options were grouped into the “employed” group, while others (except for the last two options, which were treated as missing values) were grouped into the “unemployed” group.

In the Survey of Health, Ageing and Retirement in Europe (SHARE) cohort, socioeconomic information was obtained through questionnaires. All questionnaires could be obtained through the website (https://share-eric.eu/data/data-access).

- Household income: Household total income (before-tax) was obtained through questionnaires at the household-level (all respondents in the household). Household per capita income was derived by dividing the gross total household income by the household size, obtained as continuous values provided by the respondents.
- Education attainment: Participants reported their education qualifications as none, primary education, lower secondary education, upper secondary education, post-secondary non tertia, first stage of tertiary e, and second stage of tertiary. Those who choose the first three options were grouped into the “lower secondary education or below” group, last two options were grouped into the “tertiary education” group, while others were grouped into the “upper secondary education” group.
- Employment status: Participants could report their employment status as employed or self-employed, retired, unemployed, permanently sick or disabled, homemakers. Those who choose the first three options were grouped into the “employed” group, while others were grouped into the “unemployed” group. Don't know, refuse, and other missing responses are ignored.

In the Korean Longitudinal Study of Aging (KLoSA), socioeconomic information was obtained through questionnaires. All questionnaires could be obtained through the website (http://survey.keis.or.kr/eng/klosa/klosa01.jsp).

- Household income: Total household income (before-tax) refers to the combined income of all household members, including the respondent. These data were reported by participants as a continuous numerical value.
- Education attainment: Participants were required to report their highest educational attainment by selecting from the following options: no education (illiterate), no education (reading), elementary school, middle school, high school, two-year college, college graduate, post college (Master), and post college (PhD). Based on their selections, individuals were categorized into three groups: those choosing the first three options were classified as “primary school or below”; those selecting the last four options were grouped as "tertiary education"; and the remaining participants were assigned to the “secondary education” group.
- Employment status: Participants were asked whether they were currently engaged in paid work. Responses were coded numerically, with a value of 0 indicating the participant was not currently employed in paid work and a value of 1 indicating current engagement in paid employment.

### Assessment of socioeconomic status using latent class analysis

In the UKB cohort, pre-tax gross household income, educational qualifications, and employment status were used to generate an overall SES parameter. We did not consider medical insurance in the UK since it operates a National Health Service system designed to provide comprehensive, universal, and free medical services. Participants were divided into five groups based on pre-tax gross household income, namely “less than £18,000”, “£18,000 to £30,999”, “£31,000 to £51,999”, “£52,000 to £100,000”, and “more than £100,000”. Educational qualifications were categorized into seven groups, including “College or University Degree”, “A level/AS levels or Equivalent”, “O level/GCSEs or Equivalent”, “CSEs or equivalent”, “NVQ or HND or HNC or Equivalent”, “Other Professional Qualifications”, and “None of the Above” (equivalent to below high school diploma). The UKB only obtained employment status rather than specific occupational information at baseline. We regrouped participants into two groups: employed (including those in paid employment or self-employed, retired, doing unpaid or voluntary work, full or part-time student) and unemployed (including looking after home and/or family, unable to work because of sickness or disability, unemployed).

In the SHARE cohort, household income, educational qualifications, and employment status were used to generate an SES parameter. Participants were categorized into quartiles based on household income level, namely “lowest”, “lower-middle”, “upper-middle”, and “highest”. Educational qualifications were classified per ISCED-2011 into three groups, including “lower secondary education or below”, “upper secondary education”, and “tertiary education”. We regrouped participants into two groups: employed (including employed or self-employed, retired) and unemployed (including unemployed, permanently sick or disabled, homemakers).

In the KLoSA cohort, household income, educational qualifications, and employment status were used to generate an SES parameter. Participants were categorized into quartiles based on household income level, namely “lowest”, “lower-middle”, “upper-middle”, and “highest”. Educational qualifications were classified into three groups, including “primary school or below”, “secondary education”, and “tertiary education”. The employment variable was coded as a binary indicator, with a value of 0 representing an unemployed participant and a value of 1 representing an employed participant.

Latent class analyses were performed for different potential classification numbers to select a reasonable model. The convergence criterion for parameter estimation was set as the maximum absolute deviation of parameter estimates in two consecutive iterations being 1×10^-^¹⁰, meaning that iterations would terminate when the difference between parameter estimates in two consecutive iterations was less than 1×10^-^¹⁰. A maximum of 3000 iterations was allowed to avoid local optimal solutions. The Akaike information criterion (AIC), Bayesian information criterion (BIC), and likelihood ratio statistic G^2^ were used for model selection. The average posterior probability reflects the uncertainty of the posterior classification and is also used for model selection, with a value of 0.7 or higher indicating acceptable uncertainty. The item response probability, a type of posterior probability, is used to define latent categories.

Since the model with six latent classes failed to converge, we only report information for models with five or fewer latent classes. As shown in these figures, the G^2^ statistic, AIC, and BIC continued to decrease as the number of latent classes increased. However, the decline plateaued after the three-latent-class solution.

We also examined the average posterior probabilities to facilitate model selection. The table below shows the average posterior probabilities, the prevalence of latent classes, and the item response probabilities in models with three to five latent classes. In the UKB and SHARE cohorts, the three-class latent model demonstrated an average posterior probability exceeding 0.7, whereas both the four-class and five-class models yielded average posterior probabilities below this threshold. For the KLoSA cohort, although the AIC and BIC values were the lowest among the five potential classes and the posterior probability for one class was close to 0.7, the three-class model was selected as the optimal solution. This decision was made to maintain consistency in the definition of SES across all three cohorts and was based on the assessment criteria for posterior classification uncertainty.

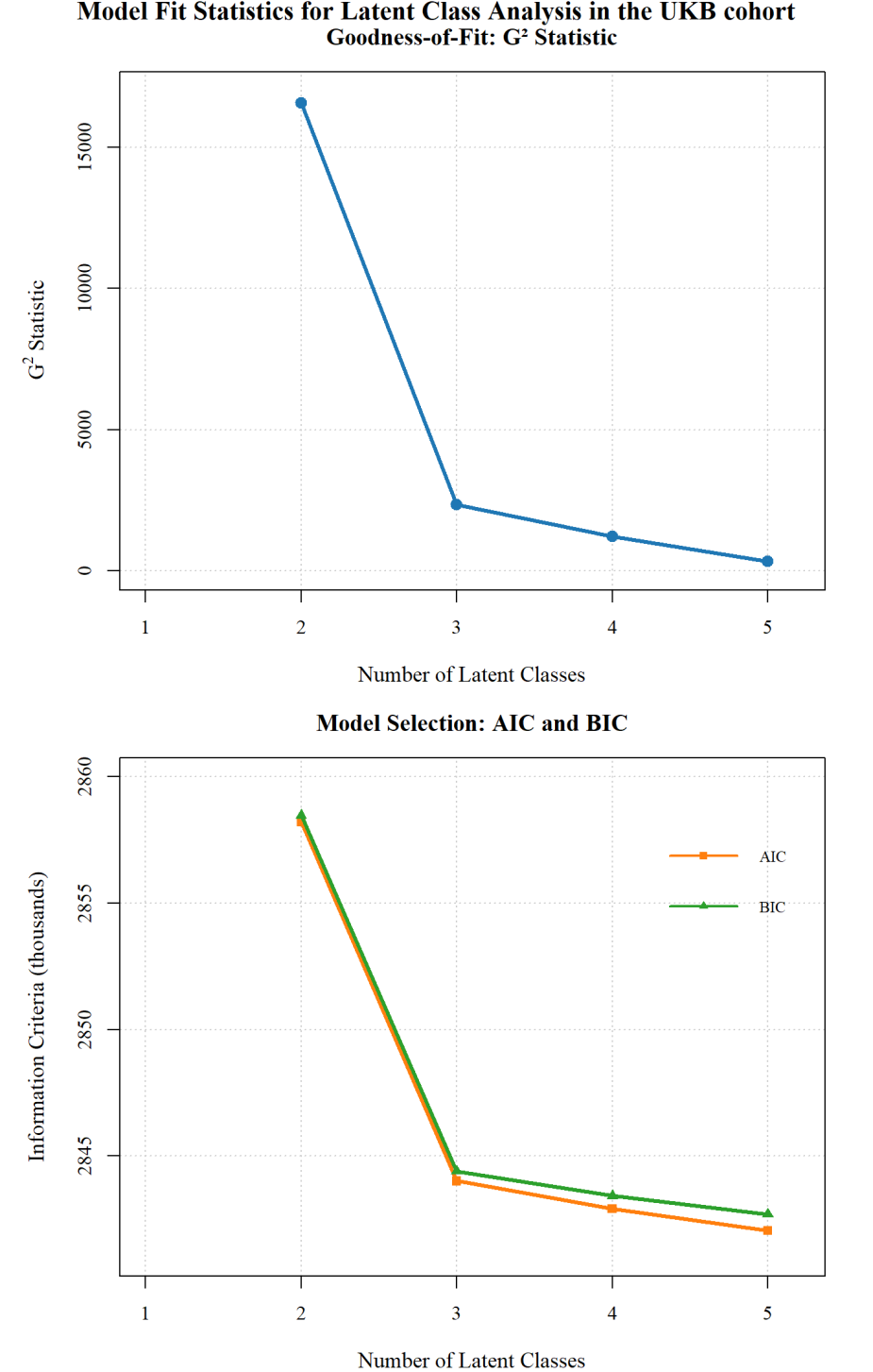

Figure S1. Model fit statistics for latent class analysis in the UKB cohort.

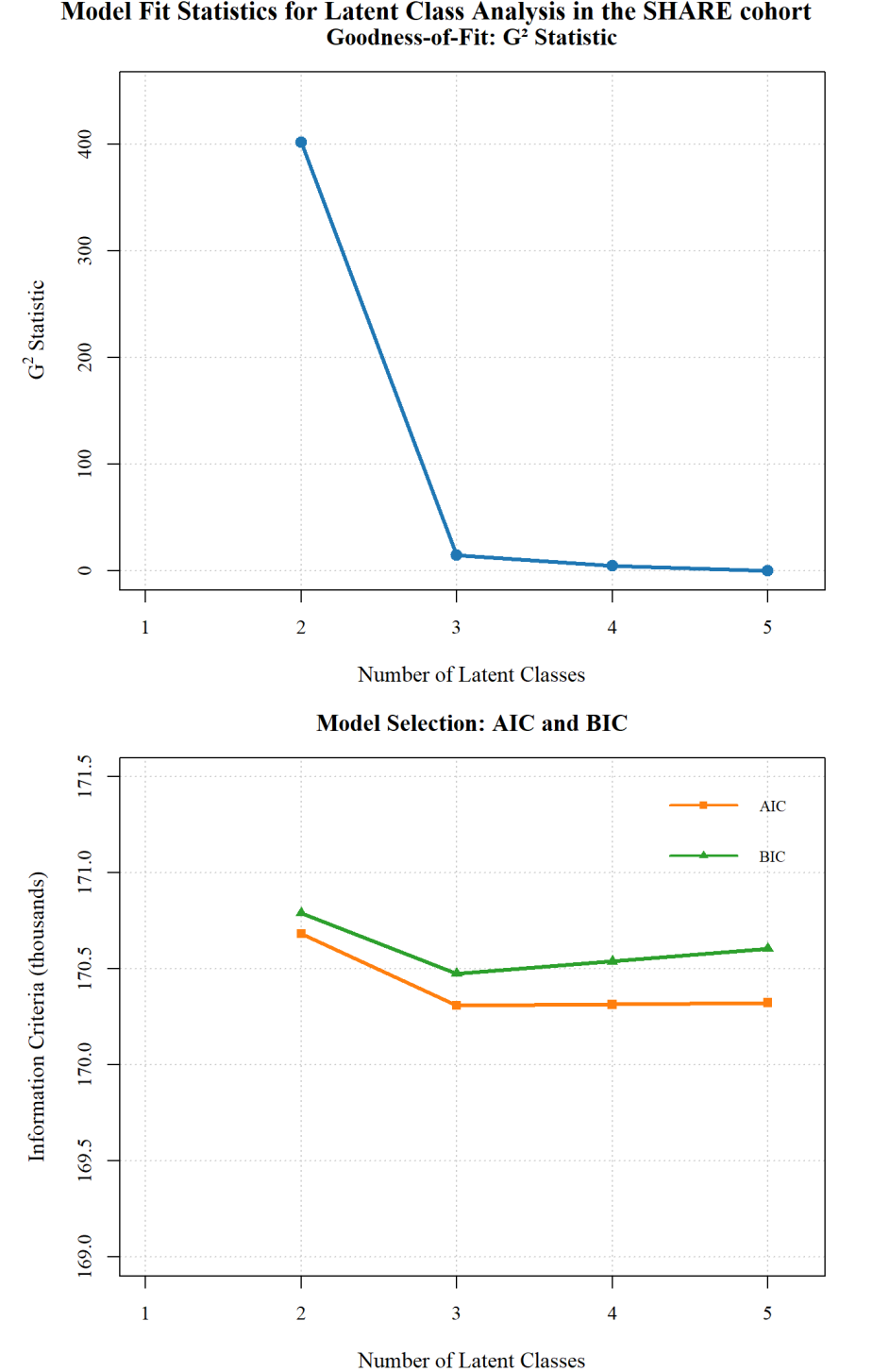

Figure S2. Model fit statistics for latent class analysis in the SHARE cohort.

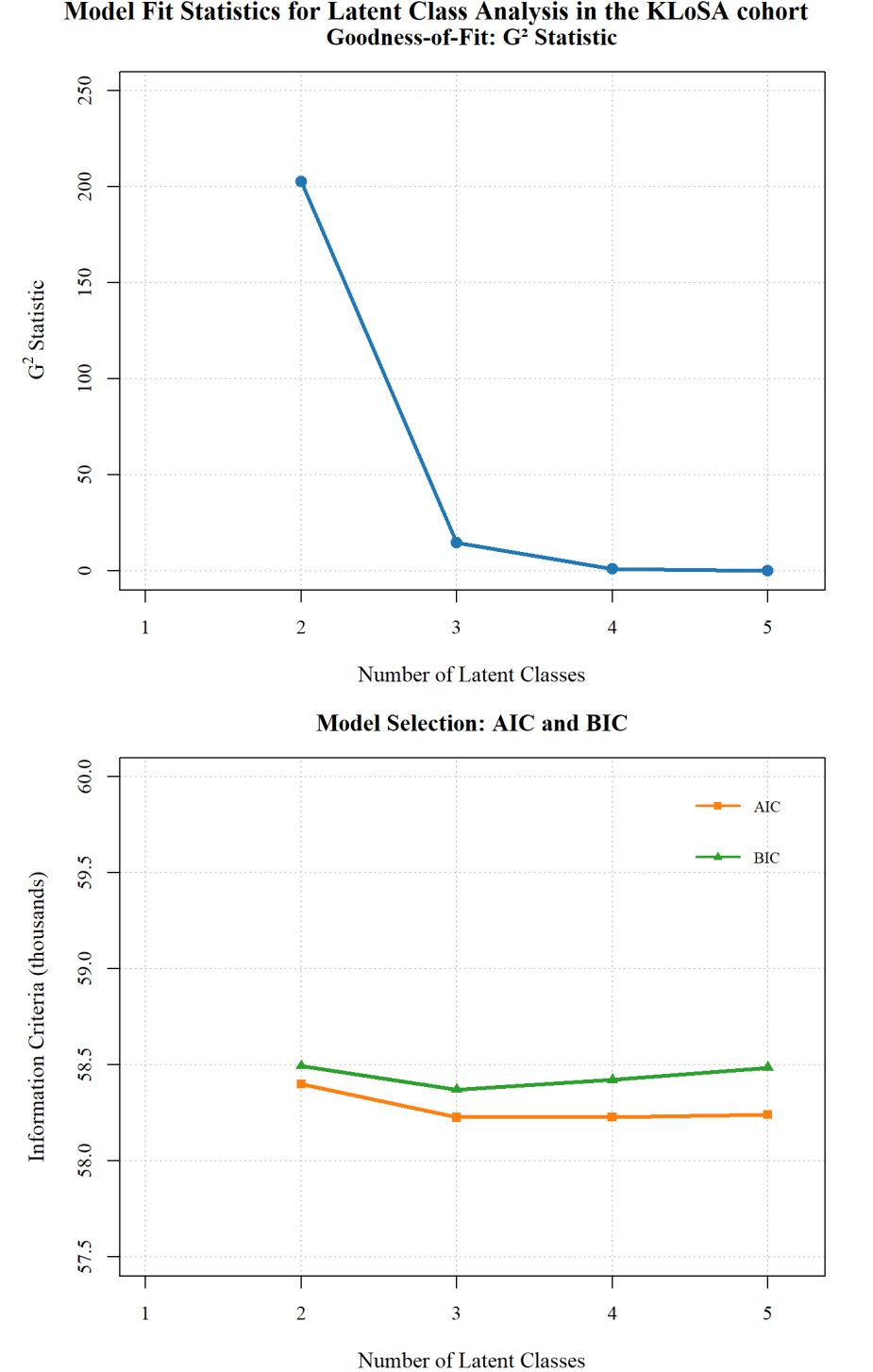

Figure S3. Model fit statistics for latent class analysis in the KLoSA cohort.

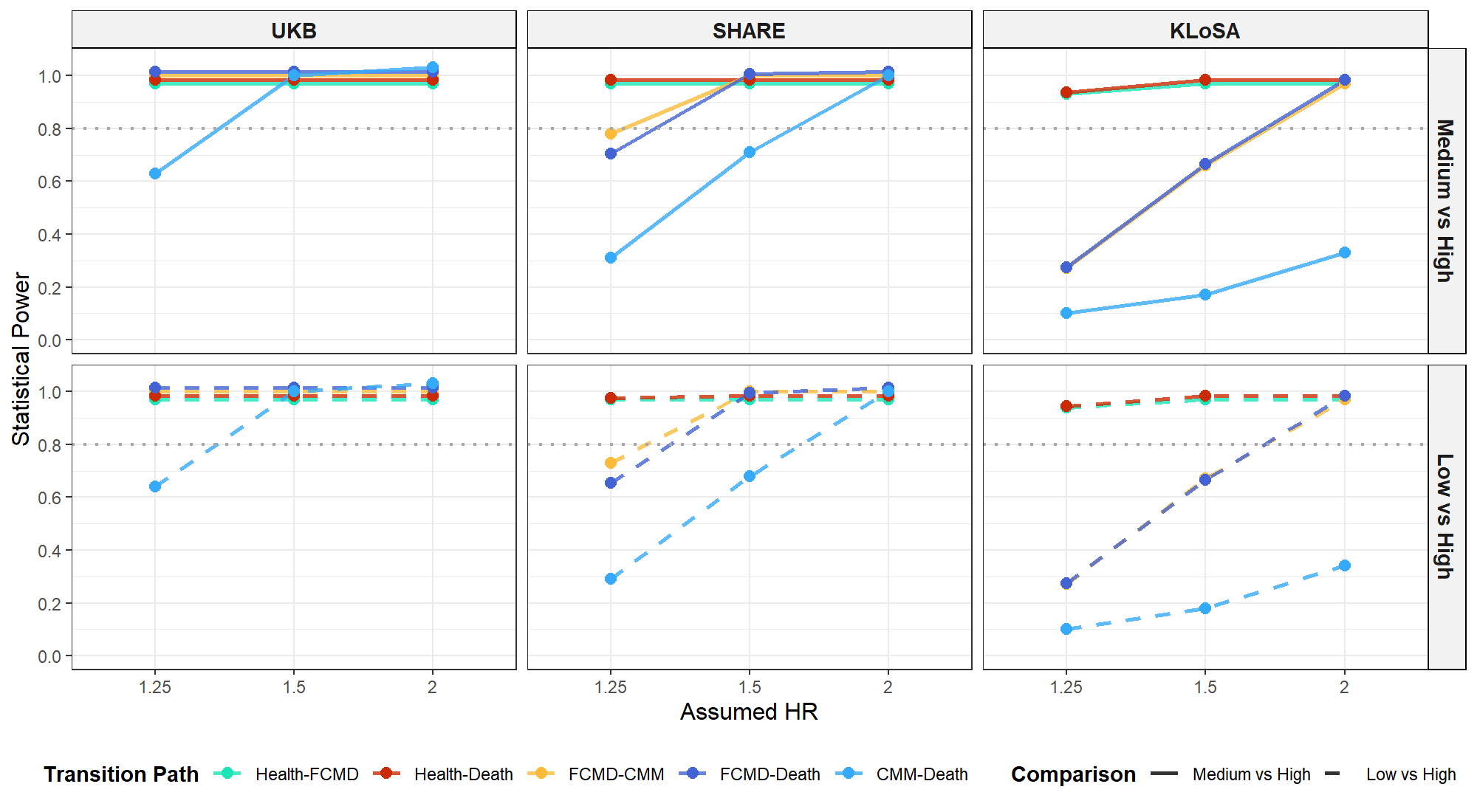

Figure S4. Power Curve for multistate transitions across three cohorts under varying assumed hazard ratios. SES, socioeconomic status; HR, hazard ratio; FCMD, first-onset cardiometabolic disease; CMM, cardiometabolic multimorbidity

Table S1. Mean posterior probabilities and item-response probabilities in models with three to five latent classes in the UKB cohort.

| **Item** | **Latent class1** | **Latent class 2** | **Latent class 3** | **Latent class 4** | **Latent class 5** |
| --- | --- | --- | --- | --- | --- |
| **Three-latent-class solution** | | | | | |
| **Mean PP** | 0.77 | 0.80 | 0.93 |  |  |
| **Income** |  |  |  |  |  |
| <£18000 | 0.00 | 0.72 | 0.00 | NA | NA |
| £18000-30999 | 0.41 | 0.22 | 0.00 | NA | NA |
| £31000-51999 | 0.42 | 0.04 | 0.24 | NA | NA |
| £52000-100000 | 0.17 | 0.01 | 0.54 | NA | NA |
| >£100000 | 0.00 | 0.01 | 0.23 | NA | NA |
| **Qualifications** |  |  |  |  |  |
| College or university degree | 0.30 | 0.15 | 0.72 | NA | NA |
| A/AS levels or equivalent | 0.13 | 0.08 | 0.13 | NA | NA |
| O/GCSEs level or equivalent | 0.27 | 0.21 | 0.09 | NA | NA |
| CSEs or equivalent | 0.07 | 0.06 | 0.01 | NA | NA |
| NVQ/HND/HNC or equivalent | 0.08 | 0.08 | 0.02 | NA | NA |
| Other professional qualifications | 0.06 | 0.05 | 0.03 | NA | NA |
| Less than high school | 0.08 | 0.36 | 0.00 | NA | NA |
| **Employment** |  |  |  |  |  |
| Employed | 0.98 | 0.84 | 0.96 | NA | NA |
| Unemployed | 0.02 | 0.16 | 0.04 | NA | NA |
| **Four-latent-class solution** | | | | | |
| **Mean PP** | 0.68 | 0.66 | 0.78 | 0.72 |  |
| **Income** |  |  |  |  |  |
| <£18000 | 0.91 | 0.01 | 0.00 | 0.02 | NA |
| £18000-30999 | 0.05 | 0.23 | 0.18 | 0.60 | NA |
| £31000-51999 | 0.01 | 0.01 | 0.51 | 0.31 | NA |
| £52000-100000 | 0.02 | 0.43 | 0.31 | 0.07 | NA |
| >£100000 | 0.01 | 0.31 | 0.00 | 0.00 | NA |
| **Qualifications** |  |  |  |  |  |
| College or university degree | 0.15 | 0.75 | 0.52 | 0.01 | NA |
| A/AS levels or equivalent | 0.08 | 0.11 | 0.15 | 0.10 | NA |
| O/GCSEs level or equivalent | 0.21 | 0.08 | 0.19 | 0.34 | NA |
| CSEs or equivalent | 0.07 | 0.01 | 0.05 | 0.09 | NA |
| NVQ/HND/HNC or equivalent | 0.08 | 0.02 | 0.05 | 0.11 | NA |
| Other professional qualifications | 0.05 | 0.03 | 0.04 | 0.08 | NA |
| Less than high school | 0.36 | 0.00 | 0.00 | 0.27 | NA |
| **Employment** |  |  |  |  |  |
| Employed | 0.83 | 0.95 | 0.97 | 0.95 | NA |
| Unemployed | 0.17 | 0.05 | 0.03 | 0.05 | NA |
| **Five-latent-class solution** | | | | | |
| **Mean PP** | 0.64 | 0.64 | 0.80 | 0.61 | 0.69 |
| **Income** |  |  |  |  |  |
| <£18000 | 0.00 | 0.76 | 0.00 | 0.34 | 0.00 |
| £18000-30999 | 0.39 | 0.15 | 0.10 | 0.41 | 0.18 |
| £31000-51999 | 0.44 | 0.05 | 0.16 | 0.19 | 0.38 |
| £52000-100000 | 0.18 | 0.03 | 0.29 | 0.05 | 0.44 |
| >£100000 | 0.00 | 0.01 | 0.45 | 0.01 | 0 |
| **Qualifications** |  |  |  |  |  |
| College or university degree | 0.09 | 0.15 | 0.75 | 0.16 | 0.69 |
| A/AS levels or equivalent | 0.17 | 0.09 | 0.11 | 0.06 | 0.14 |
| O/GCSEs level or equivalent | 0.41 | 0.21 | 0.08 | 0.23 | 0.11 |
| CSEs or equivalent | 0.16 | 0.09 | 0.01 | 0.00 | 0.00 |
| NVQ/HND/HNC or equivalent | 0.09 | 0.07 | 0.02 | 0.11 | 0.03 |
| Other professional qualifications | 0.06 | 0.03 | 0.03 | 0.10 | 0.03 |
| Less than high school | 0.02 | 0.36 | 0.00 | 0.36 | 0.00 |
| **Employment** |  |  |  |  |  |
| Employed | 0.96 | 0.76 | 0.94 | 1.00 | 0.98 |
| Unemployed | 0.04 | 0.24 | 0.06 | 0.00 | 0.02 |

Notes: mean PP, mean posterior probability; NA, not available.

Table S2. Mean posterior probabilities and item-response probabilities in models with three to five latent classes in the SHARE cohort.

| **Item** | **Latent class1** | **Latent class 2** | **Latent class 3** | **Latent class 4** | **Latent class 5** |
| --- | --- | --- | --- | --- | --- |
| **Three-latent-class solution** | | | | | |
| **Mean PP** | 0.75 | 0.73 | 0.85 |  |  |
| **Income** |  |  |  |  |  |
| lowest | 0.66 | 0.09 | 0.08 | NA | NA |
| lower-middle | 0.21 | 0.34 | 0.14 | NA | NA |
| upper-middle | 0.04 | 0.36 | 0.30 | NA | NA |
| highest | 0.09 | 0.20 | 0.47 | NA | NA |
| **Qualifications** |  |  |  |  |  |
| lower secondary education or below | 0.82 | 0.65 | 0.00 | NA | NA |
| upper secondary education | 0.14 | 0.33 | 0.40 | NA | NA |
| tertiary education | 0.04 | 0.02 | 0.60 | NA | NA |
| **Employment** |  |  |  |  |  |
| Employed | 0.57 | 0.80 | 0.92 | NA | NA |
| Unemployed | 0.43 | 0.20 | 0.08 | NA | NA |
| **Four-latent-class solution** | | | | | |
| **Mean PP** | 0.62 | 0.84 | 0.58 | 0.6 |  |
| **Income** |  |  |  |  |  |
| lowest | 0.77 | 0.08 | 0.30 | 0.04 | NA |
| lower-middle | 0.11 | 0.14 | 0.42 | 0.28 | NA |
| upper-middle | 0.10 | 0.31 | 0.00 | 0.55 | NA |
| highest | 0.01 | 0.47 | 0.28 | 0.12 | NA |
| **Qualifications** |  |  |  |  |  |
| lower secondary education or below | 0.85 | 0.00 | 0.70 | 0.65 | NA |
| upper secondary education | 0.12 | 0.41 | 0.25 | 0.35 | NA |
| tertiary education | 0.03 | 0.59 | 0.05 | 0.00 | NA |
| **Employment** |  |  |  |  |  |
| Employed | 0.52 | 0.92 | 0.72 | 0.82 | NA |
| Unemployed | 0.48 | 0.08 | 0.28 | 0.18 | NA |
| **Five-latent-class solution** | | | | | |
| **Mean PP** | 0.57 | 0.58 | 0.68 | 0.61 | 0.70 |
| **Income** |  |  |  |  |  |
| lowest | 0.59 | 0.01 | 0.37 | 0.66 | 0.06 |
| lower-middle | 0.08 | 0.51 | 0.01 | 0.31 | 0.14 |
| upper-middle | 0.16 | 0.28 | 0.40 | 0.03 | 0.30 |
| highest | 0.18 | 0.20 | 0.22 | 0.00 | 0.49 |
| **Qualifications** |  |  |  |  |  |
| lower secondary education or below | 0.75 | 0.62 | 0.55 | 0.89 | 0.07 |
| upper secondary education | 0.25 | 0.38 | 0.43 | 0.02 | 0.33 |
| tertiary education | 0.00 | 0.01 | 0.02 | 0.09 | 0.60 |
| **Employment** |  |  |  |  |  |
| Employed | 0.21 | 0.79 | 0.96 | 0.66 | 0.92 |
| Unemployed | 0.80 | 0.21 | 0.04 | 0.34 | 0.08 |

Notes: mean PP, mean posterior probability; NA, not available.

Table S3. Mean posterior probabilities and item-response probabilities in models with three to five latent classes in the KLoSA cohort.

| **Item** | **Latent class1** | **Latent class 2** | **Latent class 3** | **Latent class 4** | **Latent class 5** |
| --- | --- | --- | --- | --- | --- |
| **Three-latent-class solution** | | | | | |
| **Mean PP** | 0.68 | 0.75 | 0.74 |  |  |
| **Income** |  |  |  |  |  |
| lowest | 0.37 | 0.14 | 0.03 | NA | NA |
| lower-middle | 0.22 | 0.20 | 0.09 | NA | NA |
| upper-middle | 0.13 | 0.34 | 0.08 | NA | NA |
| highest | 0.27 | 0.31 | 0.79 | NA | NA |
| **Qualifications** |  |  |  |  |  |
| lower secondary education or below | 0.69 | 0.59 | 0.00 | NA | NA |
| upper secondary education | 0.25 | 0.41 | 0.69 | NA | NA |
| tertiary education | 0.07 | 0.00 | 0.31 | NA | NA |
| **Employment** |  |  |  |  |  |
| Employed | 0.02 | 0.48 | 0.69 | NA | NA |
| Unemployed | 0.98 | 0.52 | 0.31 | NA | NA |
| **Four-latent-class solution** | | | | | |
| **Mean PP** | 0.69 | 0.89 | 0.72 | 0.77 |  |
| **Income** |  |  |  |  |  |
| lowest | 0.36 | 0.01 | 0.38 | 0.01 | NA |
| lower-middle | 0.22 | 0.24 | 0.05 | 0.08 | NA |
| upper-middle | 0.14 | 0.49 | 0.02 | 0.08 | NA |
| highest | 0.28 | 0.26 | 0.55 | 0.83 | NA |
| **Qualifications** |  |  |  |  |  |
| lower secondary education or below | 0.74 | 0.52 | 0.15 | 0.03 | NA |
| upper secondary education | 0.19 | 0.48 | 0.85 | 0.64 | NA |
| tertiary education | 0.07 | 0.00 | 0.00 | 0.33 | NA |
| **Employment** |  |  |  |  |  |
| Employed | 0.13 | 0.49 | 0.36 | 0.72 | NA |
| Unemployed | 0.87 | 0.51 | 0.64 | 0.28 | NA |
| **Five-latent-class solution** | | | | | |
| **Mean PP** | 0.68 | 0.48 | 0.63 | 0.32 | 0.52 |
| **Income** |  |  |  |  |  |
| lowest | 0.02 | 0.25 | 0.08 | 0.02 | 0.37 |
| lower-middle | 0.11 | 0.12 | 0.26 | 0.06 | 0.23 |
| upper-middle | 0.03 | 0.14 | 0.45 | 0.15 | 0.15 |
| highest | 0.85 | 0.48 | 0.21 | 0.77 | 0.25 |
| **Qualifications** |  |  |  |  |  |
| lower secondary education or below | 0.09 | 0.29 | 0.59 | 0.01 | 0.81 |
| upper secondary education | 0.49 | 0.68 | 0.40 | 0.78 | 0.12 |
| tertiary education | 0.43 | 0.02 | 0.01 | 0.21 | 0.08 |
| **Employment** |  |  |  |  |  |
| Employed | 0.66 | 0.24 | 0.51 | 0.88 | 0.08 |
| Unemployed | 0.34 | 0.76 | 0.49 | 0.12 | 0.92 |

Notes: mean PP, mean posterior probability; NA, not available.

Table S4. Definitions of the three cardiometabolic diseases.

|  | **UKB** | | **SHARE** | **KLoSA** | |
| --- | --- | --- | --- | --- | --- |
|  | **Fields** | **Codes** | **Codes** | **Waves** | **Codes** |
| **Type 2 diabetes** | 41270 | E11 | r2diabe | w2 | w02C011, w02C012y, w02C012m |
|  | 20002 | 1223 | r4diabe | w3 | w03C011, w03C012y, w03C012m |
|  | 2443 | 1 | r5diabe | w4 | w04C011, w04C012y, w04C012m |
|  | 6153 | 3 | r6diabe | w5 | w05C011, w05C012y, w05C012m |
|  | 6157 | 3 | r7diabe | w6 | w06C011, w06C012y, w06C012m |
|  | 2976 | - | r8diabe | w7 | w07C011, w07C012y, w07C012m |
|  | - | - | r9diabe | w8 | w08C011, w08C012y, w08C012m |
|  | - | - | - | w9 | w09C011, w09C012y, w09C012m |
| **Coronary artery disease** | 41270 | I20, I21, I22, I23, I24, I25 | r2hearte | w2 | w02C033, w02C034y, w02C034m |
|  | 20002 | 1066, 1074, 1075 | r4hearte | w3 | w03C033, w03C034y, w03C034m |
|  | 6150 | 1,2 | r5hearte | w4 | w04C033, w04C034y, w04C034m |
|  | 3627 | - | r6hearte | w5 | w05C033, w05C034y, w05C034m |
|  | 3894 | - | r7hearte | w6 | w06C033, w06C034y, w06C034m |
|  | - | - | r8hearte | w7 | w07C033, w07C034y, w07C034m |
|  | - | - | r9hearte | w8 | w08C033, w08C034y, w08C034m |
|  | - | - | - | w9 | w09C033, w09C034y, w09C034m |
| **Stroke** | 41270 | I60, I61, I62, I63, I64, I69 | r2stroke | w2 | w02C038, w02C039y, w02C039m |
|  | 20002 | 1081, 1086, 1491, 1583 | r4stroke | w3 | w03C038, w03C039y, w03C039m |
|  | 6150 | 3 | r5stroke | w4 | w04C038, w04C039y, w04C039m |
|  | 4056 | - | r6stroke | w5 | w05C038, w05C039y, w05C039m |
|  | 42006 | - | r7stroke | w6 | w06C038, w06C039y, w06C039m |
|  | 42008 | - | r8stroke | w7 | w07C038, w07C039y, w07C039m |
|  | 42010 | - | r9stroke | w8 | w08C038, w08C039y, w08C039m |
|  | 42012 | - | - | w9 | w09C038, w09C039y, w09C039m |

Table S5. Definitions and data-fields of covariates.

|  | UKB | | SHARE | | KLoSA | |
| --- | --- | --- | --- | --- | --- | --- |
| Phenotype | Data fields | Field names | Data fields | Field names | Data fields | Field names |
| **Socioeconomic indicators** | | | | | | |
| income | 738 | Average total household income before tax | hh1itot | w1 Incm: HHold Total Income (before-tax) | r1itothhinc | w1 income:r total household income, direct quest Cont |
| education | 6138 | Qualifications | raedisced | r education by isced code | raeduc_k | R Education category |
| employment | 6142 | Current employment status | r1lbrf_s | w1 R labor force status | r1work | w1 R working for pay |
| **Covariates** | | | | | | |
| Chronological age | 21022 | Age at recruitment | r1agey | w1 r age (years) at ivw | r1agey | w1 R age in years at ivw |
| Sex | 31 | Sex | ragender | r gender | ragender | R gender |
| Ethnicity | 21000 | Ethnic background | - | - | - | - |
| BMI | 21001 | Body mass index (BMI) | r1bmi | w1 R Body Mass Index=kg/m2 | r1bmi | w1 R body mass index=kg/m2 |
| Smoke status | 20116 | Smoking status | r1smokev | w1 R smoke ever | r1smokev | w1 R ever smoked |
| Drink status | 1558 | Alcohol drinker status | r1drinkx | w1 R Frequency of drinking | r1drinkx | w1 R frequency of drinking last year |
| Physical activity | 22032 | IPAQ activity group | r1mdactx | w1 R Freq moderate phys activity | r1vigactf_k | w1 R freq exercise(times/per week) |
| Sleep patterns | 1160 | Sleep duration | r1sleep | w1 EURO-D: R Sleep (0,1) | r1sleeprl | w1 R CESD sleep was restless |
|  | 1180 | Morning/evening person (chronotype) | - | - | - | - |
|  | 1200 | Sleeplessness / insomnia | - | - | - | - |
|  | 1210 | Snoring | - | - | - | - |
|  | 1220 | Daytime dozing / sleeping | - | - | - | - |
| Blood pressure | 4080 | Systolic blood pressure, automated reading | r1hibpe | w1 R ever had high blood pressure | r1hibpe | w1 R ever had high BP |
|  | 4079 | diastolic blood pressure, automated reading | - | - | - | - |

Notes: BMI, Body mass index.

Table S6. Summary information of all included participants in the UKB cohort under different statuses.

| **Characteristics** | **All*(*N*=387,665)** | **FCMD*(*N*=53,334)** | **CMM*(*N*=7,566)** | **Death*(*N*=24,121)** |
| --- | --- | --- | --- | --- |
| **Age (years)** | 55.8 (8.1) | 59.5 (7.2) | 60.9 (6.6) | 61.1 (6.6) |
| **Sex (%)** |  |  |  |  |
| Female | 209,877 (54.1) | 21,082 (39.5) | 2,531 (33.5) | 10,078 (41.8) |
| Male | 177,788 (45.9) | 32,252 (60.5) | 5,035 (66.5) | 14,043 (58.2) |
| **SES (%)** |  |  |  |  |
| High | 78,920 (20.4) | 6,476 (12.1) | 633 (8.37) | 10,916 (45.3) |
| Medium | 206,618 (53.3) | 26,126 (49.0) | 3,359 (44.4) | 2,477 (10.3) |
| Low | 102,127 (26.3) | 20,732 (38.9) | 3,574 (47.2) | 10,728 (44.5) |
| **Income (%)** |  |  |  |  |
| < £18000 | 82,112 (21.2) | 16,943 (31.8) | 2,936 (38.8) | 8,959 (37.1) |
| £18000-30999 | 97,850 (25.2) | 15,060 (28.2) | 2,188 (28.9) | 7,009 (29.1) |
| £31000-51999 | 103,652 (26.7) | 12,127 (22.7) | 1,513 (20.0) | 4,800 (19.9) |
| £52000-100000 | 82,133 (21.2) | 7,500 (14.1) | 760 (10.0) | 2,699 (11.2) |
| >£100000 | 21,918 (5.7) | 1,704 (3.2) | 169 (2.2) | 654 (2.7) |
| **Qualifications (%)** |  |  |  |  |
| College or university degree | 139,358 (35.9) | 14,522 (27.2) | 1,815 (24.0) | 6,674 (27.7) |
| A/AS levels or equivalent | 45,892 (11.8) | 5,322 (10.0) | 647 (8.5) | 2,317 (9.6) |
| O/GCSEs level or equivalent | 83,058 (21.4) | 11,112 (20.8) | 1,535 (20.3) | 4,840 (20.1) |
| CSEs or equivalent | 21,340 (5.5) | 2,596 (4.9) | 327 (4.3) | 878 (3.6) |
| NVQ/HND/HNC or equivalent | 25,021 (6.5) | 4,609 (8.6) | 693 (9.2) | 1,936 (8.0) |
| Other professional qualifications | 19,420 (5.1) | 3,114 (5.9) | 426 (5.6) | 1,396 (5.8) |
| Less than high school | 53,576 (13.8) | 12,059 (22.6) | 2,123 (28.1) | 6,080 (25.2) |
| **Employment (%)** |  |  |  |  |
| Employed | 362,305 (93.5) | 48,935 (91.8) | 6,773 (89.5) | 21,935 (90.9) |
| Unemployed | 25,360 (6.5) | 4,399 (8.2) | 793 (10.5) | 2,186 (9.1) |
| **Sleep mode (%)** |  |  |  |  |
| 0 | 558 (0.1) | 195 (0.4) | 38 (0.5) | 62 (0.3) |
| 1 | 8,328 (2.2) | 1,887 (3.5) | 339 (4.5) | 723 (3.0) |
| 2 | 43,273 (11.2) | 7,767 (14.6) | 1,257 (16.6) | 3,212 (13.3) |
| 3 | 108,933 (28.1) | 16,610 (31.1) | 2,425 (32.1) | 7,207 (29.9) |
| 4 | 146,256 (37.7) | 18,513 (34.7) | 2,555 (33.8) | 8,783 (36.4) |
| 5 | 80,317 (20.7) | 8,362 (15.7) | 952 (12.6) | 4,134 (17.1) |
| **Physical activity (%)** |  |  |  |  |
| 0 | 72,238 (18.6) | 11,569 (21.7) | 1,823 (24.1) | 5,180 (21.5) |
| 1 | 158,847 (41.0) | 21,442 (40.2) | 2,987 (39.5) | 9,876 (40.9) |
| 2 | 156,580 (40.4) | 20,323 (38.1) | 2,756 (36.4) | 9,065 (37.6) |
| **Alcohol consumption (%)** |  |  |  |  |
| Less than once per week | 110,800 (28.6) | 18,343 (34.4) | 2,925 (38.7) | 7,536 (31.2) |
| More than once per week | 276,865 (71.4) | 34,991 (65.6) | 4,641 (61.3) | 16,585 (68.8) |
| **Smoke status (%)** |  |  |  |  |
| Never | 215,353 (55.6) | 24,817 (46.5) | 3,109 (41.1) | 9,840 (40.8) |
| Former or current smoker | 172,312 (44.4) | 28,517 (53.5) | 4,457 (58.9) | 14,281 (59.2) |
| **Deprive index** | -1.4 (3.0) | -1.0 (3.2) | -0.6 (3.4) | -1.0 (3.3) |
| **BMI** | 27.2 (4.7) | 29.3 (5.3) | 30.5 (5.5) | 27.9 (5.2) |
| **Diastolic BP** | 82.4 (10.1) | 84.3 (10.2) | 84.2 (10.5) | 83.3 (10.5) |
| **Systolic BP** | 137 (18.6) | 144 (18.5) | 145 (19.0) | 143 (19.6) |
| **Race (%)** |  |  |  |  |
| Non-White | 17,777 (4.6) | 3,305 (6.2) | 623 (8.2) | 667 (2.8) |
| White | 369,888 (95.4) | 50,029 (93.8) | 6,943 (91.8) | 23,454 (97.2) |

Notes: Data were presented as frequency (%) and mean (standard deviation, SD); FCMD, first-onset cardiometabolic disease; CMM, cardiometabolic multimorbidity; SES, socioeconomic status; BMI, body mass index; BP, blood pressure; the asterisk (*) indicates the diagnosis or occurrence during the follow-up.

Table S7. Summary information of all included participants in the SHARE cohort under different statuses.

| **Characteristics** | **All*(*N*=22,505)** | **FCMD*(*N*=4,706)** | **CMM*(*N*=1,025)** | **Death*(*N*=4,087)** |
| --- | --- | --- | --- | --- |
| **Age (years)** | 62.5 (10.3) | 64.8 (9.5) | 65.6 (9.0) | 71.6 (10.3) |
| **Sex (%)** |  |  |  |  |
| Female | 13,066 (58.1) | 2,507 (53.3) | 501 (48.9) | 2,097 (51.3) |
| Male | 9,439 (41.9) | 2,199 (46.7) | 524 (51.1) | 1,990 (48.7) |
| **SES (%)** |  |  |  |  |
| High | 6,249 (27.8) | 1,115 (23.7) | 215 (21.0) | 667 (16.3) |
| Medium | 10,459 (46.5) | 2,171 (46.1) | 474 (46.2) | 1,945 (47.6) |
| Low | 5,797 (25.8) | 1,420 (30.2) | 336 (32.8) | 1,475 (36.1) |
| **Income (%)** |  |  |  |  |
| lowest | 5,313 (23.6) | 1,278 (27.2) | 301 (29.4) | 1,449 (35.5) |
| lower-middle | 5,412 (24.0) | 1,270 (27.0) | 302 (29.5) | 1,135 (27.8) |
| upper-middle | 5,745 (25.5) | 1,151 (24.5) | 241 (23.5) | 882 (21.6) |
| highest | 6,035 (26.8) | 1,007 (21.4) | 181 (17.7) | 621 (15.2) |
| **Qualifications (%)** |  |  |  |  |
| lower secondary education or below | 10,969 (48.7) | 2,601 (55.3) | 593 (57.9) | 2,592 (63.4) |
| upper secondary education | 7,017 (31.2) | 1,286 (27.3) | 269 (26.2) | 997 (24.4) |
| tertiary education | 4,519 (20.1) | 819 (17.4) | 163 (15.9) | 498 (12.2) |
| **Employment (%)** |  |  |  |  |
| Employed | 17,381 (77.2) | 3,643 (77.4) | 806 (78.6) | 3,246 (79.4) |
| Unemployed | 5,124 (22.8) | 1,063 (22.6) | 219 (21.4) | 841 (20.6) |
| **Sleep (%)** |  |  |  |  |
| No | 15,807 (70.8) | 3,176 (67.9) | 690 (67.6) | 2,835 (70.6) |
| Yes | 6,516 (29.2) | 1,502 (32.1) | 331 (32.4) | 1,179 (29.4) |
| **High blood pressure (%)** |  |  |  |  |
| No | 16,506 (73.3) | 3,047 (64.7) | 610 (59.5) | 2,677 (65.5) |
| Yes | 5,999 (26.7) | 1,659 (35.3) | 415 (40.5) | 1,410 (34.5) |
| **Physical activity (%)** |  |  |  |  |
| > 1 per week | 16,260 (72.3) | 3,314 (70.4) | 688 (67.1) | 2,614 (64.0) |
| 1 per week | 2,924 (13.0) | 623 (13.2) | 135 (13.2) | 484 (11.8) |
| 1-3 per month | 1,124 (5.0) | 254 (5.5) | 69 (6.7) | 211 (5.2) |
| hardly ever or never | 2,196 (9.7) | 515 (10.9) | 133 (13.0) | 778 (19.0) |
| **Alcohol consumption (%)** |  |  |  |  |
| Less than once per week | 11,037 (49.1) | 2,415 (51.3) | 567 (55.3) | 2,282 (55.9) |
| More than once per week | 11,462 (50.9) | 2,290 (48.7) | 458 (44.7) | 1,801 (44.1) |
| **Smoke status (%)** |  |  |  |  |
| Never | 12,036 (53.5) | 2,599 (55.2) | 555 (54.1) | 2,158 (52.8) |
| Former or current smoker | 10,467 (46.5) | 2,106 (44.8) | 470 (45.9) | 1,928 (47.2) |
| **BMI** | 26.1 (4.1) | 27.1 (4.4) | 27.9 (4.6) | 26.0 (4.3) |

Notes: Data were presented as frequency (%) and mean (standard deviation, SD); FCMD, first-onset cardiometabolic disease; CMM, cardiometabolic multimorbidity; SES, socioeconomic status; BMI, body mass index; the asterisk (*) indicates the diagnosis or occurrence during the follow-up.

Table S8. Summary information of all included participants in the KLoSA cohort under different statuses.

| **Characteristics** | **All*(*N*=8,357)** | **FCMD*(*N*=1,714)** | **CMM*(*N*=240)** | **Death*(*N*=1,896)** |
| --- | --- | --- | --- | --- |
| **Age (years)** | 60.7 (11.2) | 62.1 (9.6) | 62.9 (8.8) | 70.3 (11.4) |
| **Sex (%)** |  |  |  |  |
| Female | 3,818 (57.5) | 945 (55.1) | 123 (51.2) | 980 (51.7) |
| Male | 2,825 (42.5) | 769 (44.9) | 117 (48.8) | 916 (48.3) |
| **SES (%)** |  |  |  |  |
| High | 2,881 (34.5) | 473 (27.6) | 55 (22.9) | 292 (15.4) |
| Medium | 2,630 (31.5) | 603 (35.2) | 82 (34.2) | 567 (29.9) |
| Low | 2,846 (34.1) | 638 (37.2) | 103 (42.9) | 1,037 (54.7) |
| **Income (%)** |  |  |  |  |
| lowest | 1,461 (17.5) | 332 (19.4) | 51 (21.2) | 490 (25.8) |
| lower-middle | 1,426 (17.1) | 326 (19.0) | 49 (20.4) | 411 (21.7) |
| upper-middle | 1,654 (19.8) | 373 (21.8) | 59 (24.6) | 374 (19.7) |
| highest | 3,816 (45.7) | 683 (39.8) | 81 (33.8) | 621 (32.8) |
| **Qualifications (%)** |  |  |  |  |
| lower secondary education or below | 3,567 (42.7) | 807 (47.1) | 112 (46.7) | 1,253 (66.1) |
| upper secondary education | 3,790 (45.4) | 729 (42.5) | 106 (44.2) | 513 (27.1) |
| tertiary education | 1,000 (12.0) | 178 (10.4) | 22 (9.1) | 130 (6.9) |
| **Employment (%)** |  |  |  |  |
| Employed | 3,509 (42.0) | 682 (39.8) | 83 (34.6) | 485 (25.6) |
| Unemployed | 4,848 (58.0) | 1,032 (60.2) | 157 (65.4) | 1,411 (74.4) |
| **Sleep (%)** |  |  |  |  |
| No | 6,028 (72.1) | 1,199 (70.0) | 163 (67.9) | 1,182 (62.3) |
| Yes | 2,329 (27.9) | 515 (30.0) | 77 (32.1) | 714 (37.7) |
| **High blood pressure (%)** |  |  |  |  |
| No | 6,545 (78.3) | 1,199 (70.0) | 147 (61.3) | 1,355 (71.5) |
| Yes | 1,812 (21.7) | 515 (30.0) | 93 (38.8) | 541 (28.5) |
| **Physical activity (%)** |  |  |  |  |
| <= 1 per week | 5,176 (61.9) | 1,042 (60.8) | 148 (61.7) | 1,349 (71.1) |
| > 1 per week | 3,181 (38.1) | 672 (39.2) | 92 (38.3) | 547 (28.9) |
| **Alcohol consumption (%)** |  |  |  |  |
| Less than once per week | 6,451 (77.2) | 1,325 (77.3) | 188 (78.3) | 1,486 (78.4) |
| More than once per week | 1,906 (22.8) | 389 (22.7) | 52 (21.7) | 410 (21.6) |
| **Smoke status (%)** |  |  |  |  |
| Never | 5,990 (71.7) | 1,181 (68.9) | 152 (63.3) | 1,238 (65.3) |
| Former or current smoker | 2,367 (28.3) | 533 (31.1) | 88 (36.7) | 658 (34.7) |
| **BMI** | 23.1 (2.8) | 23.7 (2.8) | 24.2 (2.8) | 22.3 (3.1) |

Notes: Data were presented as frequency (%) and mean (standard deviation, SD); FCMD, first-onset cardiometabolic disease; CMM, cardiometabolic multimorbidity; SES, socioeconomic status; BMI, body mass index; the asterisk (*) indicates the diagnosis or occurrence during the follow-up.

Table S9. Cumulative incidence for the probability of CMM transitions across three latent SES classes for three cohorts.

| **Transitions** | **UKB** | **SHARE** | **KLoSA** |
| --- | --- | --- | --- |
|  | Incidence rate (%) | Incidence rate (%) | Incidence rate (%) |
| Health → FCMD (5 years) |  |  |  |
| High SES | 0.39 | 1.26 | 0.85 |
| Medium SES | 0.81 | 1.51 | 1.51 |
| Low SES | 1.95 | 2.36 | 2.25 |
| Health → FCMD (10 years) |  |  |  |
| High SES | 0.71 | 2.96 | 1.49 |
| Medium SES | 1.50 | 4.08 | 2.84 |
| Low SES | 3.46 | 4.90 | 3.47 |
| Health → FCMD (15 years) |  |  |  |
| High SES | 0.85 | 3.61 | 2.04 |
| Medium SES | 1.76 | 4.91 | 3.40 |
| Low SES | 3.95 | 6.41 | 3.95 |
| FCMD → CMM (5 years) |  |  |  |
| High SES | 9.42 | 16.55 | 10.68 |
| Medium SES | 12.00 | 18.11 | 10.19 |
| Low SES | 16.34 | 19.22 | 11.84 |
| FCMD → CMM (10 years) |  |  |  |
| High SES | 14.64 | 23.44 | 14.41 |
| Medium SES | 19.66 | 25.61 | 16.69 |
| Low SES | 28.22 | 29.43 | 18.69 |
| FCMD → CMM (15 years) |  |  |  |
| High SES | 16.66 | 26.81 | 21.77 |
| Medium SES | 23.10 | 29.11 | 23.68 |
| Low SES | 32.68 | 32.05 | 22.57 |
| Health → Death (5 years) |  |  |  |
| High SES | 0.63 | 2.53 | 1.82 |
| Medium SES | 0.98 | 5.23 | 4.91 |
| Low SES | 1.80 | 6.14 | 10.18 |
| Health → Death (10 years) |  |  |  |
| High SES | 1.66 | 5.30 | 5.14 |
| Medium SES | 2.59 | 10.27 | 12.19 |
| Low SES | 4.75 | 13.07 | 25.92 |
| Health → Death (15 years) |  |  |  |
| High SES | 2.50 | 7.59 | 9.20 |
| Medium SES | 4.00 | 13.59 | 20.92 |
| Low SES | 7.35 | 18.17 | 38.02 |
| FCMD → Death (5 years) |  |  |  |
| High SES | 7.76 | 8.23 | 5.20 |
| Medium SES | 9.04 | 11.19 | 6.52 |
| Low SES | 13.08 | 15.53 | 11.68 |
| FCMD → Death (10 years) |  |  |  |
| High SES | 11.08 | 14.16 | 11.30 |
| Medium SES | 13.67 | 20.66 | 14.38 |
| Low SES | 20.26 | 29.21 | 25.75 |
| FCMD → Death (15 years) |  |  |  |
| High SES | 13.05 | 18.76 | 13.70 |
| Medium SES | 15.93 | 28.01 | 17.88 |
| Low SES | 24.51 | 38.27 | 35.59 |
| CMM → Death (5 years) |  |  |  |
| High SES | 13.84 | 11.87 | 5.70 |
| Medium SES | 21.79 | 26.49 | 7.46 |
| Low SES | 26.93 | 31.41 | 18.70 |
| CMM → Death (10 years) |  |  |  |
| High SES | 22.90 | 24.88 | 16.95 |
| Medium SES | 33.10 | 52.46 | 35.33 |
| Low SES | 43.87 | 51.49 | 44.11 |
| CMM → Death (15 years) |  |  |  |
| High SES | 24.90 | 31.85 | 16.95 |
| Medium SES | 40.27 | 61.03 | 35.33 |
| Low SES | 55.15 | 58.94 | 54.11 |

Notes: SES, socioeconomic status; FCMD, first-onset cardiometabolic disease; CMM, cardiometabolic multimorbidity; SES, socioeconomic status.

Table S10. Time advancement (years) for low-SES individuals reaching the equivalent 10-year cumulative risk of high-SES individuals across three cohorts.

| **Transitions** | **Mean time**  (years) | **UKB** | **SHARE** | **KLoSA** |
| --- | --- | --- | --- | --- |
|  |  | Time (years) | Time (years) | Time (years) |
| Health → FCMD | 6.2 | 8.2 | 3.5 | 6.8 |
| FCMD → CMM | 4.3 | 5.7 | 3.6 | 3.7 |
| Health → Death | 6.0 | 5.4 | 5.7 | 7.1 |
| FCMD → Death | 5.7 | 6.5 | 5.5 | 5.1 |
| CMM → Death | 5.6 | 6.2 | 5.5 | 5.3 |

Notes: SES, socioeconomic status; FCMD, first-onset cardiometabolic disease; CMM, cardiometabolic multimorbidity.

Table S11. Associations of SES levels with multiple-state transitions from health to FCMD.

| Transitions | meta | | | UKB | | SHARE | | KLoSA | |
| --- | --- | --- | --- | --- | --- | --- | --- | --- | --- |
|  | HR [95%CI] | *P* | *P*_heterogeneity_ | HR [95%CI] | *P* | HR [95%CI] | *P* | HR [95%CI] | *P* |
| Health → FCMD |  |  |  |  |  |  |  |  |  |
| High SES | 1 (reference) | |  | 1 (reference) | | 1 (reference) | | 1 (reference) | |
| Medium SES | 1.16 [1.06-1.29] | 0.002 | 0.005 | 1.14 [1.11-1.17] | 6.64×10^-21^ | 1.06 [0.99-1.14] | 0.110 | 1.36 [1.19-1.54] | 2.73×10^-6^ |
| Low SES | 1.33 [1.30-1.37] | 4.86×10^-95^ | 0.073 | 1.35 [1.31-1.39] | 1.11×10^-86^ | 1.22 [1.12-1.32] | 4.10×10^-6^ | 1.33 [1.16-1.53] | 7.60×10^-5^ |
| *P* for trend |  | 1.21×10^-79^ |  |  | 7.88×10^-106^ |  | 2.00×10^-6^ |  | 1.43×10^-4^ |
| Health → T2D |  |  |  |  |  |  |  |  |  |
| High SES | 1 (reference) | |  | 1 (reference) | | 1 (reference) | | 1 (reference) | |
| Medium SES | 1.22 [1.17-1.27] | 1.31×10^-20^ | 0.054 | 1.21 [1.15-1.26] | 1.14×10^-15^ | 1.15 [1.02-1.31] | 0.025 | 1.43 [1.21-1.69] | 3.44×10^-5^ |
| Low SES | 1.45 [1.39-1.52] | 2.72×10^-62^ | 0.890 | 1.45 [1.38-1.52] | 6.66×10^-50^ | 1.45 [1.26-1.66] | 7.72×10^-8^ | 1.48 [1.23-1.79] | 4.63×10^-5^ |
| *P* for trend |  | 5.13×10^-76^ |  |  | 3.77×10^-59^ |  | 1.97×10^-8^ |  | 5.97×10^-5^ |
| Health → CAD |  |  |  |  |  |  |  |  |  |
| High SES | 1 (reference) | |  | 1 (reference) | | 1 (reference) | | 1 (reference) | |
| Medium SES | 1.10 [1.06-1.14] | 1.02×10^-6^ | 0.072 | 1.11 [1.06-1.15] | 7.38×10^-7^ | 1.00 [0.89-1.11] | 0.929 | 1.29 [0.99-1.67] | 0.055 |
| Low SES | 1.25 [1.20-1.30] | 6.90×10^-28^ | 0.186 | 1.27 [1.22-1.33] | 3.22×10^-27^ | 1.12 [1.00-1.26] | 0.073 | 1.17 [0.88-1.56] | 0.288 |
| *P* for trend |  | 1.38×10^-27^ |  |  | 1.39 ×10^-32^ |  | 0.059 |  | 0.333 |
| Health → Stroke |  |  |  |  |  |  |  |  |  |
| High SES | 1 (reference) | |  | 1 (reference) | | 1 (reference) | | 1 (reference) | |
| Medium SES | 1.09 [1.02-1.16] | 0.011 | 0.748 | 1.08 [1.01-1.16] | 0.025 | 1.08 [0.90-1.30] | 0.400 | 1.24 [0.93-1.65] | 0.140 |
| Low SES | 1.20 [0.98-1.45] | 0.071 | 0.043 | 1.34 [1.24-1.45] | 6.59×10^-14^ | 1.01 [0.81-1.25] | 0.930 | 1.21 [0.89-1.65] | 0.231 |
| *P* for trend |  | 0.111 |  |  | 4.51×10^-19^ |  | 0.936 |  | 0.264 |

Notes: SES, socioeconomic status; FCMD, first-onset cardiometabolic disease; CMM, cardiometabolic multimorbidity.

Table S12. Associations of SES levels with multiple-state transitions from FCMD to CMM.

| Transitions | meta | | | UKB | | SHARE | | KLoSA | |
| --- | --- | --- | --- | --- | --- | --- | --- | --- | --- |
|  | HR [95%CI] | *P* | *P*_heterogeneity_ | HR [95%CI] | *P* | HR [95%CI] | *P* | HR [95%CI] | *P* |
| FCMD → CMM |  |  |  |  |  |  |  |  |  |
| High SES | 1 (reference) | |  | 1 (reference) | | 1 (reference) | | 1 (reference) | |
| Medium SES | 1.13 [1.05-1.22] | 0.002 | 0.578 | 1.15 [1.05-1.25] | 0.002 | 1.04 [0.88-1.23] | 0.633 | 1.11 [0.77-1.58] | 0.582 |
| Low SES | 1.33 [1.23-1.44] | 3.35×10^-13^ | 0.509 | 1.36 [1.25-1.49] | 6.96×10^-12^ | 1.21 [1.01-1.44] | 0.043 | 1.39 [0.96-2.03] | 0.085 |
| *P* for trend |  | 1.77×10^-18^ |  |  | 8.20×10^-17^ |  | 0.032 |  | 0.083 |
| T2D → CMM |  |  |  |  |  |  |  |  |  |
| High SES | 1 (reference) | |  | 1 (reference) | | 1 (reference) | | 1 (reference) | |
| Medium SES | 1.10 [0.99-1.22] | 0.063 | 0.157 | 1.15 [1.03-1.29] | 0.015 | 0.97 [0.75-1.24] | 0.785 | 0.70 [0.41-1.19] | 0.186 |
| Low SES | 1.30 [1.18-1.44] | 3.30×10^-7^ | 0.650 | 1.32 [1.18-1.48] | 2.95×10^-6^ | 1.29 [1.00-1.68] | 0.057 | 0.99 [0.58-1.71] | 0.983 |
| *P* for trend |  | 1.64×10^-8^ |  |  | 4.83×10^-8^ |  | 0.025 |  | 0.795 |
| CAD → CMM |  |  |  |  |  |  |  |  |  |
| High SES | 1 (reference) | |  | 1 (reference) | | 1 (reference) | | 1 (reference) | |
| Medium SES | 1.07 [0.94-1.21] | 0.298 | 0.144 | 1.04 [0.89-1.20] | 0.637 | 1.05 [0.82-1.36] | 0.688 | 2.05 [1.08-3.90] | 0.029 |
| Low SES | 1.30 [0.93-1.82] | 0.130 | 0.034 | 1.32 [1.13-1.54] | 3.48×10^-4^ | 0.98 [0.73-1.31] | 0.881 | 2.61 [1.29-5.29] | 0.008 |
| *P* for trend |  | 0.101 |  |  | 1.22×10^-7^ |  | 0.824 |  | 0.009 |
| Stroke → CMM |  |  |  |  |  |  |  |  |  |
| High SES | 1 (reference) | |  | 1 (reference) | | 1 (reference) | | 1 (Reference) | |
| Medium SES | 1.35 [1.08-1.69] | 0.009 | 0.519 | 1.47 [1.11-1.94] | 0.007 | 1.08 [0.70-1.69] | 0.725 | 1.45 [0.67-3.14] | 0.344 |
| Low SES | 1.51 [1.19-1.93] | 8.01×10^-4^ | 0.829 | 1.59 [1.18-2.12] | 0.002 | 1.38 [0.83-2.30] | 0.217 | 1.27 [0.56-2.91] | 0.566 |
| *P* for trend |  | 0.002 |  |  | 0.007 |  | 0.208 |  | 0.568 |

Notes: SES, socioeconomic status; FCMD, first-onset cardiometabolic disease; CMM, cardiometabolic multimorbidity.

Table S13. Associations of SES levels with multiple-state transitions from health to death.

| Transitions | meta | | | UKB | | SHARE | | KLoSA | |
| --- | --- | --- | --- | --- | --- | --- | --- | --- | --- |
|  | HR [95%CI] | *P* | *P*_heterogeneity_ | HR [95%CI] | *P* | HR [95%CI] | *P* | HR [95%CI] | *P* |
| Health → Death |  |  |  |  |  |  |  |  |  |
| High SES | 1 (reference) | |  | 1 (reference) | | 1 (reference) | | 1 (reference) | |
| Medium SES | 1.23 [1.17-1.28] | 1.82×10^-17^ | 0.799 | 1.21 [1.15-1.28] | 2.11×10^-13^ | 1.26 [1.13-1.40] | 1.65×10^-5^ | 1.23 [1.04-1.44] | 0.012 |
| Low SES | 1.69 [1.61-1.77] | 2.73×10^-104^ | 0.256 | 1.72 [1.63-1.82] | 8.59×10^-83^ | 1.67 [1.49-1.87] | 3.25×10^-19^ | 1.49 [1.27-1.76] | 1.07×10^-6^ |
| *P* for trend |  | 1.88×10^-22^ |  |  | 9.50×10^-114^ |  | 1.58×10^-20^ |  | 9.39×10^-8^ |

Notes: SES, socioeconomic status; FCMD, first-onset cardiometabolic disease; CMM, cardiometabolic multimorbidity.

Table S14. Associations of SES levels with multiple-state progression in transitions from FCMD to death.

| Transitions | meta | | | UKB | | SHARE | | KLoSA | |
| --- | --- | --- | --- | --- | --- | --- | --- | --- | --- |
|  | HR [95%CI] | *P* | *P*_heterogeneity_ | HR [95%CI] | *P* | HR [95%CI] | *P* | HR [95%CI] | *P* |
| FCMD → Death |  |  |  |  |  |  |  |  |  |
| High SES | 1 (reference) | |  | 1 (reference) | | 1 (reference) | | 1 (reference) | |
| Medium SES | 1.06 [0.97-1.15] | 0.190 | 0.589 | 1.04 [0.95-1.14] | 0.407 | 1.15 [0.94-1.41] | 0.165 | 0.98 [0.65-1.50] | 0.937 |
| Low SES | 1.41 [1.30-1.54] | 3.36×10^-15^ | 0.079 | 1.40 [1.27-1.54] | 1.18×10^-11^ | 1.62 [1.31-2.01] | 8.78×10^-6^ | 0.98 [0.66-1.47] | 0.938 |
| *P* for trend |  | 3.06×10^-4^ |  |  | 1.83×10^-24^ |  | 9.38×10^-7^ |  | 0.949 |
| T2D → Death |  |  |  |  |  |  |  |  |  |
| High SES | 1 (reference) | |  | 1 (reference) | | 1 (reference) | | 1 (reference) | |
| Medium SES | 1.05 [0.90-1.22] | 0.545 | 0.909 | 1.05 [0.89-1.24] | 0.581 | 0.99 [0.66-1.48] | 0.953 | 1.12 [0.56-2.25] | 0.746 |
| Low SES | 1.29 [1.11-1.51] | 0.001 | 0.863 | 1.30 [1.09-1.54] | 0.003 | 1.32 [0.87-2.02] | 0.195 | 1.07 [0.54-2.12] | 0.848 |
| *P* for trend |  | 9.35×10^-6^ |  |  | 1.62×10^-5^ |  | 0.108 |  | 0.400 |
| CAD → Death |  |  |  |  |  |  |  |  |  |
| High SES | 1 (reference) | |  | 1 (reference) | | 1 (reference) | | 1 (reference) | |
| Medium SES | 1.10 [0.96-1.26] | 0.174 | 0.717 | 1.11 [0.95-1.30] | 0.190 | 1.11 [0.85-1.46] | 0.440 | 0.83 [0.36-1.92] | 0.657 |
| Low SES | 1.59 [1.38-1.83] | 1.01×10^-10^ | 0.235 | 1.65 [1.40-1.94] | 1.39×10^-9^ | 1.51 [1.13-2.03] | 0.005 | 0.81 [0.36-1.84] | 0.619 |
| *P* for trend |  | 3.19×10^-20^ |  |  | 3.39×10^-19^ |  | 0.002 |  | 0.641 |
| Stroke → Death |  |  |  |  |  |  |  |  |  |
| High SES | 1 (reference) | |  | 1 (reference) | | 1 (reference) | | 1 (reference) | |
| Medium SES | 1.07 [0.92-1.25] | 0.403 | 0.228 | 1.01 [0.85-1.19] | 0.928 | 1.55 [0.97-2.46] | 0.066 | 0.98 [0.47-2.06] | 0.968 |
| Low SES | 1.68 [0.95-2.96] | 0.074 | 0.005 | 1.26 [1.06-1.50] | 0.008 | 2.99 [1.82-4.89] | 1.35×10^-5^ | 1.25 [0.64-2.44] | 0.513 |
| *P* for trend |  | 0.047 |  |  | 7.00×10^-5^ |  | 2.02×10^-6^ |  | 0.430 |

Notes: SES, socioeconomic status; FCMD, first-onset cardiometabolic disease; CMM, cardiometabolic multimorbidity

Table S15. Associations of SES levels with multi-state transitions from CMM to death.

| Transitions | meta | | | UKB | | SHARE | | KLoSA | |
| --- | --- | --- | --- | --- | --- | --- | --- | --- | --- |
|  | HR [95%CI] | *P* | *P*_heterogeneity_ | HR [95%CI] | *P* | HR [95%CI] | *P* | HR [95%CI] | *P* |
| CMM → Death |  |  |  |  |  |  |  |  |  |
| High SES | 1 (reference) | |  | 1 (reference) | | 1 (reference) | | 1 (reference) | |
| Medium SES | 1.48 [1.22-1.79] | 7.05×10^-5^ | 0.192 | 1.34 [1.07-1.69] | 0.010 | 2.02 [1.39-2.93] | 2.30×10^-4^ | 1.03 [0.31-3.45] | 0.926 |
| Low SES | 1.72 [1.41-2.09] | 5.21×10^-8^ | 0.212 | 1.62 [1.29-2.03] | 3.24×10^-5^ | 2.30 [1.54-3.42] | 4.41×10^-5^ | 0.83 [0.24-2.89] | 0.771 |
| *P* for trend |  | 2.65×10^-9^ |  |  | 1.06×10^-6^ |  | 2.03×10^-4^ |  | 0.660 |

Notes: SES, socioeconomic status; FCMD, first-onset cardiometabolic disease; CMM, cardiometabolic multimorbidity; T2D, type 2 diabetes; CAD, coronary artery disease.

Table S16. Estimates and 95% confidence intervals of the RMST difference (in years) stratified by transitions and SES.

| Transitions | meta | | | UKB |  | SHARE | | KLoSA | |
| --- | --- | --- | --- | --- | --- | --- | --- | --- | --- |
|  | RMST [95%CI] | *P* | *P*_heterogeneity_ | RMST [95%CI] | *P* | RMST [95%CI] | *P* | RMST [95%CI] | *P* |
| Health → FCMD |  |  |  |  |  |  |  |  |  |
| High SES | Reference | |  | Reference | | Reference | | Reference | |
| Medium SES | -0.20 [-0.48 to 0.08] | 0.161 | 3.10×10^-15^ | -0.05 [-0.06 to -0.04] | 2.20×10^-16^ | -0.07 [-0.15 to 0.01] | 0.104 | -0.49 [-0.59 to -0.38] | 2.20×10^-16^ |
| Low SES | -0.47 [-0.79 to -0.16] | 0.003 | 4.00×10^-14^ | -0.57 [-0.60 to -0.55] | 2.20×10^-16^ | -0.15 [-0.26 to -0.05] | 5.00×10^-3^ | -0.68 [-0.80 to -0.57] | 2.20×10^-16^ |
| FCMD → CMM |  |  |  |  |  |  |  |  |  |
| High SES | Reference | |  | Reference | | Reference | | Reference | |
| Medium SES | -0.26 [-0.33 to -0.19] | 1.40×10^-13^ | 0.616 | -0.26 [-0.33 to -0.19] | 2.67×10^-13^ | -0.26 [-0.60 to 0.09] | 0.141 | -0.02 [-0.50 to 0.46] | 0.934 |
| Low SES | -0.76 [-1.14 to -0.37] | 1.20×10^-4^ | 0.009 | -1.04 [-1.11 to -0.97] | 2.20×10^-16^ | -0.54 [-0.95 to -0.14] | 0.008 | -0.49 [-1.03 to 0.04] | 0.072 |
| Health → Death |  |  |  |  |  |  |  |  |  |
| High SES | Reference | |  | Reference | | Reference | | Reference | |
| Medium SES | -0.23 [-0.55 to 0.09] | 0.158 | 7.10×10^-42^ | -0.02 [-0.02 to -0.01] | 1.11×10^-16^ | -0.12 [-0.19 to -0.06] | 2.00×10^-4^ | -0.55 [-0.63 to -0.47] | 2.20×10^-16^ |
| Low SES | -0.12 [-0.13 to -0.10] | 6.10×10^-59^ | 0.129 | -0.12 [-0.13 to -0.10] | 2.20×10^-16^ | -0.20 [-0.29 to -0.12] | 2.15×10^-6^ | -0.13 [-0.30 to 0.04] | 0.148 |
| FCMD → Death |  |  |  |  |  |  |  |  |  |
| High SES | Reference | |  | Reference | | Reference | | Reference | |
| Medium SES | -0.18 [-0.23 to -0.12] | 1.30×10^-9^ | 0.429 | -0.18 [-0.23 to -0.12] | 2.65×10^-9^ | -0.25 [-0.53 to 0.04] | 0.091 | 0.11 [-0.36 to 0.58] | 0.635 |
| Low SES | -0.57 [-1.38 to 0.25] | 0.176 | 1.30×10^-4^ | -1.03 [-1.10 to -0.97] | 2.20×10^-16^ | -0.87 [-1.23 to -0.52] | 1.41×10^-6^ | 0.37 [-0.29 to 1.03] | 0.273 |
| CMM → Death |  |  |  |  |  |  |  |  |  |
| High SES | Reference | |  | Reference | | Reference | | Reference | |
| Medium SES | -1.39 [-1.67 to -1.12] | 2.80×10^-23^ | 0.586 | -1.39 [-1.68 to -1.10] | 2.20×10^-16^ | -1.71 [-2.68 to -0.74] | 5.47×10^-4^ | -0.81 [-2.22 to 0.59] | 0.257 |
| Low SES | -1.57 [-2.41 to -0.73] | 2.40×10^-4^ | 0.006 | -1.03 [-1.10 to -0.97] | 2.20×10^-16^ | -2.08 [-2.75 to -1.40] | 1.67×10^-9^ | -2.11 [-4.01 to -0.22] | 0.029 |

Notes: SES, socioeconomic status; FCMD, first-onset cardiometabolic disease; CMM, cardiometabolic multimorbidity.

Table S17. Effect of SES stratified by age on the transitions of CMM in the UKB cohort.

|  | **Age<60 years** | | | | | **Age≥60 years** | | | | |
| --- | --- | --- | --- | --- | --- | --- | --- | --- | --- | --- |
| **Transitions** | **High SES** | **Medium SES** | | **Low SES** | | **High SES** | **Medium SES** | | **Low SES** | |
|  | Ref. | HR [95%CI] | *P* | HR [95%CI] | *P* | Ref. | HR [95%CI] | *P* | HR [95%CI] | *P* |
| Health → FCMD | 1 | 1.15 [1.11-1.20] | <0.001 | 1.46 [1.40-1.52] | <0.001 | 1 | 1.07 [1.02-1.12] | 0.003 | 1.23 [1.17-1.28] | <0.001 |
| Health → Death | 1 | 1.20 [1.12-1.29] | <0.001 | 1.95 [1.80-2.11] | <0.001 | 1 | 1.16 [1.08-1.26] | <0.001 | 1.55 [1.44-1.68] | <0.001 |
| FCMD → CMM | 1 | 1.23 [1.09-1.40] | <0.001 | 1.53 [1.34-1.74] | <0.001 | 1 | 1.05 [0.93-1.18] | 0.441 | 1.21 [1.07-1.37] | 0.002 |
| FCMD → Death | 1 | 1.06 [0.92-1.23] | 0.417 | 1.54 [1.32-1.80] | <0.001 | 1 | 1.00 [0.89-1.14] | 0.941 | 1.31 [1.16-1.49] | <0.001 |
| CMM → Death | 1 | 1.72 [1.11-2.67] | 0.016 | 2.65 [1.70-4.14] | <0.001 | 1 | 1.17 [0.90-1.52] | 0.248 | 1.31 [1.00-1.70] | 0.047 |

Notes: Ref., reference; HR, hazard ratio; CI, confidence interval; FCMD, first-onset cardiometabolic disease; CMM, cardiometabolic multimorbidity; SES, socioeconomic status. Model adjusted for age, sex, smoke status, alcohol consumption, physical activity, BMI, blood pressure, sleep mode.

Table S18. Effect of SES stratified by age on the transitions of CMM in the SHARE cohort.

|  | **Age<60 years** | | | | | **Age≥60 years** | | | | |
| --- | --- | --- | --- | --- | --- | --- | --- | --- | --- | --- |
| **Transitions** | **High SES** | **Medium SES** | | **Low SES** | | **High SES** | **Medium SES** | | **Low SES** | |
|  | Ref. | HR [95%CI] | *P* | HR [95%CI] | *P* | Ref. | HR [95%CI] | *P* | HR [95%CI] | *P* |
| Health → FCMD | 1 | 1.02 [0.93-1.12] | 0.684 | 1.15 [1.03-1.27] | 0.01 | 1 | 1.07 [0.95-1.21] | 0.255 | 1.32 [1.15-1.51] | <0.001 |
| Health → Death | 1 | 1.20 [1.06-1.35] | 0.003 | 1.63 [1.43-1.84] | <0.001 | 1 | 1.45 [1.16-1.82] | 0.001 | 1.72 [1.33-2.22] | <0.001 |
| FCMD → CMM | 1 | 0.98 [0.80-1.19] | 0.838 | 1.09 [0.87-1.35] | 0.461 | 1 | 1.10 [0.82-1.48] | 0.530 | 1.46 [1.06-2.01] | 0.020 |
| FCMD → Death | 1 | 1.11 [0.89-1.38] | 0.349 | 1.56 [1.24-1.96] | <0.001 | 1 | 1.42 [0.82-2.47] | 0.207 | 2.14 [1.20-3.82] | 0.010 |
| CMM → Death | 1 | 2.17 [1.45-3.25] | <0.001 | 2.23 [1.44-3.45] | <0.001 | 1 | 0.86 [0.30-2.46] | 0.773 | 2.22 [0.83-5.91] | 0.112 |

Notes: Ref., reference; HR, hazard ratio; CI, confidence interval; FCMD, first-onset cardiometabolic disease; CMM, cardiometabolic multimorbidity; SES, socioeconomic status. Model adjusted for age, sex, smoke status, alcohol consumption, physical activity, BMI, hypertension, sleep mode.

Table S19. Effect of SES stratified by age on the transitions of CMM in the KLoSA cohort.

|  | **Age<60 years** | | | | | **Age≥60 years** | | | | |
| --- | --- | --- | --- | --- | --- | --- | --- | --- | --- | --- |
| **Transitions** | **High SES** | **Medium SES** | | **Low SES** | | **High SES** | **Medium SES** | | **Low SES** | |
|  | Ref. | HR [95%CI] | *P* | HR [95%CI] | *P* | Ref. | HR [95%CI] | *P* | HR [95%CI] | *P* |
| Health → FCMD | 1 | 1.31 [1.11-1.56] | 0.002 | 1.16 [0.93-1.45] | 0.185 | 1 | 1.14 [0.94-1.39] | 0.191 | 1.23 [1.01-1.50] | 0.037 |
| Health → Death | 1 | 1.38 [1.07-1.77] | 0.012 | 1.97 [1.46-2.65] | <0.001 | 1 | 1.33 [1.06-1.67] | 0.014 | 1.57 [1.26-1.95] | <0.001 |
| FCMD → CMM | 1 | 1.16 [0.69-1.93] | 0.576 | 1.35 [0.69-2.62] | 0.377 | 1 | 0.88 [0.53-1.47] | 0.619 | 1.18 [0.73-1.93] | 0.498 |
| FCMD → Death | 1 | 2.20 [0.71-6.76] | 0.170 | 2.12 [0.50-8.97] | 0.305 | 1 | 0.83 [0.53-1.31] | 0.418 | 0.88 [0.58-1.32] | 0.526 |
| CMM → Death | 1 | 0.97 [0.07-13.37] | 0.983 | 0.81 [0.02-28.89] | 0.906 | 1 | 0.96 [0.23-3.88] | 0.949 | 0.78 [0.20-3.10] | 0.721 |

Notes: Ref., reference; HR, hazard ratio; CI, confidence interval; FCMD, first-onset cardiometabolic disease; CMM, cardiometabolic multimorbidity; SES, socioeconomic status. Model adjusted for age, sex, smoke status, alcohol consumption, physical activity, BMI, hypertension, sleep mode.

Table S20. Meta-analysis of SES and CMM transitions stratified by age in three cohorts.

|  | **Age<60 years** | | | | | **Age≥60 years** | | | | |
| --- | --- | --- | --- | --- | --- | --- | --- | --- | --- | --- |
| **Transitions** | **High SES** | **Medium SES** | | **Low SES** | | **High SES** | **Medium SES** | | **Low SES** | |
|  | Ref. | HR [95%CI] | *P* | HR [95%CI] | *P* | Ref. | HR [95%CI] | *P* | HR [95%CI] | *P* |
| Health → FCMD | 1 | 1.15 [1.11-1.19] | <0.001 | 1.44 [1.38-1.49] | <0.001 | 1 | 1.06 [1.02-1.11] | 0.003 | 1.22 [1.17-1.27] | <0.001 |
| Health → Death | 1 | 1.23 [1.15-1.31] | <0.001 | 1.93 [1.79-2.08] | <0.001 | 1 | 1.18 [1.11-1.26] | <0.001 | 1.57 [1.47-1.67] | <0.001 |
| FCMD → CMM | 1 | 1.21 [1.08-1.35] | 0.001 | 1.51 [1.34-1.71] | <0.001 | 1 | 1.02 [0.93-1.13] | 0.634 | 1.18 [1.06-1.31] | 0.002 |
| FCMD → Death | 1 | 1.09 [0.95-1.26] | 0.213 | 1.58 [1.36-1.83] | <0.001 | 1 | 1.01 [0.91-1.13] | 0.791 | 1.32 [1.19-1.47] | <0.001 |
| CMM → Death | 1 | 1.53 [1.03-2.29] | 0.036 | 2.53 [1.69-3.79] | <0.001 | 1 | 1.48 [0.88-2.47] | 0.140 | 1.48 [1.19-1.86] | <0.001 |

Notes: Ref., reference; HR, hazard ratio; CI, confidence interval; FCMD, first-onset cardiometabolic disease; CMM, cardiometabolic multimorbidity; SES, socioeconomic status. Model adjusted for age, sex, smoke status, alcohol consumption, physical activity, BMI, hypertension, sleep mode.

Table S21. Effect of SES stratified by gender on the transitions of CMM in the UKB cohort.

|  | **Male** | | | | | **Female** | | | | |
| --- | --- | --- | --- | --- | --- | --- | --- | --- | --- | --- |
| **Transitions** | **High SES** | **Medium SES** | | **Low SES** | | **High SES** | **Medium SES** | | **Low SES** | |
|  | Ref. | HR [95%CI] | *P* | HR [95%CI] | *P* | Ref. | HR [95%CI] | *P* | HR [95%CI] | *P* |
| Health → FCMD | 1 | 1.14 [1.11-1.18] | <0.001 | 1.33 [1.29-1.38] | <0.001 | 1 | 1.15 [1.09-1.21] | <0.001 | 1.38 [1.31-1.46] | <0.001 |
| Health → Death | 1 | 1.24 [1.16-1.32] | <0.001 | 1.91 [1.78-2.06] | <0.001 | 1 | 1.16 [1.07-1.26] | <0.001 | 1.48 [1.36-1.62] | <0.001 |
| FCMD → CMM | 1 | 1.16 [1.05-1.28] | 0.003 | 1.37 [1.24-1.51] | <0.001 | 1 | 1.12 [0.94-1.35] | 0.209 | 1.33 [1.11-1.60] | 0.002 |
| FCMD → Death | 1 | 1.13 [1.01-1.26] | 0.040 | 1.63 [1.45-1.83] | <0.001 | 1 | 0.83 [0.70-0.98] | 0.028 | 0.98 [0.82-1.16] | 0.804 |
| CMM → Death | 1 | 1.15 [0.98-1.61] | 0.077 | 1.58 [1.23-2.04] | <0.001 | 1 | 1.79 [1.04-3.09] | 0.037 | 1.89 [1.10-3.26] | 0.022 |

Notes: Ref., reference; HR, hazard ratio; CI, confidence interval; FCMD, first-onset cardiometabolic disease; CMM, cardiometabolic multimorbidity; SES, socioeconomic status. Model adjusted for age, sex, smoke status, alcohol consumption, physical activity, BMI, blood pressure, sleep mode.

Table S22. Effect of SES stratified by gender on the transitions of CMM in the SHARE cohort.

|  | **Male** | | | | | **Female** | | | | |
| --- | --- | --- | --- | --- | --- | --- | --- | --- | --- | --- |
| **Transitions** | **High SES** | **Medium SES** | | **Low SES** | | **High SES** | **Medium SES** | | **Low SES** | |
|  | Ref. | HR [95%CI] | *P* | HR [95%CI] | *P* | Ref. | HR [95%CI] | *P* | HR [95%CI] | *P* |
| Health → FCMD | 1 | 1.05 [0.95-1.16] | 0.362 | 1.19 [1.06-1.34] | 0.004 | 1 | 1.08 [0.96-1.21] | 0.185 | 1.25 [1.11-1.41] | <0.001 |
| Health → Death | 1 | 1.37 [1.19-1.57] | <0.001 | 1.78 [1.52-2.07] | <0.001 | 1 | 1.12 [0.96-1.32] | 0.152 | 1.54 [1.30-1.82] | <0.001 |
| FCMD → CMM | 1 | 1.11 [0.90-1.37] | 0.345 | 1.41 [1.11-1.79] | 0.005 | 1 | 0.96 [0.74-1.26] | 0.784 | 1.06 [0.80-1.40] | 0.673 |
| FCMD → Death | 1 | 1.26 [0.98-1.63] | 0.072 | 1.84 [1.39-2.44] | <0.001 | 1 | 0.97 [0.70-1.35] | 0.858 | 1.33 [0.95-1.86] | 0.099 |
| CMM → Death | 1 | 2.01 [1.27-3.17] | 0.003 | 2.35 [1.42-3.89] | <0.001 | 1 | 1.80 [0.94-3.43] | 0.076 | 1.87 [0.96-3.64] | 0.067 |

Notes: Ref., reference; HR, hazard ratio; CI, confidence interval; FCMD, first-onset cardiometabolic disease; CMM, cardiometabolic multimorbidity; SES, socioeconomic status. Model adjusted for age, sex, smoke status, alcohol consumption, physical activity, BMI, hypertension, sleep mode.

Table S23. Effect of SES stratified by gender on the transitions of CMM in the KLoSA cohort.

|  | **Male** | | | | | **Female** | | | | |
| --- | --- | --- | --- | --- | --- | --- | --- | --- | --- | --- |
| **Transitions** | **High SES** | **Medium SES** | | **Low SES** | | **High SES** | **Medium SES** | | **Low SES** | |
|  | Ref. | HR [95%CI] | *P* | HR [95%CI] | *P* | Ref. | HR [95%CI] | *P* | HR [95%CI] | *P* |
| Health → FCMD | 1 | 1.45 [1.21-1.72] | <0.001 | 1.41 [1.14-1.74] | 0.002 | 1 | 1.25 [1.03-1.51] | 0.022 | 1.25 [1.03-1.52] | 0.023 |
| Health → Death | 1 | 1.24 [1.02-1.52] | 0.033 | 1.56 [1.26-1.94] | <0.001 | 1 | 1.25 [0.95-1.64] | 0.114 | 1.49 [1.14-1.94] | 0.003 |
| FCMD → CMM | 1 | 1.08 [0.69-1.69] | 0.731 | 1.33 [0.78-2.28] | 0.297 | 1 | 1.11 [0.62-2.01] | 0.725 | 1.45 [0.82-2.56] | 0.207 |
| FCMD → Death | 1 | 1.10 [0.67-1.78] | 0.715 | 0.82 [0.50-1.34] | 0.422 | 1 | 0.97 [0.41-2.29] | 0.939 | 1.29 [0.58-2.88] | 0.533 |
| CMM → Death | 1 | 0.97 [0.23-4.14] | 0.967 | 0.92 [0.20-4.22] | 0.917 | 1 | 1.09 [0.11-10.64] | 0.938 | 0.68 [0.07-6.58] | 0.742 |

Notes: Ref., reference; HR, hazard ratio; CI, confidence interval; FCMD, first-onset cardiometabolic disease; CMM, cardiometabolic multimorbidity; SES, socioeconomic status. Model adjusted for age, sex, smoke status, alcohol consumption, physical activity, BMI, hypertension, sleep mode.

Table S24. Meta-analysis of SES and CMM transitions stratified by gender in three cohorts.

|  | **Male** | | | | | **Female** | | | | |
| --- | --- | --- | --- | --- | --- | --- | --- | --- | --- | --- |
| **Transitions** | **High SES** | **Medium SES** | | **Low SES** | | **High SES** | **Medium SES** | | **Low SES** | |
|  | Ref. | HR [95%CI] | *P* | HR [95%CI] | *P* | Ref. | HR [95%CI] | *P* | HR [95%CI] | *P* |
| Health → FCMD | 1 | 1.17 [1.04-1.32] | 0.010 | 1.32 [1.28-1.37] | <0.001 | 1 | 1.14 [1.09-1.20] | <0.001 | 1.35 [1.29-1.42] | <0.001 |
| Health → Death | 1 | 1.26 [1.19-1.34] | <0.001 | 1.86 [1.75-1.98] | <0.001 | 1 | 1.16 [1.08-1.24] | <0.001 | 1.50 [1.39-1.61] | <0.001 |
| FCMD → CMM | 1 | 1.15 [1.05-1.25] | 0.002 | 1.37 [1.25-1.51] | <0.001 | 1 | 1.07 [0.93-1.24] | 0.347 | 1.26 [1.08-1.46] | 0.002 |
| FCMD → Death | 1 | 1.15 [1.03-1.27] | 0.009 | 1.46 [1.06-2.00] | 0.020 | 1 | 0.86 [0.74-1.00] | 0.044 | 1.05 [0.90-1.22] | 0.525 |
| CMM → Death | 1 | 1.39 [1.12-1.72] | 0.003 | 1.69 [1.35-2.11] | <0.001 | 1 | 1.76 [1.17-2.66] | 0.007 | 1.82 [1.20-2.75] | 0.005 |

Notes: Ref., reference; HR, hazard ratio; CI, confidence interval; FCMD, first-onset cardiometabolic disease; CMM, cardiometabolic multimorbidity; SES, socioeconomic status. Model adjusted for age, sex, smoke status, alcohol consumption, physical activity, BMI, hypertension, sleep mode.

Table S25. Effect of SES stratified by smoking status on the transitions of CMM in the UKB cohort.

|  | **Former or current smoker** | | | | | **Never** | | | | |
| --- | --- | --- | --- | --- | --- | --- | --- | --- | --- | --- |
| **Transitions** | **High SES** | **Medium SES** | | **Low SES** | | **High SES** | **Medium SES** | | **Low SES** | |
|  | Ref. | HR [95%CI] | *P* | HR [95%CI] | *P* | Ref. | HR [95%CI] | *P* | HR [95%CI] | *P* |
| Health → FCMD | 1 | 1.17 [1.13-1.22] | <0.001 | 1.40 [1.34-1.46] | <0.001 | 1 | 1.15 [1.10-1.20] | <0.001 | 1.40 [1.34-1.46] | <0.001 |
| Health → Death | 1 | 1.20 [1.12-1.29] | <0.001 | 1.81 [1.67-1.95] | <0.001 | 1 | 1.29 [1.19-1.40] | <0.001 | 1.93 [1.77-2.11] | <0.001 |
| FCMD → CMM | 1 | 1.10 [0.98-1.24] | 0.096 | 1.25 [1.11-1.40] | <0.001 | 1 | 1.17 [1.04-1.32] | 0.011 | 1.36 [1.20-1.54] | <0.001 |
| FCMD → Death | 1 | 1.16 [1.01-1.32] | 0.032 | 1.61 [1.41-1.85] | <0.001 | 1 | 1.14 [0.98-1.32] | 0.079 | 1.57 [1.36-1.82] | <0.001 |
| CMM → Death | 1 | 1.34 [1.02-1.78] | 0.043 | 1.68 [1.26-1.23] | <0.001 | 1 | 1.45 [1.03-2.03] | 0.031 | 1.93 [1.38-2.71] | <0.001 |

Notes: Ref., reference; HR, hazard ratio; CI, confidence interval; FCMD, first-onset cardiometabolic disease; CMM, cardiometabolic multimorbidity; SES, socioeconomic status. Model adjusted for age, sex, smoke status, alcohol consumption, physical activity, BMI, blood pressure, sleep mode.

Table S26. Effect of SES stratified by smoking status on the transitions of CMM in the SHARE cohort.

|  | **Former or current smoker** | | | | | **Never** | | | | |
| --- | --- | --- | --- | --- | --- | --- | --- | --- | --- | --- |
| **Transitions** | **High SES** | **Medium SES** | | **Low SES** | | **High SES** | **Medium SES** | | **Low SES** | |
|  | Ref. | HR [95%CI] | *P* | HR [95%CI] | *P* | Ref. | HR [95%CI] | *P* | HR [95%CI] | *P* |
| Health → FCMD | 1 | 1.02 [0.92-1.13] | 0.688 | 1.17 [1.03-1.32] | 0.014 | 1 | 1.10 [0.99-1.23] | 0.076 | 1.27 [1.13-1.42] | <0.001 |
| Health → Death | 1 | 1.31 [1.14-1.50] | <0.001 | 1.61 [1.37-1.88] | <0.001 | 1 | 1.22 [1.04-1.44] | 0.015 | 1.71 [1.45-2.02] | <0.001 |
| FCMD → CMM | 1 | 0.99 [0.79-1.23] | 0.902 | 0.95 [0.73-1.24] | 0.710 | 1 | 1.15 [0.89-1.49] | 0.280 | 1.50 [1.15-1.96] | 0.003 |
| FCMD → Death | 1 | 1.24 [0.95-1.62] | 0.119 | 1.61 [1.19-2.17] | 0.002 | 1 | 1.04 [0.76-1.41] | 0.819 | 1.58 [1.15-2.16] | 0.004 |
| CMM → Death | 1 | 1.80 [1.11-2.93] | 0.017 | 2.56 [1.49-4.40] | <0.001 | 1 | 2.23 [1.23-4.05] | 0.008 | 2.14 [1.16-3.94] | 0.015 |

Notes: Ref., reference; HR, hazard ratio; CI, confidence interval; FCMD, first-onset cardiometabolic disease; CMM, cardiometabolic multimorbidity; SES, socioeconomic status. Model adjusted for age, sex, smoke status, alcohol consumption, physical activity, BMI, hypertension, sleep mode.

Table S27. Effect of SES stratified by smoking status on the transitions of CMM in the KLoSA cohort.

|  | **Former or current smoker** | | | | | **Never** | | | | |
| --- | --- | --- | --- | --- | --- | --- | --- | --- | --- | --- |
| **Transitions** | **High SES** | **Medium SES** | | **Low SES** | | **High SES** | **Medium SES** | | **Low SES** | |
|  | Ref. | HR [95%CI] | *P* | HR [95%CI] | *P* | Ref. | HR [95%CI] | *P* | HR [95%CI] | *P* |
| Health → FCMD | 1 | 1.62 [1.31-2.00] | <0.001 | 1.44 [1.11-1.87] | 0.007 | 1 | 1.22 [1.04-1.43] | 0.015 | 1.27 [1.07-1.50] | 0.006 |
| Health → Death | 1 | 1.18 [0.92-1.52] | 0.184 | 1.74 [1.34-2.25] | <0.001 | 1 | 1.27 [1.03-1.57] | 0.024 | 1.40 [1.14-1.73] | 0.001 |
| FCMD → CMM | 1 | 1.11 [0.66-1.88] | 0.698 | 1.35 [0.71-2.57] | 0.368 | 1 | 1.10 [0.67-1.78] | 0.710 | 1.36 [0.84-2.20] | 0.205 |
| FCMD → Death | 1 | 1.05 [0.59-1.87] | 0.870 | 0.79 [0.43-1.47] | 0.465 | 1 | 0.97 [0.52-1.78] | 0.910 | 1.18 [0.68-2.04] | 0.558 |
| CMM → Death | 1 | 3.03 [0.35-26.21] | 0.314 | 3.79 [0.41-35.02] | 0.241 | 1 | 0.40 [0.08-2.06] | 0.275 | 0.24 [0.05-1.16] | 0.076 |

Notes: Ref., reference; HR, hazard ratio; CI, confidence interval; FCMD, first-onset cardiometabolic disease; CMM, cardiometabolic multimorbidity; SES, socioeconomic status. Model adjusted for age, sex, smoke status, alcohol consumption, physical activity, BMI, hypertension, sleep mode.

Table S28. Meta-analysis of SES and CMM transitions stratified by smoking status in three cohorts.

|  | **Former or current smoker** | | | | | **Never** | | | | |
| --- | --- | --- | --- | --- | --- | --- | --- | --- | --- | --- |
| **Transitions** | **High SES** | **Medium SES** | | **Low SES** | | **High SES** | **Medium SES** | | **Low SES** | |
|  | Ref. | HR [95%CI] | *P* | HR [95%CI] | *P* | Ref. | HR [95%CI] | *P* | HR [95%CI] | *P* |
| Health → FCMD | 1 | 1.21 [1.02-1.42] | 0.025 | 1.32 [1.15-1.51] | <0.001 | 1 | 1.15 [1.10-1.19] | <0.001 | 1.38 [1.32-1.43] | <0.001 |
| Health → Death | 1 | 1.22 [1.15-1.30] | <0.001 | 1.77 [1.65-1.89] | <0.001 | 1 | 1.28 [1.19-1.37] | <0.001 | 1.70 [1.42-2.03] | <0.001 |
| FCMD → CMM | 1 | 1.07 [0.97-1.19] | 0.165 | 1.15 [0.94-1.41] | 0.172 | 1 | 1.16 [1.05-1.29] | 0.005 | 1.38 [1.24-1.54] | <0.001 |
| FCMD → Death | 1 | 1.17 [1.04-1.31] | 0.009 | 1.45 [1.10-1.91] | 0.008 | 1 | 1.11 [0.98-1.27] | 0.112 | 1.55 [1.36-1.76] | <0.001 |
| CMM → Death | 1 | 1.46 [1.14-1.86] | 0.002 | 1.90 [1.42-2.56] | <0.001 | 1 | 1.54 [1.15-2.06] | 0.003 | 1.47 [0.71-3.02] | 0.295 |

Notes: Ref., reference; HR, hazard ratio; CI, confidence interval; FCMD, first-onset cardiometabolic disease; CMM, cardiometabolic multimorbidity; SES, socioeconomic status. Model adjusted for age, sex, smoke status, alcohol consumption, physical activity, BMI, hypertension, sleep mode.

Table S29. Effect of SES stratified by alcohol consumption on the transitions of CMM in the UKB cohort.

|  | **More than once per week** | | | | | **Less than once per week** | | | | |
| --- | --- | --- | --- | --- | --- | --- | --- | --- | --- | --- |
| **Transitions** | **High SES** | **Medium SES** | | **Low SES** | | **High SES** | **Medium SES** | | **Low SES** | |
|  | Ref. | HR [95%CI] | *P* | HR [95%CI] | *P* | Ref. | HR [95%CI] | *P* | HR [95%CI] | *P* |
| Health → FCMD | 1 | 1.15 [1.11-1.18] | <0.001 | 1.35 [1.31-1.40] | <0.001 | 1 | 1.11 [1.05-1.18] | <0.001 | 1.32 [1.24-1.40] | <0.001 |
| Health → Death | 1 | 1.20 [1.14-1.27] | <0.001 | 1.81 [1.70-1.93] | <0.001 | 1 | 1.28 [1.13-1.45] | <0.001 | 1.61 [1.42-1.82] | <0.001 |
| FCMD → CMM | 1 | 1.12 [1.02-1.24] | 0.023 | 1.33 [1.20-1.47] | <0.001 | 1 | 1.23 [1.03-1.47] | 0.022 | 1.46 [1.22-1.74] | <0.001 |
| FCMD → Death | 1 | 1.05 [0.94-1.16] | 0.394 | 1.46 [1.31-1.63] | <0.001 | 1 | 1.09 [0.88-1.36] | 0.422 | 1.37 [1.10-1.70] | 0.004 |
| CMM → Death | 1 | 1.38 [1.07-1.79] | 0.014 | 1.71 [1.32-2.23] | <0.001 | 1 | 1.16 [0.72-1.86] | 0.537 | 1.33 [0.83-2.12] | 0.237 |

Notes: Ref., reference; HR, hazard ratio; CI, confidence interval; FCMD, first-onset cardiometabolic disease; CMM, cardiometabolic multimorbidity; SES, socioeconomic status. Model adjusted for age, sex, smoke status, alcohol consumption, physical activity, BMI, blood pressure, sleep mode.

Table S30. Effect of SES stratified by alcohol consumption on the transitions of CMM in the SHARE cohort.

|  | **More than once per week** | | | | | **Less than once per week** | | | | |
| --- | --- | --- | --- | --- | --- | --- | --- | --- | --- | --- |
| **Transitions** | **High SES** | **Medium SES** | | **Low SES** | | **High SES** | **Medium SES** | | **Low SES** | |
|  | Ref. | HR [95%CI] | *P* | HR [95%CI] | *P* | Ref. | HR [95%CI] | *P* | HR [95%CI] | *P* |
| Health → FCMD | 1 | 1.09 [0.88-1.35] | 0.437 | 1.10 [0.85-1.43] | 0.472 | 1 | 0.98 [0.85-1.12] | 0.753 | 1.22 [1.07-1.41] | 0.004 |
| Health → Death | 1 | 1.10 [0.80-1.50] | 0.562 | 1.71 [1.21-2.42] | 0.002 | 1 | 1.28 [1.05-1.55] | 0.014 | 1.79 [1.48-2.17] | <0.001 |
| FCMD → CMM | 1 | 1.40 [0.83-2.35] | 0.205 | 1.27 [0.69-2.33] | 0.449 | 1 | 0.90 [0.67-1.20] | 0.461 | 1.23 [0.92-1.64] | 0.166 |
| FCMD → Death | 1 | 1.71 [0.84-3.46] | 0.137 | 1.13 [0.50-2.53] | 0.771 | 1 | 0.89 [0.62-1.26] | 0.503 | 1.33 [0.94-1.90] | 0.111 |
| CMM → Death | 1 | 1.81 [0.59-5.58] | 0.301 | 2.78 [0.82-9.43] | 0.100 | 1 | 1.51 [0.78-2.92] | 0.224 | 1.84 [0.96-3.53] | 0.067 |

Notes: Ref., reference; HR, hazard ratio; CI, confidence interval; FCMD, first-onset cardiometabolic disease; CMM, cardiometabolic multimorbidity; SES, socioeconomic status. Model adjusted for age, sex, smoke status, alcohol consumption, physical activity, BMI, hypertension, sleep mode.

Table S31. Effect of SES stratified by sleep patterns on the transitions of CMM in the UKB cohort.

|  | **Unhealthy** | | | | | **Healthy** | | | | |
| --- | --- | --- | --- | --- | --- | --- | --- | --- | --- | --- |
| **Transitions** | **High SES** | **Medium SES** | | **Low SES** | | **High SES** | **Medium SES** | | **Low SES** | |
|  | Ref. | HR [95%CI] | *P* | HR [95%CI] | *P* | Ref. | HR [95%CI] | *P* | HR [95%CI] | *P* |
| Health → FCMD | 1 | 1.15 [1.10-1.20] | <0.001 | 1.40 [1.34-1.46] | <0.001 | 1 | 1.13 [1.09-1.17] | <0.001 | 1.29 [1.24-1.35] | <0.001 |
| Health → Death | 1 | 1.29 [1.19-1.40] | <0.001 | 1.93 [1.77-2.11] | <0.001 | 1 | 1.16 [1.09-1.24] | <0.001 | 1.56 [1.46-1.68] | <0.001 |
| FCMD → CMM | 1 | 1.17 [1.04-1.32] | 0.011 | 1.36 [1.20-1.54] | <0.001 | 1 | 1.13 [1.00-1.27] | 0.054 | 1.37 [1.21-1.56] | <0.001 |
| FCMD → Death | 1 | 1.14 [0.98-1.32] | 0.079 | 1.57 [1.36-1.82] | <0.001 | 1 | 0.97 [0.86-1.10] | 0.657 | 1.27 [1.11-1.44] | <0.001 |
| CMM → Death | 1 | 1.45 [1.03-2.03] | 0.031 | 1.93 [1.38-2.71] | <0.001 | 1 | 1.25 [0.92-1.70] | 0.152 | 1.35 [0.99-1.84] | 0.057 |

Notes: Ref., reference; HR, hazard ratio; CI, confidence interval; FCMD, first-onset cardiometabolic disease; CMM, cardiometabolic multimorbidity; SES, socioeconomic status. Model adjusted for age, sex, smoke status, alcohol consumption, physical activity, BMI, blood pressure, sleep mode.

Table S32. Effect of SES stratified by sleep patterns on the transitions of CMM in the SHARE cohort.

|  | **Unhealthy** | | | | | **Healthy** | | | | |
| --- | --- | --- | --- | --- | --- | --- | --- | --- | --- | --- |
| **Transitions** | **High SES** | **Medium SES** | | **Low SES** | | **High SES** | **Medium SES** | | **Low SES** | |
|  | Ref. | HR [95%CI] | *P* | HR [95%CI] | *P* | Ref. | HR [95%CI] | *P* | HR [95%CI] | *P* |
| Health → FCMD | 1 | 1.13 [0.99-1.30] | 0.076 | 1.24 [1.06-1.44] | 0.006 | 1 | 1.03 [0.94-1.12] | 0.556 | 1.22 [1.10-1.34] | <0.001 |
| Health → Death | 1 | 1.33 [1.08-1.65] | 0.008 | 1.88 [1.50-2.36] | <0.001 | 1 | 1.24 [1.10-1.40] | <0.001 | 1.61 [1.41-1.83] | <0.001 |
| FCMD → CMM | 1 | 0.97 [0.71-1.31] | 0.826 | 1.07 [0.77-1.48] | 0.706 | 1 | 1.09 [0.89-1.32] | 0.409 | 1.31 [1.06-1.63] | 0.014 |
| FCMD → Death | 1 | 1.21 [0.81-1.82] | 0.351 | 1.48 [0.97-2.27] | 0.070 | 1 | 1.13 [0.90-1.43] | 0.295 | 1.73 [1.35-2.21] | <0.001 |
| CMM → Death | 1 | 2.04 [1.02-4.08] | 0.044 | 2.05 [0.97-4.32] | 0.059 | 1 | 1.95 [1.25-3.05] | 0.003 | 2.20 [1.36-3.55] | 0.001 |

Notes: Ref., reference; HR, hazard ratio; CI, confidence interval; FCMD, first-onset cardiometabolic disease; CMM, cardiometabolic multimorbidity; SES, socioeconomic status. Model adjusted for age, sex, smoke status, alcohol consumption, physical activity, BMI, hypertension, sleep mode.

Table S33. Effect of SES stratified by sleep patterns on the transitions of CMM in the KLoSA cohort.

|  | **Unhealthy** | | | | | **Healthy** | | | | |
| --- | --- | --- | --- | --- | --- | --- | --- | --- | --- | --- |
| **Transitions** | **High SES** | **Medium SES** | | **Low SES** | | **High SES** | **Medium SES** | | **Low SES** | |
|  | Ref. | HR [95%CI] | *P* | HR [95%CI] | *P* | Ref. | HR [95%CI] | *P* | HR [95%CI] | *P* |
| Health → FCMD | 1 | 1.62 [1.24-2.12] | <0.001 | 1.38 [1.04-1.84] | 0.027 | 1 | 1.24 [1.07-1.44] | 0.004 | 1.32 [1.12-1.56] | <0.001 |
| Health → Death | 1 | 1.13 [0.82-1.54] | 0.455 | 1.52 [1.12-2.06] | 0.007 | 1 | 1.27 [1.05-1.53] | 0.012 | 1.46 [1.20-1.78] | <0.001 |
| FCMD → CMM | 1 | 0.88 [0.42-1.86] | 0.735 | 1.37 [0.64-2.95] | 0.414 | 1 | 1.20 [0.80-1.80] | 0.391 | 1.28 [0.83-2.00] | 0.268 |
| FCMD → Death | 1 | 0.65 [0.29-1.50] | 0.316 | 0.78 [0.35-1.74] | 0.540 | 1 | 1.14 [0.70-1.87] | 0.594 | 1.05 [0.66-1.67] | 0.847 |
| CMM → Death | 1 | 0.49 [0.04-6.55] | 0.589 | 0.29 [0.02-4.37] | 0.372 | 1 | 1.21 [0.29-4.97] | 0.795 | 1.30 [0.31-5.41] | 0.722 |

Notes: Ref., reference; HR, hazard ratio; CI, confidence interval; FCMD, first-onset cardiometabolic disease; CMM, cardiometabolic multimorbidity; SES, socioeconomic status. Model adjusted for age, sex, smoke status, alcohol consumption, physical activity, BMI, hypertension, sleep mode.

Table S34. Meta-analysis of SES and CMM transitions stratified by sleep patterns in three cohorts.

|  | **Unhealthy** | | | | | **Healthy** | | | | |
| --- | --- | --- | --- | --- | --- | --- | --- | --- | --- | --- |
| **Transitions** | **High SES** | **Medium SES** | | **Low SES** | | **High SES** | **Medium SES** | | **Low SES** | |
|  | Ref. | HR [95%CI] | *P* | HR [95%CI] | *P* | Ref. | HR [95%CI] | *P* | HR [95%CI] | *P* |
| Health → FCMD | 1 | 1.21 [1.06-1.38] | 0.005 | 1.39 [1.33-1.45] | <0.001 | 1 | 1.12 [1.09-1.16] | <0.001 | 1.28 [1.23-1.33] | <0.001 |
| Health → Death | 1 | 1.29 [1.20-1.39] | <0.001 | 1.90 [1.76-2.05] | <0.001 | 1 | 1.18 [1.12-1.25] | <0.001 | 1.56 [1.47-1.66] | <0.001 |
| FCMD → CMM | 1 | 1.13 [1.01-1.27] | 0.027 | 1.32 [1.18-1.48] | <0.001 | 1 | 1.12 [1.02-1.24] | 0.022 | 1.35 [1.21-1.50] | <0.001 |
| FCMD → Death | 1 | 1.13 [0.99-1.29] | 0.075 | 1.53 [1.34-1.76] | <0.001 | 1 | 1.01 [0.91-1.12] | 0.864 | 1.34 [1.20-1.49] | <0.001 |
| CMM → Death | 1 | 1.52 [1.13-2.06] | 0.006 | 1.90 [1.40-2.58] | <0.001 | 1 | 1.44 [1.12-1.84] | 0.004 | 1.55 [1.20-2.00] | <0.001 |

Notes: Ref., reference; HR, hazard ratio; CI, confidence interval; FCMD, first-onset cardiometabolic disease; CMM, cardiometabolic multimorbidity; SES, socioeconomic status. Model adjusted for age, sex, smoke status, alcohol consumption, physical activity, BMI, hypertension, sleep mode.

Table S35. Effect of SES stratified by physical activity on the transitions of CMM in the UKB cohort.

|  | **Low-to-moderate** | | | | | **High** | | | | |
| --- | --- | --- | --- | --- | --- | --- | --- | --- | --- | --- |
| **Transitions** | **High SES** | **Medium SES** | | **Low SES** | | **High SES** | **Medium SES** | | **Low SES** | |
|  | Ref. | HR [95%CI] | *P* | HR [95%CI] | *P* | Ref. | HR [95%CI] | *P* | HR [95%CI] | *P* |
| Health → FCMD | 1 | 1.13 [1.09-1.17] | <0.001 | 1.35 [1.30-1.40] | <0.001 | 1 | 1.16 [1.10-1.21] | <0.001 | 1.35 [1.29-1.42] | <0.001 |
| Health → Death | 1 | 1.24 [1.17-1.33] | <0.001 | 1.83 [1.71-1.96] | <0.001 | 1 | 1.15 [1.06-1.26] | 0.001 | 1.54 [1.40-1.69] | <0.001 |
| FCMD → CMM | 1 | 1.15 [1.03-1.27] | 0.009 | 1.37 [1.23-1.52] | <0.001 | 1 | 1.15 [0.99-1.34] | 0.076 | 1.34 [1.14-1.57] | <0.001 |
| FCMD → Death | 1 | 1.07 [0.96-1.20] | 0.228 | 1.45 [1.28-1.63] | <0.001 | 1 | 0.98 [0.83-1.15] | 0.780 | 1.31 [1.10-1.55] | 0.002 |
| CMM → Death | 1 | 1.50 [1.14-1.97] | 0.004 | 1.81 [1.38-2.39] | <0.001 | 1 | 1.04 [0.70-1.55] | 0.845 | 1.23 [0.83-1.84] | 0.307 |

Notes: Ref., reference; HR, hazard ratio; CI, confidence interval; FCMD, first-onset cardiometabolic disease; CMM, cardiometabolic multimorbidity; SES, socioeconomic status. Model adjusted for age, sex, smoke status, alcohol consumption, physical activity, BMI, blood pressure, sleep mode.

Table S36. Effect of SES stratified by physical activity on the transitions of CMM in the SHARE cohort.

|  | **Low-to-moderate** | | | | | **High** | | | | |
| --- | --- | --- | --- | --- | --- | --- | --- | --- | --- | --- |
| **Transitions** | **High SES** | **Medium SES** | | **Low SES** | | **High SES** | **Medium SES** | | **Low SES** | |
|  | Ref. | HR [95%CI] | *P* | HR [95%CI] | *P* | Ref. | HR [95%CI] | *P* | HR [95%CI] | *P* |
| Health → FCMD | 1 | 1.25 [1.08-1.45] | 0.003 | 1.36 [1.16-1.59] | <0.001 | 1 | 1.25 [1.08-1.45] | 0.003 | 1.36 [1.16-1.59] | <0.001 |
| Health → Death | 1 | 1.37 [1.12-1.67] | 0.002 | 1.82 [1.48-2.23] | <0.001 | 1 | 1.37 [1.12-1.67] | 0.002 | 1.82 [1.48-2.23] | <0.001 |
| FCMD → CMM | 1 | 1.16 [0.85-1.58] | 0.358 | 1.21 [0.87-1.68] | 0.266 | 1 | 1.16 [0.85-1.58] | 0.358 | 1.21 [0.87-1.68] | 0.266 |
| FCMD → Death | 1 | 1.07 [0.74-1.55] | 0.729 | 1.54 [1.06-2.23] | 0.025 | 1 | 1.07 [0.74-1.55] | 0.729 | 1.54 [1.06-2.23] | 0.025 |
| CMM → Death | 1 | 1.73 [0.87-3.45] | 0.117 | 1.97 [0.94-4.10] | 0.072 | 1 | 1.73 [0.87-3.45] | 0.117 | 1.97 [0.94-4.10] | 0.072 |

Notes: Ref., reference; HR, hazard ratio; CI, confidence interval; FCMD, first-onset cardiometabolic disease; CMM, cardiometabolic multimorbidity; SES, socioeconomic status. Model adjusted for age, sex, smoke status, alcohol consumption, physical activity, BMI, hypertension, sleep mode.

Table S37. Effect of SES stratified by physical activity on the transitions of CMM in the KLoSA cohort.

|  | **Low-to-moderate** | | | | | **High** | | | | |
| --- | --- | --- | --- | --- | --- | --- | --- | --- | --- | --- |
| **Transitions** | **High SES** | **Medium SES** | | **Low SES** | | **High SES** | **Medium SES** | | **Low SES** | |
|  | Ref. | HR [95%CI] | *P* | HR [95%CI] | *P* | Ref. | HR [95%CI] | *P* | HR [95%CI] | *P* |
| Health → FCMD | 1 | 1.26 [1.07-1.50] | 0.007 | 1.21 [0.99-1.47] | 0.058 | 1 | 1.42 [1.16-1.73] | <0.001 | 1.47 [1.20-1.81] | <0.001 |
| Health → Death | 1 | 1.24 [1.00-1.53] | 0.050 | 1.45 [1.17-1.81] | <0.001 | 1 | 1.21 [0.93-1.58] | 0.158 | 1.64 [1.28-2.10] | <0.001 |
| FCMD → CMM | 1 | 1.40 [0.86-2.29] | 0.175 | 1.84 [1.08-3.12] | 0.024 | 1 | 0.82 [0.47-1.43] | 0.477 | 0.96 [0.55-1.67] | 0.885 |
| FCMD → Death | 1 | 0.89 [0.51-1.55] | 0.673 | 0.84 [0.48-1.47] | 0.543 | 1 | 0.93 [0.48-1.80] | 0.821 | 1.10 [0.62-1.97] | 0.745 |
| CMM → Death | 1 | 1.05 [0.21-5.15] | 0.956 | 0.76 [0.14-4.08] | 0.747 | 1 | 0.73 [0.06-8.36] | 0.798 | 1.08 [0.16-7.24] | 0.937 |

Notes: Ref., reference; HR, hazard ratio; CI, confidence interval; FCMD, first-onset cardiometabolic disease; CMM, cardiometabolic multimorbidity; SES, socioeconomic status. Model adjusted for age, sex, smoke status, alcohol consumption, physical activity, BMI, hypertension, sleep mode.

Table S38. Meta-analysis of SES and CMM transitions stratified by physical activity in three cohorts.

|  | **Low-to-moderate** | | | | | **High** | | | | |
| --- | --- | --- | --- | --- | --- | --- | --- | --- | --- | --- |
| **Transitions** | **High SES** | **Medium SES** | | **Low SES** | | **High SES** | **Medium SES** | | **Low SES** | |
|  | Ref. | HR [95%CI] | *P* | HR [95%CI] | *P* | Ref. | HR [95%CI] | *P* | HR [95%CI] | *P* |
| Health → FCMD | 1 | 1.14 [1.10-1.18] | <0.001 | 1.35 [1.30-1.39] | <0.001 | 1 | 1.15 [1.10-1.19] | <0.001 | 1.38 [1.32-1.43] | <0.001 |
| Health → Death | 1 | 1.25 [1.18-1.33] | <0.001 | 1.80 [1.69-1.91] | <0.001 | 1 | 1.28 [1.19-1.37] | <0.001 | 1.70 [1.42-2.03] | <0.001 |
| FCMD → CMM | 1 | 1.16 [1.05-1.28] | 0.003 | 1.37 [1.24-1.51] | <0.001 | 1 | 1.16 [1.05-1.29] | 0.005 | 1.38 [1.24-1.54] | <0.001 |
| FCMD → Death | 1 | 1.06 [0.96-1.18] | 0.256 | 1.43 [1.27-1.60] | <0.001 | 1 | 1.11 [0.98-1.27] | 0.112 | 1.55 [1.36-1.76] | <0.001 |
| CMM → Death | 1 | 1.52 [1.18-1.95] | 0.001 | 1.79 [1.39-2.31] | <0.001 | 1 | 1.54 [1.15-2.06] | 0.003 | 1.47 [0.71-3.02] | 0.295 |

Notes: Ref., reference; HR, hazard ratio; CI, confidence interval; FCMD, first-onset cardiometabolic disease; CMM, cardiometabolic multimorbidity; SES, socioeconomic status. Model adjusted for age, sex, smoke status, alcohol consumption, physical activity, BMI, hypertension, sleep mode.

Table S39. Associations between individual socioeconomic factors with transitions in CMM in the UKB cohort.

| **Transitions** | **Income** | | **Education** | | **Employment** | |
| --- | --- | --- | --- | --- | --- | --- |
|  | HR [95%CI] | *P* | HR [95%CI] | *P* | HR [95%CI] | *P* |
| **Model A** | | | | | | |
| Health → FCMD | 1.19 [1.17-1.21] | <0.001 | 1.03 [1.03-1.04] | <0.001 | 1.39 [1.35-1.44] | <0.001 |
| Health → Death | 1.52 [1.47-1.57] | <0.001 | 1.04 [1.03-1.05] | <0.001 | 2.00 [1.90-2.12] | <0.001 |
| FCMD → CMM | 1.19 [1.14-1.26] | <0.001 | 1.02 [1.01-1.03] | <0.001 | 1.28 [1.18-1.38] | <0.001 |
| FCMD → Death | 1.32 [1.24-1.40] | <0.001 | 1.03 [1.01-1.04] | <0.001 | 1.53 [1.40-1.68] | <0.001 |
| CMM → Death | 1.34 [1.19-1.50] | <0.001 | 1.02 [1.00-1.04] | 0.073 | 1.41 [1.19-1.67] | <0.001 |
| **Model B** | | | | | | |
| Health → T2D | 1.21 [1.19-1.26] | <0.001 | 1.04 [1.03-1.05] | <0.001 | 1.31 [1.25-1.37] | <0.001 |
| Health → CAD | 1.11 [1.10-1.16] | <0.001 | 1.04 [1.03-1.04] | <0.001 | 1.35 [1.28-1.42] | <0.001 |
| Health → Stroke | 1.08 [1.19-1.31] | <0.001 | 1.01 [1.00-1.03] | 0.003 | 1.53 [1.40-1.67] | <0.001 |
| Health → Death | 1.21 [1.47-1.57] | <0.001 | 1.04 [1.03-1.05] | 0.019 | 2.00 [1.90-2.12] | <0.001 |
| T2D → CMM | 1.15 [1.07-1.23] | <0.001 | 1.02 [1.01-1.03] | 0.046 | 1.20 [1.08-1.33] | <0.001 |
| T2D → Death | 1.15 [1.17-1.44] | <0.001 | 1.02 [1.00-1.04] | <0.001 | 1.38 [1.18-1.61] | <0.001 |
| CAD → CMM | 1.05 [1.15-1.38] | <0.001 | 1.02 [1.00-1.04] | <0.001 | 1.48 [1.28-1.70] | <0.001 |
| CAD → Death | 1.04 [1.28-1.54] | <0.001 | 1.05 [1.03-1.07] | <0.001 | 1.89 [1.64-2.19] | <0.001 |
| Stroke → CMM | 1.11[1.03-1.43] | 0.018 | 1.03[1.00-1.06] | 0.054 | 1.42 [1.11-1.81] | 0.006 |
| Stroke → Death | 1.47[1.07-1.32] | 0.002 | 1.01[0.99-1.03] | 0.323 | 1.13 [0.93-1.38] | 0.218 |
| CMM → Death | 1.34[1.19-1.50] | <0.001 | 1.02[1.00-1.04] | 0.073 | 1.41 [1.19-1.67] | <0.001 |

Notes: Ref., reference; HR, hazard ratio; CI, confidence interval; FCMD, first-onset cardiometabolic disease; CMM, cardiometabolic multimorbidity; T2D, type 2 diabetes; CAD, coronary artery disease; SES, socioeconomic status. Model adjusted for age, sex, smoke status, alcohol consumption, physical activity, BMI, blood pressure, sleep mode.

Table S40. Associations between individual socioeconomic factors with transitions in CMM in the SHARE cohort.

| **Transitions** | **Income** | | **Education** | | **Employment** | |
| --- | --- | --- | --- | --- | --- | --- |
|  | HR [95%CI] | *P* | HR [95%CI] | *P* | HR [95%CI] | *P* |
| **Model A** | | | | | | |
| Health → FCMD | 1.07 [1.05-1.10] | <0.001 | 1.09 [1.05-1.14] | <0.001 | 1.10 [1.02-1.18] | 0.015 |
| Health → Death | 1.16 [1.12-1.20] | <0.001 | 1.21 [1.15-1.28] | <0.001 | 1.22 [1.10-1.34] | <0.001 |
| FCMD → CMM | 1.11 [1.04-1.18] | <0.001 | 1.04 [0.95-1.13] | 0.404 | 0.93 [0.79-1.10] | 0.419 |
| FCMD → Death | 1.21 [1.13-1.30] | <0.001 | 1.12 [1.02-1.24] | 0.024 | 1.04 [0.86-1.26] | 0.700 |
| CMM → Death | 1.18 [1.05-1.33] | 0.007 | 1.36 [1.14-1.63] | <0.001 | 1.03 [0.74-1.45] | 0.850 |
| **Model B** | | | | | | |
| Health → T2D | 1.10 [1.06-1.15] | <0.001 | 1.19 [1.12-1.28] | <0.001 | 1.33 [1.18-1.49] | <0.001 |
| Health → CAD | 1.05 [1.01-1.10] | 0.014 | 1.04 [0.98-1.10] | 0.209 | 0.98 [0.87-1.10] | 0.701 |
| Health → Stroke | 1.06 [0.99-1.14] | 0.085 | 1.04 [0.94-1.15] | 0.432 | 0.93 [0.76-1.13] | 0.457 |
| Health → Death | 1.16 [1.12-1.20] | <0.001 | 1.21 [1.15-1.28] | <0.001 | 1.22 [1.10-1.34] | <0.001 |
| T2D → CMM | 1.09 [1.00-1.18] | 0.061 | 1.06 [0.93-1.21] | 0.398 | 1.05 [0.83-1.33] | 0.682 |
| T2D → Death | 1.11 [0.96-1.27] | 0.155 | 1.06 [0.86-1.30] | 0.592 | 1.13 [0.78-1.62] | 0.516 |
| CAD → CMM | 1.09 [0.99-1.20] | 0.080 | 0.97 [0.85-1.11] | 0.672 | 0.76 [0.58-1.00] | 0.051 |
| CAD → Death | 1.23 [1.12-1.36] | <0.001 | 1.10 [0.96-1.26] | 0.162 | 0.99 [0.75-1.29] | 0.930 |
| Stroke → CMM | 1.14 [0.96-1.35] | 0.130 | 1.09 [0.86-1.36] | 0.482 | 0.95 [0.59-1.54] | 0.846 |
| Stroke → Death | 1.33 [1.14-1.55] | <0.001 | 1.26 [1.01-1.57] | 0.043 | 1.21 [0.76-1.92] | 0.427 |
| CMM → Death | 1.18 [1.05-1.33] | 0.007 | 1.36 [1.14-1.63] | <0.001 | 1.03 [0.74-1.45] | 0.850 |

Notes: Ref., reference; HR, hazard ratio; CI, confidence interval; FCMD, first-onset cardiometabolic disease; CMM, cardiometabolic multimorbidity; T2D, type 2 diabetes; CAD, coronary artery disease; SES, socioeconomic status. Model adjusted for age, sex, smoke status, alcohol consumption, physical activity, BMI, hypertension, sleep mode.

Table S41. Associations between individual socioeconomic factors with transitions in CMM in the KLoSA cohort.

| **Transitions** | **Income** | | **Education** | | **Employment** | |
| --- | --- | --- | --- | --- | --- | --- |
|  | HR [95%CI] | *P* | HR [95%CI] | *P* | HR [95%CI] | *P* |
| **Model A** | | | | | | |
| Health → FCMD | 1.07 [1.02-1.11] | 0.004 | 1.12 [1.02-1.22] | 0.012 | 1.04 [0.93-1.17] | 0.471 |
| Health → Death | 1.03 [0.99-1.08] | 0.147 | 1.23 [1.12-1.35] | <0.001 | 1.26 [1.11-1.43] | <0.001 |
| FCMD → CMM | 1.08 [0.97-1.21] | 0.177 | 1.01 [0.81-1.27] | 0.903 | 1.29 [0.94-1.78] | 0.110 |
| FCMD → Death | 1.00 [0.90-1.12] | 0.980 | 1.21 [0.95-1.52] | 0.117 | 0.81 [0.57-1.14] | 0.226 |
| CMM → Death | 1.02 [0.75-1.38] | 0.903 | 1.53 [0.75-3.14] | 0.243 | 1.09 [0.43-2.78] | 0.861 |
| **Model B** | | | | | | |
| Health → T2D | 1.11 [1.05-1.18] | <0.001 | 1.10 [0.98-1.24] | 0.095 | 1.09 [0.93-1.27] | 0.286 |
| Health → CAD | 1.08 [0.99-1.18] | 0.078 | 1.01 [0.85-1.21] | 0.899 | 0.99 [0.78-1.26] | 0.954 |
| Health → Stroke | 0.95 [0.86-1.04] | 0.271 | 1.36 [1.12-1.64] | 0.002 | 0.98 [0.76-1.27] | 0.888 |
| Health → Death | 1.04 [1.00-1.08] | 0.073 | 1.25 [1.13-1.37] | <0.001 | 1.26 [1.11-1.44] | <0.001 |
| T2D → CMM | 1.09 [0.92-1.28] | 0.325 | 0.91 [0.66-1.26] | 0.576 | 1.22 [0.78-1.92] | 0.382 |
| T2D → Death | 1.10 [0.93-1.31] | 0.253 | 1.04 [0.73-1.47] | 0.837 | 0.81 [0.49-1.33] | 0.405 |
| CAD → CMM | 1.16 [0.94-1.42] | 0.161 | 1.26 [0.88-1.82] | 0.210 | 1.63 [0.93-2.86] | 0.091 |
| CAD → Death | 1.01 [0.78-1.31] | 0.947 | 0.93 [0.58-1.48] | 0.752 | 0.61 [0.28-1.36] | 0.229 |
| Stroke → CMM | 1.04 [0.81-1.34] | 0.742 | 0.95 [0.58-1.55] | 0.823 | 0.96 [0.45-2.03] | 0.909 |
| Stroke → Death | 0.95 [0.79-1.15] | 0.605 | 1.80 [1.12-2.87] | 0.014 | 1.12 [0.59-2.11] | 0.728 |
| CMM → Death | 0.99 [0.73-1.34] | 0.938 | 1.45 [0.71-2.94] | 0.304 | 1.04 [0.41-2.65] | 0.927 |

Notes: HR, hazard ratio; CI, confidence interval; FCMD, first-onset cardiometabolic disease; CMM, cardiometabolic multimorbidity; T2D, type 2 diabetes; CAD, coronary artery disease; SES, socioeconomic status. Model adjusted for age, sex, smoke status, alcohol consumption, physical activity, BMI, hypertension, sleep mode.

Table S42. Meta-analysis of associations between individual socioeconomic factors and CMM transitions across three cohorts.

| **Transitions** | **Income** | | **Education** | | **Employment** | |
| --- | --- | --- | --- | --- | --- | --- |
|  | HR [95%CI] | *P* | HR [95%CI] | *P* | HR [95%CI] | *P* |
| **Model A** | | | | | | |
| Health → FCMD | 1.11 [1.02-1.20] | 0.011 | 1.07 [1.02-1.12] | 0.009 | 1.15 [0.92-1.45] | 0.213 |
| Health → Death | 1.22 [0.98-1.53] | 0.076 | 1.16 [1.01-1.32] | 0.031 | 1.48 [1.03-2.11] | 0.032 |
| FCMD → CMM | 1.15 [1.11-1.19] | <0.001 | 1.02 [1.01-1.03] | <0.001 | 1.28 [1.19-1.38] | <0.001 |
| FCMD → Death | 1.18 [1.03-1.35] | 0.018 | 1.03 [1.02-1.04] | <0.001 | 1.02 [0.62-1.70] | 0.931 |
| CMM → Death | 1.24 [1.15-1.35] | <0.001 | 1.20 [0.92-1.56] | 0.182 | 1.38 [1.17-1.63] | <0.001 |
| **Model B** | | | | | | |
| Health → T2D | 1.15 [1.07-1.24] | <0.001 | 1.11 [1.00-1.23] | 0.050 | 1.29 [1.24-1.35] | <0.001 |
| Health → CAD | 1.09 [1.03-1.15] | 0.001 | 1.04 [1.03-1.04] | <0.001 | 1.11 [0.86-1.43] | 0.442 |
| Health → Stroke | 1.08 [0.93-1.27] | 0.319 | 1.08 [0.97-1.21] | 0.160 | 1.13 [0.78-1.64] | 0.526 |
| Health → Death | 1.22 [0.98-1.53] | 0.076 | 1.16 [1.01-1.32] | 0.031 | 1.48 [1.03-2.11] | 0.032 |
| T2D → CMM | 1.12 [1.07-1.18] | <0.001 | 1.02 [1.01-1.03] | 0.003 | 1.17 [1.07-1.29] | <0.001 |
| T2D → Death | 1.20 [1.12-1.29] | <0.001 | 1.02 [1.00-1.04] | 0.017 | 1.29 [1.13-1.48] | <0.001 |
| CAD → CMM | 1.17 [1.10-1.25] | <0.001 | 1.02 [1.00-1.04] | 0.047 | 1.20 [0.73-1.97] | 0.478 |
| CAD → Death | 1.25 [1.09-1.44] | 0.002 | 1.05 [1.03-1.07] | <0.001 | 1.14 [0.63-2.07] | 0.664 |
| Stroke → CMM | 1.15 [1.04-1.28] | 0.009 | 1.03 [1.00-1.06] | 0.047 | 1.27 [1.03-1.57] | 0.024 |
| Stroke → Death | 1.16 [0.98-1.36] | 0.080 | 1.21 [0.93-1.58] | 0.147 | 1.14 [0.96-1.36] | 0.138 |
| CMM → Death | 1.24 [1.15-1.35] | <0.001 | 1.20 [0.92-1.56] | 0.182 | 1.38 [1.17-1.63] | <0.001 |

Notes: HR, hazard ratio; CI, confidence interval; FCMD, first-onset cardiometabolic disease; CMM, cardiometabolic multimorbidity; T2D, type 2 diabetes; CAD, coronary artery disease; SES, socioeconomic status. Model adjusted for age, sex, smoke status, alcohol consumption, physical activity, BMI, hypertension, sleep mode.

Table S43 Association between SES and transitions in CMM after excluding cases diagnosed in the first 3 years in the UKB cohort.

| **Transitions** | **High SES** | **Medium SES** | | **Low SES** | |
| --- | --- | --- | --- | --- | --- |
|  | Ref. | HR [95%CI] | *P* | HR [95%CI] | *P* |
| **Model A** | | | | | |
| Health → FCMD | 1 | 1.13 [1.10-1.17] | <0.001 | 1.34 [1.30-1.39] | <0.001 |
| Health → Death | 1 | 1.19 [1.13-1.26] | <0.001 | 1.67 [1.57-1.77] | <0.001 |
| FCMD → CMM | 1 | 1.15 [1.04-1.28] | 0.005 | 1.36 [1.22-1.51] | <0.001 |
| FCMD → Death | 1 | 1.02 [0.92-1.14] | 0.649 | 1.37 [1.23-1.53] | <0.001 |
| CMM → Death | 1 | 1.36 [1.04-1.78] | 0.026 | 1.54 [1.17-2.02] | 0.002 |
| **Model B** | | | | | |
| Health → T2D | 1 | 1.11 [1.03-1.20] | <0.001 | 1.31 [1.21-1.43] | <0.001 |
| Health → CAD | 1 | 1.19 [1.10-1.29] | <0.001 | 1.40 [1.29-1.53] | <0.001 |
| Health → Stroke | 1 | 1.12 [1.00-1.26] | 0.059 | 1.28 [1.13-1.45] | <0.001 |
| Health → Death | 1 | 1.16 [1.07-1.26] | <0.001 | 1.45 [1.33-1.58] | <0.001 |
| T2D → CMM | 1 | 1.01 [0.80-1.27] | 0.939 | 1.21 [0.96-1.52] | 0.108 |
| T2D → Death | 1 | 0.93 [0.68-1.26] | 0.620 | 0.96 [0.70-1.31] | 0.781 |
| CAD → CMM | 1 | 1.36 [0.95-1.95] | 0.096 | 1.51 [1.05-2.18] | <0.001 |
| CAD → Death | 1 | 0.71 [0.53-0.97] | <0.001 | 0.90 [0.66-1.23] | 0.505 |
| Stroke → CMM | 1 | 1.45 [0.83-2.55] | 0.193 | 1.53 [0.86-2.72] | 0.148 |
| Stroke → Death | 1 | 0.93 [0.71-1.22] | 0.588 | 1.15 [0.86-1.53] | 0.337 |
| CMM → Death | 1 | 1.79 [1.04-3.10] | <0.001 | 1.87 [1.09-3.23] | <0.001 |

Notes: Ref., reference; HR, hazard ratio; CI, confidence interval; FCMD, first-onset cardiometabolic disease; CMM, cardiometabolic multimorbidity; T2D, type 2 diabetes; CAD, coronary artery disease; SES, socioeconomic status. Model adjusted for age, sex, smoke status, alcohol consumption, physical activity, BMI, blood pressure, sleep mode.

Table S44. Association between SES and transitions in CMM after excluding cases diagnosed in the first 3 years in the SHARE cohort.

| **Transitions** | **High SES** | **Medium SES** | | **Low SES** | |
| --- | --- | --- | --- | --- | --- |
|  | Ref. | HR [95%CI] | *P* | HR [95%CI] | *P* |
| **Model A** | | | | | |
| Health → FCMD | 1 | 1.03 [0.94-1.12] | 0.543 | 1.22 [1.11-1.34] | <0.001 |
| Health → Death | 1 | 1.26 [1.13-1.40] | <0.001 | 1.68 [1.50-1.87] | <0.001 |
| FCMD → CMM | 1 | 1.10 [0.90-1.33] | 0.345 | 1.30 [1.05-1.60] | 0.016 |
| FCMD → Death | 1 | 1.17 [0.91-1.49] | 0.221 | 1.59 [1.23-2.07] | <0.001 |
| CMM → Death | 1 | 1.42 [0.90-2.23] | 0.132 | 1.90 [1.18-3.06] | 0.008 |
| **Model B** | | | | | |
| Health → T2D | 1 | 1.13 [0.98-1.30] | 0.086 | 1.46 [1.25-1.70] | <0.001 |
| Health → CAD | 1 | 0.97 [0.86-1.10] | 0.680 | 1.14 [0.99-1.31] | 0.072 |
| Health → Stroke | 1 | 0.97 [0.80-1.18] | 0.768 | 0.94 [0.75-1.20] | 0.639 |
| Health → Death | 1 | 1.26 [1.13-1.40] | <0.001 | 1.68 [1.50-1.87] | <0.001 |
| T2D → CMM | 1 | 1.01 [0.76-1.34] | 0.957 | 1.36 [1.01-1.83] | 0.044 |
| T2D → Death | 1 | 1.18 [0.71-1.96] | 0.512 | 1.34 [0.79-2.28] | 0.283 |
| CAD → CMM | 1 | 1.27 [0.92-1.75] | 0.147 | 1.14 [0.79-1.65] | 0.470 |
| CAD → Death | 1 | 1.05 [0.75-1.47] | 0.786 | 1.50 [1.04-2.15] | 0.028 |
| Stroke → CMM | 1 | 0.88 [0.54-1.44] | 0.617 | 1.41 [0.81-2.46] | 0.228 |
| Stroke → Death | 1 | 1.48 [0.87-2.51] | 0.148 | 2.65 [1.52-4.64] | <0.001 |
| CMM → Death | 1 | 1.42 [0.90-2.23] | 0.132 | 1.90 [1.18-3.06] | 0.008 |

Notes: Ref., reference; HR, hazard ratio; CI, confidence interval; FCMD, first-onset cardiometabolic disease; CMM, cardiometabolic multimorbidity; T2D, type 2 diabetes; CAD, coronary artery disease; SES, socioeconomic status. Model adjusted for age, sex, smoke status, alcohol consumption, physical activity, BMI, hypertension, sleep mode.
